# The Revised Diploid Genome Sequence of an Individual Human: An Optimized Assembly Workflow for Scaling of near Telomere-to-Telomere Assemblies

**DOI:** 10.64898/2026.05.01.26352134

**Authors:** Si Lok, Timothy NH Lau, Amy HY Tong, Brett Trost, Miriam S Reuter, Bhooma Thiruvahindrapuram, Tara Paton, Jeffrey R MacDonald, Lynette Lau, Christian R Marshall, J Craig Venter, Stephen W Scherer

## Abstract

The first draft diploid genome assembly (HuRef) of an individual released in 2007 was a milestone in genomics. Here, we report HuRef2.0, a revision of HuRef assembled using a scalable two-step workflow employing only Oxford Nanopore Technologies (ONT) Simplex reads and the hifiasm assembler. Results are close in continuity to the recent telomere-to-telomere (T2T) assemblies, but were assembled from standard DNA samples without using multiple sequencing and mapping technologies, including ultra-long-reads and/or proximity-ligation. Three ONT flowcells (∼103x coverage) from fresh blood DNA produced an assembly comprising 26 contigs, with gapless assembly of 23 chromosomes. Two gaps on chromosome (Chr)Y were locally assembled to yield the final T2T-assembly, HuRef2.0, with base accuracy >Q60 and 2,393 phase blocks with an NG50 value of 2.36 Mb. Assembly from a single ONT flowcell (∼35x coverage) consistently produced an assembly more contiguous than GRCh38.p14, providing a foundation for further optimization and scaling. Assembly quality was assessed by direct chromosome-level alignments to reference genome, variant calling, and the annotation of gene-rich regions at Chr22q11, the extended *MHC* locus on Chr6, and several difficult to assemble regions of the genome, including the ribosomal RNA gene clusters, the sub-telomeric region on Chr4q35, and ChrY. More accurate but shorter Pacific Biosciences (PacBio) HiFi-reads produced less contiguous assemblies than from equivalent coverage of error-corrected ONT reads, indicating the importance of read-length. Finally, we compared HuRef2.0 to an assembly of an EBV-transformed lymphoblastoid cell line derived from the same donor. We observed no notable structural differences, indicating that low-passage archival transformed cells are reliable sources for genomic analysis.

## Introduction

The initial sequencing and assembly of the human genome was one of the most important achievements in human genomics. It transformed medicine and drove technology development^1–3^, generating an estimated $706 billion US economic activity and 3.8 million job years of employment between 1988 and 2010^4^. Landmark human genome assemblies were independently reported in 2001 by two groups: The Human Genome Consortium^5^, representing the government funded Human Genome Project (HGP) and; Celera Genomics^6^, a business unit of Applera, representing the industrial sector. Both draft assemblies were relatively fragmented by today’s standards and suffered from being composite haploid assemblies from multiple DNA donors. The HGP assembly was successively revised, first with GRCh35 in 2004^7^, leading to the current version, GRCh38.p14 (GCF_000001405.40) in 2022. Likewise, the Celera Genomics assembly was updated by the John Craig Venter Institute (JCVI) in 2007^8^. This update, known as HuRef (Human Reference Genome)^8^ was a notable technological and logistical achievement. Sequence contributions were removed from four of the original five DNA donors that made up the 2001 Celera Genomics assembly^6^. A haplotype-aware assembly strategy was also used, resulting in HuRef being the first diploid genome assembly of an individual, revealing for the first time that genetic variations mostly in the form of insertions and deletions, and larger structural variations existed between individuals are five times higher than previously estimated.

Due to the limitations of the technologies at that time, HuRef was relatively fragmented when compared to the recent gold standard of complete, gap-less, telomere-to-telomere (T2T) assemblies^9–12^, completed 15-20 years later. However, important limitations of the current T2T-assemblies are that they require specially prepared ultra-long DNA samples, costly concomitant use of multiple sequencing and scaffolding platforms, and logistically and computationally challenging cross-platform data integration, making the large-scale implementation of T2T-assemblies extremely challenging.

Here, we report HuRef2.0, the first revision of HuRef since 2007, assembled from a fresh blood DNA sample from the original donor. Building on the recent improvements in Oxford Nanopore Technologies Kit 14 Simplex reads (Oxford Nanopore Technologies, Oxford, UK; herein referred to as ONT reads) and the hifiasm-0.24.0-r702 (hifiasm) assembler^13,14,15^, we created a scalable two-step workflow that produces near-T2T-assemblies for the human genome from typical high-quality DNA samples from fresh blood. We compare this assembly to those generated from Pacific Biosciences (PacBio) HiFi-reads^16^ (Pacific Biosciences, Menlo Park, CA) from the same blood DNA sample and from JCV37772 cells, an EBV-transformed lymphoblastoid cell line derived from the same donor. Freed from the logistical complications and burdens of producing ultra-long-reads or long-range scaffolding information from optical mapping (Bionano Genomics, San Diego, CA) or proximity-ligation (Hi-C: Arima Genomics, Carlsbad, CA; Pore-C: Oxford Nanopore Technologies), our workflow provides a foundation for scalable genome assembly, in turn allowing for the economic generation of pangenome references^17^ or the resolution of clinically relevant complex genomic arrangements that are not readily reconstructed by mapping-based variant techniques.

## Results and Discussions

### Step 1: DNA Purification, DNA Sequencing, and Pre-Assembly Error-Correction

The individual whose DNA gave rise to HuRef^8^ and HuRef2.0 is J. Craig Venter (JCV), born 14 October 1946. The donor gave consent to disclose publicly his genomic data in totality, including relevant personal and family medical and phenotypic traits. DNA from fresh JCV blood and JCV37772 cells had peak lengths of ∼60 kb on the Agilent Tape Station (Agilent Technologies, Santa Clara, CA), typical of high-molecular weight DNAs purified using commercial kits. ONT-reads with quality score less than Q9 or less than 5 kb in length, and PacBio HiFi-reads less than 2 kb in length, were discarded. “Failed HiFi-reads”, which failed PacBio’s internal accuracy standards for accuracy but are typically longer, were used for scaffolding. Similarly, a legacy set of PacBio Continuous Long-Reads (CLRs) from JCV37772 cells, which are typically Q10 in quality but have read-lengths approaching ONT-reads (see Supplementary Information (SI)-2b), were used for scaffolding. Read statistics and genomic coverage at different length categories across the two sequencing platforms are presented in SI-1. While JCV blood and JCV37772 cell DNAs were similar in length (>60 kb), read profiles were different for the ONT and PacBio HiFi platforms, with read-length N50 values of 26.9 and 17.1 kb, respectively. There were very few PacBio HiFi-reads longer than 50 kb (80 reads), compared to 8.9x coverage (439,897 reads) from the ONT platform.

Our assembler of choice, hifiasm^13,14,15^, started the workflow with pre-assembly read error correction^18^, which corrected sequencing errors while retaining allelic differences and repeat divergence to enable assembly through complex regions of the genome^19,20,21,22,23,24^. For regions of low sequence diversity, the length of the reads was important to facilitate this process. The hifiasm assembler^13,14,15^ was particularly suited to this workflow due to its convenient integration with one of the efficient haplotype aware error-correction algorithms^15,25,26,27^, optimized use of ONT or PacBio reads, extensive parameters for fine-tuning the assembly to different regions of the genome, and the capability to accept accurate reads as primary input and less accurate but longer reads as secondary input for scaffolding. Typically, more accurate ONT-reads were used as the primary-reads and less accurate by still long ONT-reads were used for scaffolding. For the PacBio HiFi-assemblies, “failed HiFi-reads” and legacy CLRs from the discontinued PacBio Sequel sequencer were used as secondary input for scaffolding.

For the initial error-correction step, we trimmed less accurate terminal sequences, typically the first and last 100 bp of each ONT-read SI-2a. SI-2b and SI-2c highlight the differences in length distributions, coverage, and quality scores for ONT- and PacBio HiFi-reads used in this study. PacBio HiFi-reads generally had higher quality score than ONT-reads, but were shorter (typically between 10-25 kb). ONT-reads were more variable in length but had significant greater number of reads between 30 to 45 kb. ONT-reads called by the Super High-Accuracy (SUP) base caller had higher Q values than those called by the High-Accuracy (HAC) base caller. Although reads with high Q values are slightly under-reported by the vendor, vendor supplied Q values are generally in line with values directly tabulated against the assembled chromosome (Chr)X (SI-2d).

The contributions of base-calling accuracy (HAC vs SUP), coverage, and the number of rounds of pre-assembly error-correction of ONT-reads on assembly contiguity were determined by aligning the assembled contigs to the T2T-CHM13.v2 reference genome^9^ (See Materials and Methods). As shown in SI-3, the aforementioned variables, independent and in contribution, contributed to the T2T-assembly of specific chromosomes. SUP calls consistently produced more contiguous assemblies than HAC calls, as did more rounds of pre-assembly error-correction, however, these effects were most pronounced at low coverage. Consequently, we adopted the following pre-assembly error-correction regimen for ONT-reads, comprising: (a) 100× coverage (3 ONT flowcells), (b) use of SUP base calls, (c) trim 100 bp from both ends of reads, (d) discard reads < 5 kb or <Q9, and (e) the use of two rounds of hifiasm pre-assembly error correction.

### Step 2: Assembly of Error-Corrected Reads

More than half of the human genome comprised repetitive sequences, with each chromosome bearing unique sets of repeated structures of varying complexity^19,20,21,22,23,24^. It is evident that using the current technologies and algorithms, no one set of assembly parameters can reliably resolve all repeat regions in a single run. In the second step, we assembled error-corrected reads under different assembly parameters in parallel to produce a panel of sub-assemblies. This pragmatic approach collectively derived an optimised assembly solution for each chromosome. The final assembly is a compilation of the best assembled chromosomes amongst the sub-assemblies as determined by their alignment to the T2T-CHM13.v2 reference genome^9^.

The conditions used to generate the fourteen sub-assemblies (A-N) from ONT-reads are depicted in SI-4. Condition A used the default hifiasm parameters (k-mer length (*k*)=51; Minimizer Window Size (*w*)=51; Expected Error Rate (*bw*)=0.05; Upper Frequency Limit (*D*)=5). Conditions B to N had appropriate adjustments made to the input reads or the assembly parameters. When reads were filtered according to their mean quality, the arithmetic mean quality score (amQ) was used, defined as the arithmetic mean of the quality score of each base in the read, as output in the FASTQ file. The read coverage and length distribution stratified by amQ scores is depicted in SI-2d. SI-5 tabulates the resulting assembly metrics for the fourteen sub-assemblies and the final composite-assembly at different genomic coverages. Assembly contiguity was sensitive to coverage, as indicated by the increasing number of T2T-chromosomes in the composite-assemblies: ten T2T-chromosomes assembled from one flowcell (35x), twenty-two T2T-chromosomes from two flowcells (67x), and twenty-three T2T-chromosomes from three flowcells (103x). Assembly of ChrX and ChrY was particularly sensitive to read coverage. Read-accuracy and read-length were important contributory factors, as indicated by the superior performance of sub-assemblies L, M, and N, which applied quality or length filters to the input reads, or the addition of a third round of pre-assembly error-correction. We had kept the pre-assembly error correction (r) to three or fewer rounds to avoid the potential loss of allelic information from over-correction.

Some sub-assembly conditions gave rise to mis-assembly of certain chromosomes, as inferred by alignment to the T2T-CHM13v2.0 reference genome^9^. Mis-assembled chromosomes are depicted by red panels in SI-5, and could indicate relaxed conditions that promote correct assembly through difficult regions of the genome whilst generating artefacts at other locations. The predominant mis-assemblies were chimeric chromosomes, where a segment of one chromosome was joined to another chromosome *via* a shared repetitive element. Chimeras were not found in sub-assemblies D, I, J, or N, nor for Chr3, 4, 5, 7, 8, 11, 12, 16, 18, 19, 20, X and Y. For the remaining eleven chromosomes, the occurrence of chimeras was sporadic amongst the different subassemblies, which is consistent with them being assembly artefacts rather than true translocations in the JCV genome. Since our workflow is tuned for the human genome using the T2T-CHM13v2.0 reference, such assembly artefacts are readily recognised and managed.

The composite-assembly from three ONT flowcells comprised the best-assembled chromosomes from sub-assemblies H, I, J, K, M, N, and F (SI-6). The longest assembled chromosome with the fewest number or the smallest gaps (when aligned to T2T-CHM13v2.0) was selected for the composite-assembly. Where similar quality chromosomes were present in multiple sub-assemblies, the chromosome was selected from the most productive sub-assembly. Amongst the most productive sub-assemblies were those (J, M, and N) using the most accurate fraction of ONT-reads (>amQ40), which necessitated at least 2 flowcells to achieve sufficient coverage.

The final composite-assembly comprised the T2T-assembly of all 22 autosomes and ChrX. ChrY contained two gaps of 351,481 bp and 76,423 bp when aligned to T2T-CHM13v2.0. These gaps were filled by manual curation (see Materials and Methods) to yield the intermediate T2T-assembly, JCVT2T. Compared to the intermediates of the other T2T-assemblies^9–12^, which typically require extensive post-assembly polishing, JCVT2T was relatively free of assembly and residual sequencing errors. When ONT-reads (SUP called reads) were aligned back to the composite-assembly to scan for assembly errors, 133 changes were made to JCVT2T, of which 57 were single-base changes, 59 were indels (<50 bp), and 17 were insertion or deletions involving up to 1500 bp of sequences (see Materials and Methods).

Using Illumina 150 bp paired-end-reads (107x), GATKv4.6.1.0 HaplotypeCaller^28^ identified 80,988 potential residual sequencing errors in the autosomes. Nearly all the discordant positions (80,653; 99.6% of the total) were in homopolymer tracks; 282 were in simple tandem repeats; and 53 were isolated small errors in which alignment with long-reads had missed. The affected homopolymer tracks represented only 0.13% of the total 60.8 million homopolymer tracks >4 bp present in the composite-assembly (SI-7) – a testament to the extremely low residual error rate in unique regions after pre-assembly error-correction. Residual sequencing errors were near zero for homopolymers fewer than 10 bp but were higher for homopolymers between 10 and 15 bp, where 20% of them are affected. The error rate then declined for homopolymers longer than 15 bp, potentially due to decreased GATK variant-calling confidence in these regions possibly due to a combination of mapping difficulties and higher error rates in Illumina-reads spanning longer homopolymers. Consequently, errors in homopolymer tracks longer than 15 bp remained under-corrected. Using the high-confidence GATK calls, we made 80,779 corrections to the composite-assembly, producing the final T2T-assembly HuRef2.0 (NCBI accession JBPPLP01). HuRef2.0 has a Merqury^29^ QV of 60.11 and completeness of 97.77% (k-mer size = 21).

Assembly metrics for HuRef2.0 are comparable with the recent T2T-assemblies^9–12^, and are significantly more contiguous when compared to GRCh38.p14 and HuRef^8^ assembled from Sanger sequencing (Table 1). The difference in assembly size compared to T2T-CHM13v2.0 and HG002 can be attributed to the potential collapse of repeats in the centromeric regions, the ribosomal DNA cluster on Chr13, 14, 15, 21, and 22, and the heterochromatic region on ChrY, all of which vary between individuals^23,24^ and represent some of the few remaining low-confidence regions in the genome, as detailed in the discussion section. HuRef2.0 is more contiguous than JCVblood-PB (NCBI accession: JBTWEK01), assembled from 101x coverage of PacBio HiFi-reads produced from the same blood sample. It is also more contiguous than JCVcellLine-PB (NCBI accession: JBPQOS01), assembled using 97x HiFi-reads and 87x CLR from JCV37772 cells, a low passage EBV-transformed cell line derived from the same blood donor. These findings show that after error mitigation, longer ONT-reads are inherently more efficacious for *de novo* assembly than from shorter but more accurate PacBio HiFi-reads. Of the two PacBio HiFi-assemblies, JCVcellLine-PB is more contiguous, possibly due to the availability of legacy PacBio CLRs used for scaffolding. CLRs are produced by a discontinued PacBio platform (Sequel I or Sequel II) and are similar in length as ONT-reads but are only Q10 in accuracy (SI-1b, SI-1c, SI-2b).

**Table 1:**
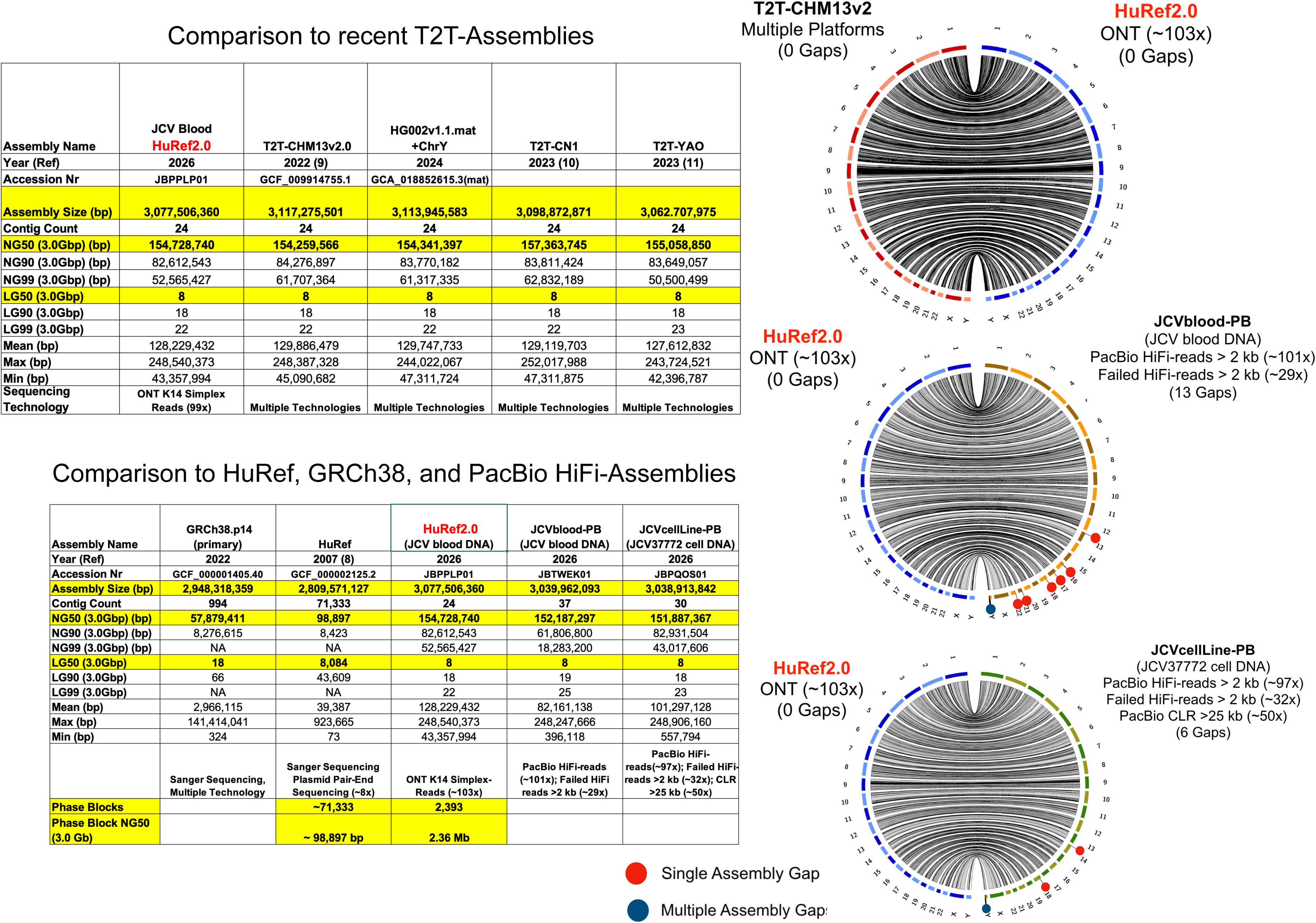
Assembly Metrics. (Left) Assembly Metrics of HuRef2.0 Compared to Recent T2T-Assemblies. HuRef2.0 is much more contiguous than the original HuRef and GRCh38.p14 (primary) and is at par with other T2T-assemblies. (Right) Circos Plots of HuRef2.0 against T2T-CHM13v2.0 and JCVblood-PB and JCVcellLine-PB. All assemblies were concordant across the 24 chromosomes with no visible inter-chromosomal translocations or rearrangements. Single and multiple assembly gaps are denoted by red and blue dots respectively.

The right panel of Table 1 shows Circos plots^30^ comparing HuRef2.0 against T2T-CHM13v2.0 and our two PacBio HiFi-assemblies. Aside from the gaps in the PacBio HiFi-assemblies, all assemblies were concordant across the 24 chromosomes. At least at the ∼400 kb resolution of the Circos plots, we did not observe notable genomic differences between assemblies from JCV37772 cells versus fresh blood, indicating that DNAs from low-passaged EBV-transformed cell lines could be a reliable resource for genomic analysis when DNA from blood is not available. From PacBio HiFi-reads, we were able to reconstruct the genome of the EBV B95-8 strain used for transformation. Ninety percent of the assembled species was identical to the B95-8 reference genome with its characteristic 12-kb deletion, with the remaining virus species harbouring minor SNPs. We found no wildtype EBV sequences in the sample. From the tiling depth, we estimate an average of 5-10 EBV episomes per transformed cell.

### Mitochondrial Genome

HuRef2.0 includes a 16,589 bp mitochondrial genome. Identical mitochondrial genomes were assembled from ONT-, PacBio HiFi-, and Illumina-reads. MITOMAP and MITOMASTER^31^ identified 34 variants (1 insertion and 33 transitions) that placed JCV mitochondrion in Haplogroup K1a3a (SI-8). Aside from localised regions with higher prevalence, mtDNA haplotype K is found in about 6% of the population of Europe and the Near East. The spread of haplotype K from the Near East to Neolithic Europe is believed to stem from demic dispersal of farming populations^32^. Haplogroup K is associated with increased risk of attention deficit and hyperactivity disorder in populations of European ancestry^33^. A recent report showed Haplogroup K is protective for autism spectrum disorder risk^34,35^, however, this finding contradicts earlier reports, which showed either no association^36^ or an increased risk^37^.

### Phasing

While T2T-assembly contiguity was the primary focus for HuRef2.0, we also employed an alignment-based method to phase a large portion of the assembly. Against the HuRef2.0 genome, SNVs and indels called by DeepVariant^38^, and structural variants called by Sniffles2^39^ were phased using WhatsHap^40^. WhatsHap^40^ phased the variants into 2,393 phase blocks, with a phase block NG50 value of 2.36 Mb, spanning 91.1% of the autosomes. Chromosomal distributions of the 2,393 phase blocks and the 2,371-intervening switch-over regions in HuRef2.0 are depicted in SI-9. These metrics for HuRef2.0 are significant improvements (>20-fold) over the 71,464 phase blocks and phase block NG50 value of 98,897 bp for HuRef^8^, contrasting the difference between Sanger dideoxy and ONT sequencing.

As expected, a sizable number of the unresolved switch-over regions in HuRef2.0 resides on the acrocentric chromosomes and the centromeres, where near-identical and lengthy repetitive sequences exceed the reach of our ONT-reads to grow and merge adjacent phase blocks. ONT-reads used to assemble HuRef2.0 had a read-length N50 value of 26.9 kb, placing a limit on the phasing of regions with a sparsity of heterozygous variants. In some switch-over regions, it is possible the other haplotype might be too divergent and therefore eluded reconstruction, as exemplified by recent diploid characterization of the p-arm pericentromere of HG002 Chr16^12^, where the paternal and maternal haplotypes showed extensive and complex rearrangements.

## Assembly Quality

### Extended Major Histocompatibility Complex (MHC)

The quality of HuRef2.0 was assessed by annotating several gene-rich and difficult to assemble regions of the genome and their alignment with T2T-CHM13v2.0. Figure 1a depicts the 7.8 Mb extended MHC region^41^ situated between *SCGN* and *KIFC1* on Chr6. The 260 principal protein coding genes in this gene-rich region in HuRef2.0 are concordant with those in T2T-CHM13v2.0^9^, with only minor differences observed in the lengths of the intergenic regions. The extended MHC region in HuRef2.0 comprised four phase blocks constructed from variant calls made against GRCh38.p14. Pseudo haplotype 1 and 2 are interrupted by switch-over regions of 204,563 bp, 77,629 bp, and 54,352 bp, perhaps due to the sparsity of heterozygous variants in these regions that might have exceeded the spanning capacity of our ONT-reads to grow and merge adjacent phase blocks. ONT-reads used to assemble HuRef2.0 have read-length N50 of 26.9 kb (Mean and Median lengths 20,219 bp and 17,427 bp, respectively). Whilst these reads are sufficient to produce a near-T2T haploid assembly, their ability to span and phase extended regions of low heterozygosity is limited.

**Figure 1:**
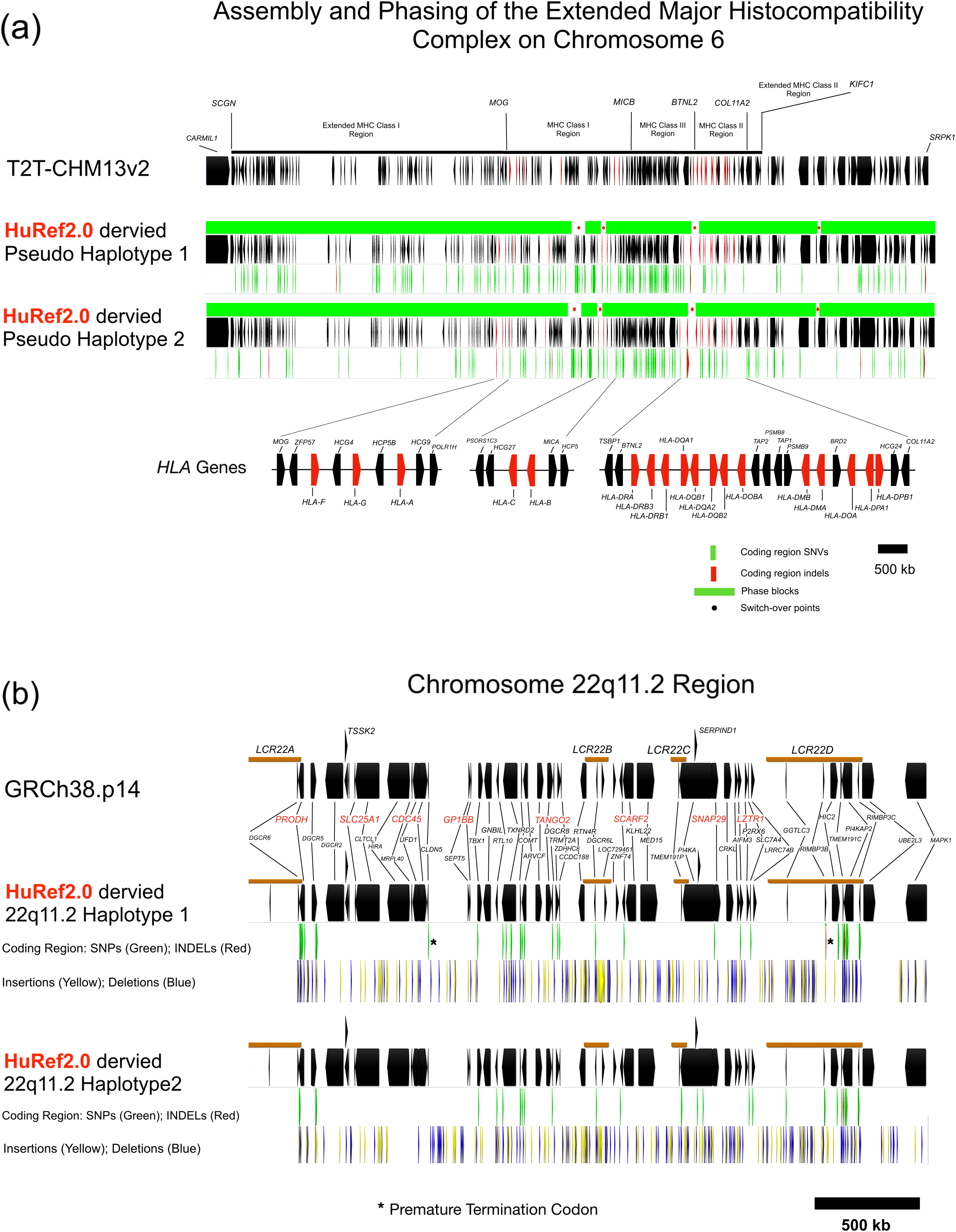
HuRef2.0 Extended *MHC* and Chr22q11DS Regions. (a) Haplotype Construction of the Extended *MHC* Region. The region is composed of 5 separate phase blocks denoted by green bar. Switch-over regions are denoted by black dots. Genes are denoted in black. *HLA* genes are highlighted in red. Green strips below the gene row denote coding SNVs compared to T2T-CHM13v2.0. Red strips denote coding region indels compared to T2T-CHM13v2.0. (b) Haplotype construction of the Chr22q11.2 region. The region between *DGCR6* and *MAPK1* is haplotype-resolved and contains no switch-over regions. Coding region SNPs and indels compared to GRCh38.p14 are denoted by green and red strips, respectively. Insertions and deletions compared to GRCh38.p14 are denoted by yellow and blue strips, respectively. Low-copy repeats (LCRs) are indicated by brown bars. Asterisk (*) denotes premature termination codon in haplotype 1 of *CLDN5* and *RIMBP3B*. (Sequences can be downloaded at https://doi.org/10.5281/zenodo.19861724).

#### Chr22q11.2-Deletion Syndrome Region

The Chr22q11.2-deletion syndrome (Chr22q11DS) region^42,43^ in HuRef2.0 is shown in Figure 1b. Defining features of this region are four low-copy-number repeats (*LCR22A* to *LCR22D*), where substantial sequence similarity is believed to facilitate meiotic recombination or cross-over, resulting in the unbalanced deletions and reciprocal duplications of large segments of DNA at this locus. Typical Chr22q11DS results from recurrent microdeletions (0.7 to 3 Mb), encompassing multiple genes, whose combined loss contributes to the syndrome’s variable and heterogeneous clinical manifestations. Our workflow has successfully assembled through the four low-copy-number repeats (*LCR22A* to *LCR22D*). In HuRef2.0, all 50 principal protein coding genes are present and are concordant with those on GRCh38.p14. The entire Chr22q11DS region resides on a 7 Mb phase block allowing the complete reconstruction of both haplotypes, showing the utility of using whole genome *de novo* assembly for high-resolution characterization of this syndrome.

While large multigene deletions indicative of Chr22q11DS were not observed in HuRef2.0, we did observed variants causing the premature translation termination of two genes. *CLDN5* is an essential component of tight junctions and is critical for the integrity of the brain barrier with roles in CNS disorders^44^. In haplotype 1, a common SNP (rs885985; G-to-A transition at CRCh38.p14 position chr2219524402) encodes a stop codon that truncates the translation of the longest isoform of CLDN5, thereby limiting translation initiation to a downstream AUG that produces a much smaller protein of 218 aa. The smaller protein appears to be the predominant species. *CLDN5* on haplotype 2 is competent to produce both protein isoforms.

Also, on haplotype 1, a C-C dinucleotide insertion at HuRef2.0 position chr2220000664 creates an in-frame termination codon four nucleotides downstream from the insertion site, producing a truncated RIMBP3B polypeptide of 654 aa of unknown significance compared to the 4,918 aa polypeptide produced by the common *RIMBP3B* alleles on haplotype 2. *RIMBP3B* is predicted to enable benzodiazepine receptor binding and is believed to be involved in fertilization and spermatid development. RIMBP3B is produced from a single large exon, hence it is not possible to bypass the lesion by alternative splicing. Unlike CLDN5, there are no reports of the use of alternative downstream translation initiation to produce a functional protein.

### ChrY

A schematics of ChrY in HuRef2.0 is presented in Figure 2. ChrY had been challenging to assemble due to highly repetitive sequences that include long palindromes, tandem repeats and segmental duplications. After filling two gaps (see Materials and Methods), our workflow produced a complete ChrY assembly comparable to recent T2T-ChrYs^45,46^. HuRef2.0 ChrY is 52 Mb in length. Compared to T2T-CHM13v2.0, HuRef2.0 ChrY shared large syntenic blocks in the euchromatic region, interspersed with notable inversions and translocations, particularly in the *DAZ* region where the four *DAZ* genes are inverted and translocated when compared to their CHM13 counterparts. As observed for the other assembled ChrYs, the heterochromatic regions displayed considerable heterogeneity between individuals^45^ and is amongst the most challenging region to assemble. The varied lengths of ChrY between individuals (45 to 85 Mb) are mostly due to heterogeneity in the heterochromatic regions, whilst the euchromatic regions showed comparatively little size variations^44,45^. The heterochromatic region in HuRef2.0 spanned nearly 20 Mb and showed good concordance with tiled ONT-reads. However, due to the complexity of this region further validation is warranted with longer reads.

**Figure 2.**
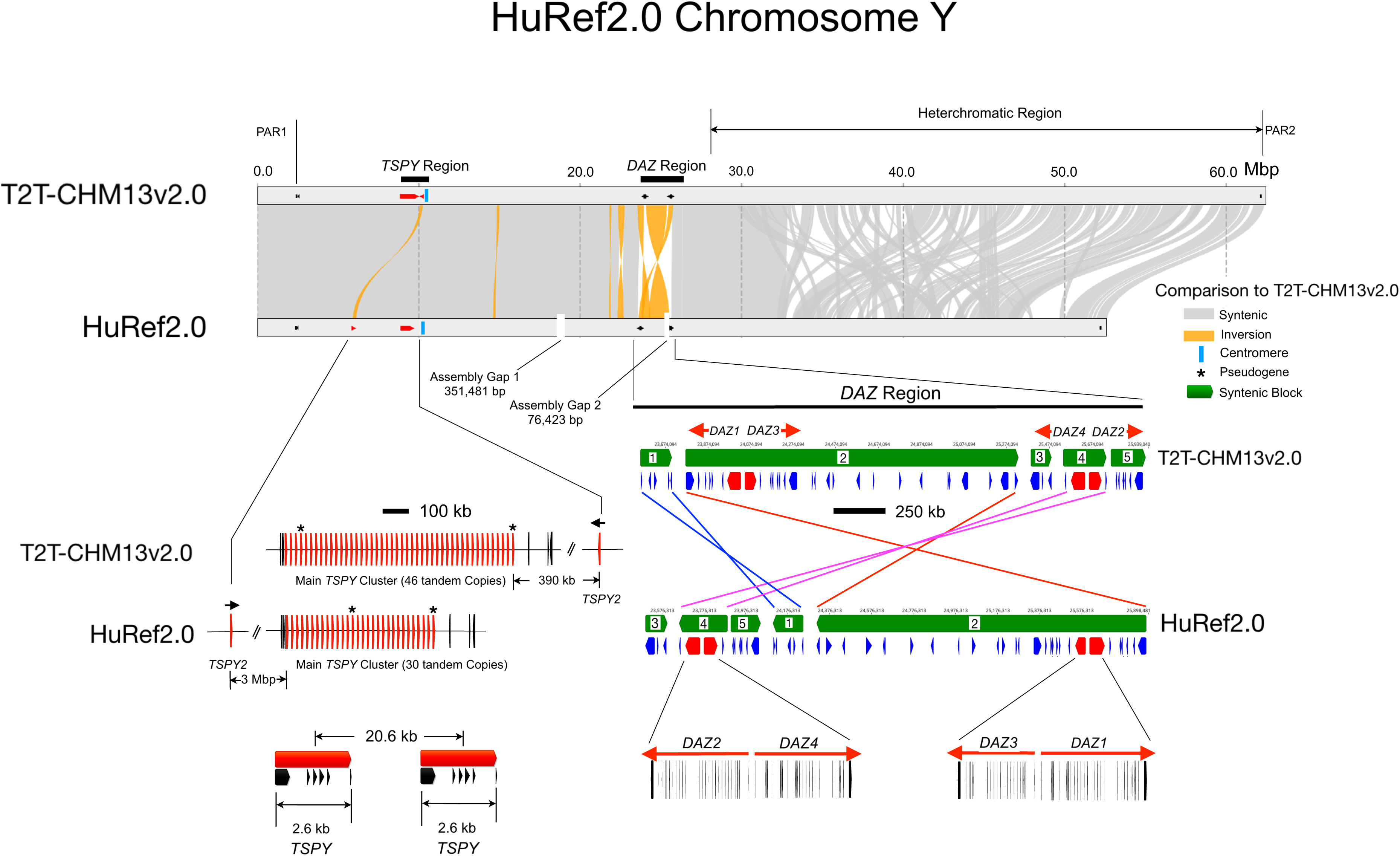
HuRef2.0 ChrY Comparison with T2T-CHM13v2.0. The position and size of the two assembly gaps prior to filling are shown. Orange ribbons show inverted or translocated regions and grey ribbons show syntenic segments. The *TSPY* cluster is magnified, showing an inversion between HuRef2.0 and T2T-CHM13v2.0 and 16 fewer copies on HuRef2.0. The *DAZ* region also shows multiple inversions and translocations between the two assemblies. The other gene families annotated on ChrY are listed in SI-10.

The primary *TSPY* gene cluster in HuRef2.0 spanned 620 kb in the *MSY* (male-specific) region of ChrY, comprising 30 tandem copies of a 20.6 kb repeating unit, each encoding a 5-exon *TSPY* gene. *TSPY* varies between 30-60 copies in the general population, with T2T-CHM13v2.0 having 46 copies. Copy number variation in *TSPY* has been reported to influence spermatogenic efficiency and correlates positively with sperm count^47^. A flanking family member, *TSPY2*, which is located 390 kb distal to the main *TSPY* cluster in T2T-CHM13v2.0, is found in an inverted orientation 3 Mb proximal to the cluster in HuRef2.0. This configuration in HuRef2.0 is apparently polymorphic in the general population, with *TSPY2* located in a similar position in GRCh38.p14 as in HuRef2.0. The remaining ampliconic gene clusters (*BPY2, CDY, HSFY, PRY, RBMY, VCY, XKRY*) are present in HuRef2.0 (see SI-10 for coordinates).

### Macro-Satellite Sub-Telomeric Region on Chr4q35

We next examined our workflow’s handling of other tandem repeats. Figure 3a depicts the *DUX4*/*DUX4L* gene cluster in the macro-satellite sub-telomeric region on Chr4q35. *DUX4* is an essential gene expressed in development. In adults, *DUX4* expression is repressed by the methylation of an upstream tandem array of nearly identical 3.3 kb non-functional *DUX4-like* (*DUX4L*) gene, which is missing a regulatory (exon-3) required for function. The loss of methylation from the genomic contraction of the *DUX4L* repeating units to below ∼10-20 copies de-repress *DUX4* expression in adults, giving rise to facioscapulohumeral muscular dystrophy^48^. T2T-CHM13v2.0^9^ has 33 tandem copies of *DUX4L*, and T2T-YAO^11^ has 28 tandem copies. Interestingly, the other two Chinese T2T submissions, T2T-CN1mat and T2T-CN1pat^10^, have 14 and 11 copies of *DUX4L,* respectivel*y*, but both haplotypes in T2T-CN1 are missing a functional *DUX4,* suggestive of a mis-assembly at the distal ends of the clusters.

**Figure 3.**
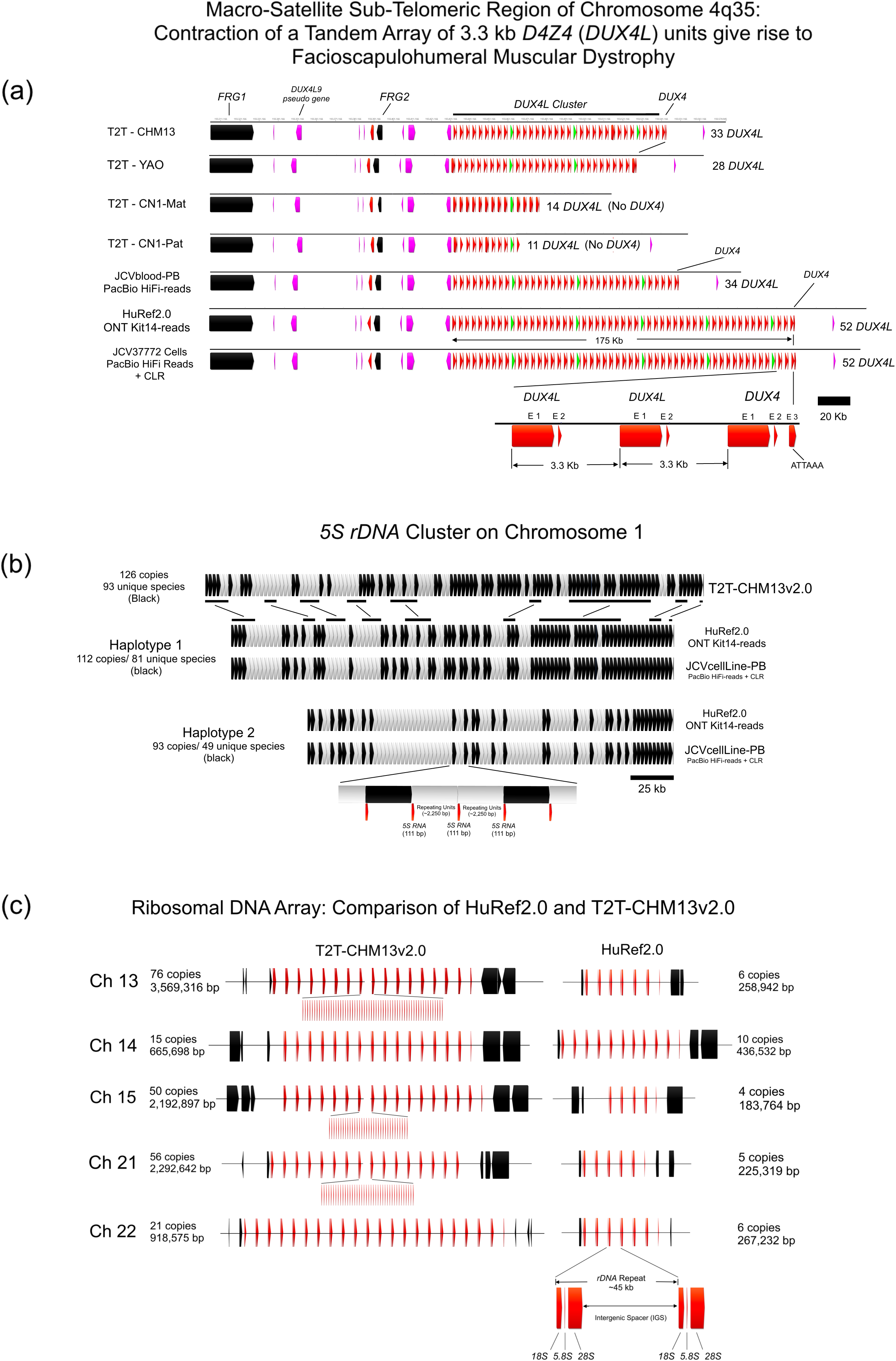
HuRef2.0 Macro-Satellite Sub-Telomeric Region of 4q35 and Ribosomal DNA Gene Clusters. (a) Macro-Satellite Sub-Telomeric Region of 4q35 in Different Near T2T-Assemblies. Contraction of the *D4Z4* repeat array gives rise to Facioscapulohumeral Muscular Dystrophy. Flanking genes are in black. Pseudogenes are in purple. *DUXL* and *DUX4* genes are in red. Both HuRef2.0 (assembled using ONT-reads from JCV blood DNA) and JCVcellLine-PB (assembled using PacBio HiFi and PacBio CLR from JCV37772 cell DNA) assembled 52 copies of *DUX4L*, supporting the accuracy of the assemblies using an orthologous sequencing platform and a different DNA source. JCVblood-PB (assembled from PacBio HiFi-reads alone from JCV blood DNA) only assembled 34 copies, revealing a limitation of the use of PacBio HiFi reads alone in assembling low complexity tandem repeats. Both T2T-CN1 haplotypes have no functional *DUX4*, indicating a possible mis-assembly of the distal end of the cluster. (b) Haplotype construction of *5S* rDNA cluster in HuRef2.0 and JCVcellLine-PB compared to T2T-CHM13v2.0. Unique copies within the cluster are highlighted in block. Duplicated copies are highlighted in grey, regions of homology are denoted by horizontal black bars. (Sequences can be downloaded at https://doi.org/10.5281/zenodo.19861724). (c) *45S* rDNA Arrays between HuRef2.0 and T2T-CHM13v2.0. The number of *45S* rDNA repeating units in HuRef2.0 is fewer than T2T-CHM13v2.0 across all five chromosomes.

In HuRef2.0, the *DUX4*/*DUX4L* gene cluster is complete and fully phased. The cluster resides on a 2.8 Mb phase block, with 52 and 28 tandem copies of *DUX4L* for haplotype 1 and haplotype 2, respectively. Identical complete and fully phased *DUX4*/*DUX4L* gene clusters were assembled from JCV37772 cells using PacBio HiFi-reads supplemented with CLRs, supporting the accuracy of the assemblies using an orthologous sequencing platform and a different DNA source. By contrast, an assembly using PacBio HiFi-reads alone (without the benefit of CLR for scaffolding) produced a cluster comprising only 34 *DUX4L* repeating units. We observed a similar finding for the *45S* ribosomal gene cluster, where reads are not long enough to span or resolve tandem repeat regions of low complexity, resulting in a deletion of internal repeating units during assembly (see Figure 3c and SI-11).

### 5S Ribosomal Gene Cluster

In T2T-CHM13v2.0, the *5S* rDNA cluster on Chr1 comprised a tandem array of 126 2.25 kb repeating units that shared ∼99% identity, of which 93 units are unique (indicated in black in Figure 3b). HuRef2.0 achieved complete and fully phased assemblies of the *5S* cluster, comprising tandem arrays of 112 and 93 repeating units for haplotype 1 and haplotype 2, respectively. The identical complete and phased clusters were also assembled from JCV37772 cells using PacBio HiFi-reads supplemented with PacBio CLR, supporting the accuracy of the assemblies and phasing using an orthologous sequencing platform and DNA source.

### 45S Ribosomal Gene Clusters

The *p*-arms of the five acrocentric chromosomes (Chr13, 14, 15, 21, and 22) are amongst the most difficult regions of the genome to assemble due to the preponderance of repetitive sequences^19,20,21,22,23,24^. The *45S* rRNA gene clusters in the *p*-arms posed a unique challenge due to their low sequence complexity and the extraordinary number and length of the tandem repeating units^21,22,23^. In contrast to the 15 to 76 tandem copies of the *45S* RNA precursor gene across the acrocentric chromosomes in T2T-CHM12v2.0^9^, the number of assembled *45S* ribosomal precursors in HuRef2.0 is much lower, comprising 6 to 10 copies per chromosome (Figure 3c). Without mapping information, we cannot independently confirm the reduced size of the *45S* rRNA gene clusters in HuRef2.0. However, we believe the *45S* rRNA clusters in HuRef2.0 might represent a limitation of our assembly workflow using only ONT Simplex reads. Here, the basic repeating unit is a ∼45 kb low-complexity segment comprising a *45S* RNA precursor, which is processed into mature *18S*, *5.8S* and *28S* rRNAs, and an intergenic spacer. The combination of low-sequence diversity together with a repeating unit that is significantly longer than the average length of the ONT-reads (read-length N50 value of 26.9 kb), the unambiguous growth of contigs cannot be supported. Under such conditions, the hifiasm assembler defaults to merge the longest high-confidence contigs on either side of the cluster, resulting in an assembled cluster that is deleted of internal units. Here, a case could be made for ultra-long-reads to span multiple identical repeated units to facilitate assembly.

HuRef2.0 is not unique in having problems with the *45S* ribosomal cluster. The *45S* clusters in T2T-CHM13v2.0^9^ were heavily patched using nine identical sequences guided by optical mapping or other long-range information, rather than being true *de novo* assemblies (SI-11a). The other recent T2T-assemblies appeared to have similar problems (SI-11c).

### Variant Calling

Figure 4 shows the intersection sets of JCV SNVs/indels and SVs called against GRCh38.p14 using Illumina short-read mapping, ONT or PacBio long-read mapping, and by direct assembly to assembly comparison (see Materials and Methods). The number of, and overlapping in, variants reflect the current inherent strength and bias of each method^49^. Of the 382 rare OMIM phenotype-associated SNV/indel coding variants identified (Figure 4a; SI-12): 131 were identified by all four methods, 15 were identified using only Illumina short-reads, 7 were identified using only ONT-reads, 8 were identified using only PacBio HiFi-reads, and 3 were identified using only an assembly-to-assembly comparison.

**Figure 4.**
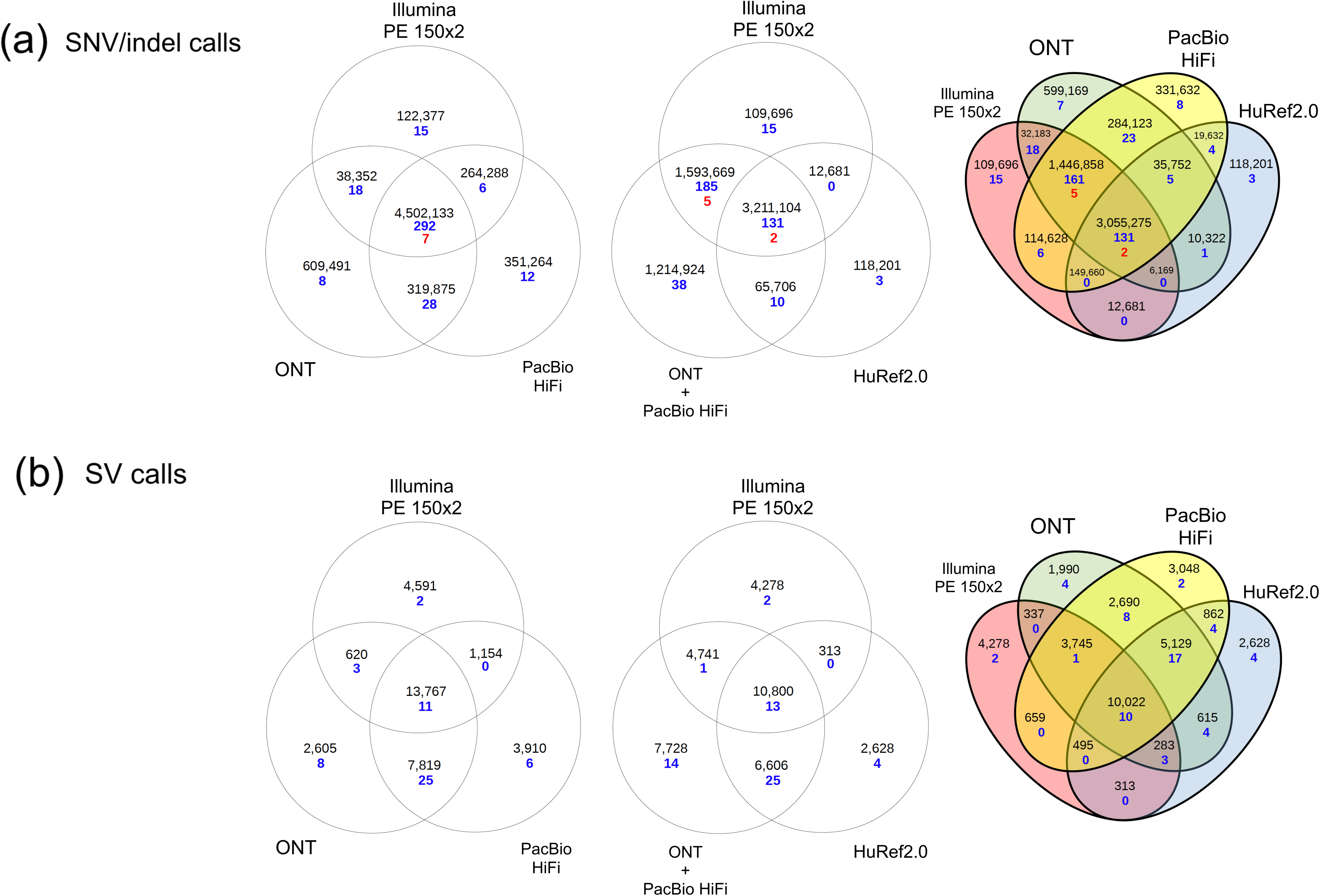
Variant Calling and Medical Annotation of JCV. (a) JCV SNVs and indels called against GRCh38 by short reads (117x Illumina PE 150×2) using DRAGEN^56^, ONT-reads (103x) using wf-human (https://github.com/epi2me-labs/wf-human-variation), PacBio HiFi-reads (101x) using DeepVariant^38^, and direct comparison of HuRef2.0 to GRCh38 using paftools^55^. Blue numbers represent rare (<3%) OMIM phenotype-associated variants found in coding regions (SI-12). Red numbers represent medically relevant sequence variants reported in ClinVar^60^ or represented a loss of function (SL-13). (b) JCV SV called by short reads (117x Illumina PE 150×2) using DRAGEN^56^, ONT-reads (103x) using SVIM^57^, PacBio HiFi-reads (101x) using pbsv (https://github.com/PacificBiosciences/pbsv), and direct comparison of HuRef2.0 to GRCh38 using SVIM-asm^57^. Blue numbers represent rare (<1%) variants overlapping OMIM phenotype-associated genes (https://www.omim.org), the full list of which can be found SI-14.

Based on current annotation, the donor carries seven medically relevant variants, as defined as those classified as pathogenic or likely pathogenic by ACMG guidelines^50^ (Figure 5a, SI-13), most of which were heterozygous variants associated with recessive conditions. All seven medically relevant variants identified were called by both long- and short-reads, but five of these, in the *COL4A4, BTD, ACOX2, GPR179,* and the *RAD51C* genes were not called by assembly-to-assembly comparison. The haplotypes with the aforementioned variants were not present in our haploid assembly. This suggests that future implementation of assembly-to-assembly calls should be performed with fully phased diploid assemblies and references.

**Figure 5.**
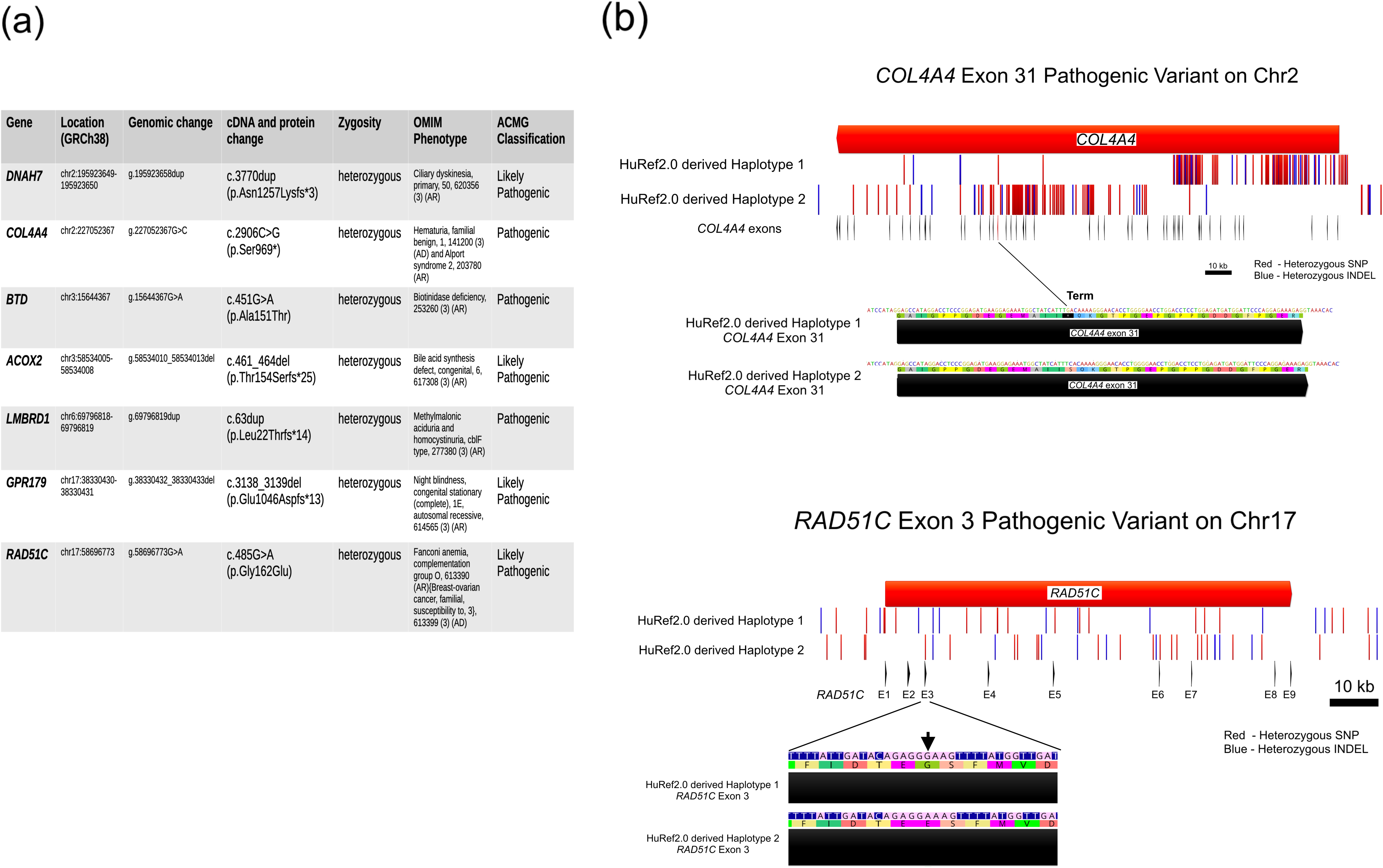
Variants that are Classified as Pathogenic or likely Pathogenic by ACMG Guidelines^61^. (a) The Seven Variants in HuRef2.0 that are classified as Pathogenic or likely Pathogenic by ACMG Guidelines^61^ (SI-13). (b) Haplotype-Resolved Regions of *COL4A4* and *RAD51C* in JCV, both containing variants of particular relevance. There is a heterozygous nonsense mutation in exon 31 of *COL4A4* (haplotype 1), and a heterozygous missense mutation (Gly to Glu) at exon 3 of *RAD51C* (haplotype 2). SNPs and indels against GRCh38 are highlighted in red and blue, respectively. (Sequences can be downloaded at https://doi.org/10.5281/zenodo.19861724).

Two variants are of particular relevance to the donor. A heterozygous missense variant p.(Gly162Glu) in *RAD51C* had been previously described in the literature in individuals with breast or/and ovarian cancer (Figure 5b). Importantly, two independent studies had found this variant to have a deleterious impact on protein function in various cellular assays, including deficient homologous recombination, impaired interactions with *RAD51* paralogs, and deleterious responses to cisplatin and olaparib treatment^51,52^, suggesting the variant to be likely pathogenic for ovarian and breast cancer susceptibility. Genetic findings often have implications beyond an individual, and the donor previously reported a family history of ovarian cancer^8^.

The second variant of potential interest is a heterozygous stop-gain variant p.(Ser969*) in *COL4A4*, a gene associated with autosomal recessive Alport syndrome^53^ (Figure 5b). While heterozygous carriers are not at risk of renal disease, the presence of a monoallelic pathogenic variant can manifest as benign hematuria, which can be misinterpreted as a symptom of urinary infection or malignancy. The analysis also indicated potential carrier status for other recessive disorders (SI-12, SI-13, SI-14).

Figure 4b shows the intersection sets of JCV SV calls (variants greater or equal to 50 bp) among the four methods. Although we identified no clinically relevant SVs, we identified 59 rare variants overlapping OMIM phenotype-associated genes (SI-14). Of these, 10 were called by the four methods; 2, 4, 2, and 4 calls were unique to Illumina short-reads, ONT long-reads, PacBio HiFi-reads, and assembly-to-assembly comparison, respectively.

### A Foundation for Scaling

We are currently making incremental improvements to further reduce time and cost associated with performing high-quality assemblies. First, we are optimizing assembly from single ONT flowcells (∼35x) to reduce operational cost and speed the workflow for eventual deployment. Within sampling variations, assembly from one ONT flowcell have so far produced useful human assemblies having between 3 and 13 complete chromosomes, NG50 values between 110 to 133 Mb, contig counts between 86 and 163 contigs, and ∼3,500 phase blocks (SI-15). In parallel, we sought to improve ONT read-lengths. By simply eliminating the DNA shearing step in ONT library construction, we have improved read-length N50 values by more than 5-8 kb, resulting in measurable improvements in assembly performance at lower coverage. Finally, as an alternative to the present cumbersome ultra-long read protocols that require specially prepared DNA, we are transitioning from ONT Simplex reads to a simplified and scalable stand-alone hexamminecobalt(III) chloride-based long-read protocol^54^ with only minor adjustments to the existing standard DNA purification protocol. Specifically, we aimed to improve phasing using the improved long-reads. Based on the roadmap provided by HuRef2.0, 1,908 of the 2,384 switch-over regions are less than 100 kb in length, to which we believe could be resolved with the new improved long-reads (SI-16). Resolution of these switch-over regions could reduce the number of phase blocks from 2,393 to 485, with a concomitant increase in phase block NG50 value from 2.36 to ∼17 Mb.

## Material and Methods

### Donor History and DNA Purification

JCV DNA for genome assembly was purified from 5 ml fresh blood or from low passage (< 5 passages) JCV37772 cells, an EBV-transformed lymphoblastic cell line developed from the same donor, using Qiagen MagAttract HMW DNA kit (Qiagen, Germantown, MD). Purified DNA from fresh blood (NCBI: BioSample SAMN48688833) and from JCV37772 cells (NCBI: BioSample SAMN48687824) have bulk peak lengths of 60 kb on an Agilent Tape Station (Agilent Technologies).

### Library Construction and DNA Sequencing

Library construction and sequencing were performed according to vendor’s instructions. DNA was quantified by Qubit DNA Assay (ThermoFisher, Waltham, MA). ONT library construction was performed in accordance with protocol SQK-LSK114, incorporating a light DNA shearing step with g-tubes (Covaris, Woburn, MA). Sequencing was carried out using the R10.4.1 flowcell on the PromethION24 sequencer (96 hr run). HAC or SUP base calls were made using Dorado (https://github.com/nanoporetech/dorado). The PacBio HiFi library was constructed using SMRTbell prep kit 3.0, incorporating the shearing step outlined in Appendix A4. Sequencing was carried out on the PacBio Revio (24hr run).

### ONT Base Calling

High accuracy basecalling (HAC) was performed on the PromethION machine with Dorado basecall server v7.4.14 using model dna_r10.4.1_e8.2_400bps_hac@v4.3.0. Super accuracy basecalling (SUP) was done with standalone dorado v0.9.0 using model <u>dna_r10.4.1_e8.2_400bps_sup@v5.0.0</u>.

### Sub-Assembly Parameters

Hifiasm-0.24.0-r702 was used for genome assembly. The -r parameter instructs the program how many rounds of correction to perform. Zero is not allowed as a valid input. To effectively run zero rounds of the correction step, we commented out lines 5576-5591 and lines 5596-5597 from Correct.cpp and line 2111 and line 2114 from ecovlp.cpp. The bw parameter is not an official hifiasm parameter intended to be changed by the user. Its relationship to the expected error rate of ONT was established merely based on our understanding of the code. Its value was adjusted by changing the first occurrence of 0.05 to 0.02 on line 3231 of ecovlp.cpp.

#### HuRef2.0 ChrY Gap Filling

The main contigs of ChrY were selected from sub-assembly H, consisting of three contigs totaling 52,137,524bp, reaching telomeres on both p and q arms with two gaps in the middle. The first gap was estimated to be 351 kb wide based on alignment with T2T-CHM13v2. A short contig (493,661 bp) from sub-assembly K was found to reside within the gap and was used to shorten the gap to around 157 kb wide. The sequences on either side of the gap were then extended into the gap by aligning reads to the edge sequences and constructing a consensus by local assembly. The process was repeated until the gap was filled. The second gap on ChrY was estimated to be around 82 kb based on alignment with T2T-CHM13v2.0 and was filled by read alignment and local assembly as described for the first gap.

### Assembly Polishing and Validation

To correct for any assembly errors, ONT SUP reads used for assembling the genome were aligned back to the assembly with minimap2 v2.28^55^ using the “-x lr:hqae” parameter. Two programs were used to detect sites in the assembly that is different to both alleles as revealed by the read alignment. DeepVariant v1.8.0^38^ was run with model type set as ONT_R104. Results were filtered to include only those with coverage between 10X to 200X and read ratio of variant to reference larger than 0.8. Sites coinciding with homopolymer tracks and simple repeats were excluded. Sniffles v2.6.2^39^ with default parameters was also used and only results with call quality greater than 30 were selected. Filtered results from both sources are all manually inspected to ensure reads are properly aligned and reveal true assembly error.

### Post-Assembly Correction by GATK

To search for sequencing errors not corrected during the hifiasm pre-assembly stage, we aligned 107.7X coverage of Illumina reads onto the assembly using bwa-mem (v0.7.17) (https://doi.org/10.48550/arXiv.1303.3997) and called SNV and indels using GATK (v4.6.1.0) HaplotypeCaller^28^ with default parameters. To prevent over-correction of unique bases in the repeated portion of the genome, only regions with mapping quality ≥ 50 and 50-150× alignment depth were considered. Sex chromosomes were excluded in this process as their repeat structure and lower coverage makes them less suitable to be corrected by short-reads. We corrected only the events with quality greater than 200 as reported by GATK and homopolymer tracks less than 30bp long. To evaluate the quality of the final assembly, a reference-independent k-mer analysis was performed using Merqury^29^ with 107.7X coverage of Illumina reads. A k-mer size of 21 was chosen as recommended by the program to reduce the collision rate to 0.1% given the genome size.

### Phasing

The ONT SUP-reads used for assembling the genome were aligned back onto HuRef2 with minimap2 v2.28^55^ using the “-x lr:hqae” parameter specific for accurate long-reads with Q20 or above. DeepVariant v1.8.0^38^ was used for call SNV and indels from the alignment while Sniffles2 v2.6.2^39^ was used to call structural variants. The union of all the variants was used as input to phase the genome with WhatsHap v2.4^40^. Two or more contiguous phased variants form a phase block, within which the allele of each variant can be assigned their respective haplotype with respect to each other. The gap between phase blocks represents switch over regions, where either the haplotype difference is too great for read alignment, or the sparsity of heterozygous variants eluded the spanning capability of the currently available reads.

### Single Nucleotide Variant and Indel Detection

Illumina short-read (PE150×2) alignment and variant calling was done using DRAGEN Germline Whole Genome 4.2.4 with default parameters^56^. For ONT-reads, the wf-human-variation (EPI2ME Labs, v2.1.0-gb82338d, https://github.com/epi2me-labs/wf-human-variation) workflow developed by ONT was used. The “--bam_min_coverage 20” option was applied to evaluate regions with 20X or higher coverage. Variants for which the reference or alternative base was “N” were discarded. For assembly-to-assembly comparison, we first aligned contigs from HuRef2.0 to GRCh38 using minimap v2.24^55^ with the “-x asm5” option (representing an expected sequence divergence of 0.1%), then called SNVs/indels using the paftools.js script from minimap2. Only regions with high mapping quality (MAPQ=60), single-contig coverage, and correct chromosome match were considered. Entries with the same chromosome and start position were merged as they represent a single event of short sequence substitution, instead of two separate insertion and deletion. For all three detection methods, we removed mitochondrial variants, variants larger than 50 bp, and variants on GRCh38 alternate contigs/haplotypes.

### Structural Variant Detection

SVs were detected using the same three approaches as detecting SNVs and indels, but with different bioinformatics tools. For Illumina, alignment and variant calling were both performed using DRAGEN Germline Whole Genome 4.2.4 with default parameters^56^. Variants with flags: “LowPopulationVariance”, “NoPairSupport”, “MaxDepth”, “MinQUAL”, “MaxMQ0Frac”, and “It50bp” were removed. For ONT-reads, the reads were aligned using minimap v2.11^53^ and variants were detected using SVIM^57^, both with default parameters. For assembly-to-assembly comparison, the contigs from HuRef2.0 were aligned to GRCh38 using minimap v2.11 with the “-x asm5” option, and variants were detected using SVIM-asm^55^ with the “haploid” parameter. For both SVIM^57^ and SVIM-asm^57^, we removed mitochondrial variants, variants smaller than 50 bp, and variants on GRCh38 alternate contigs/haplotypes. Variants with the same chromosome and start position but different alternate alleles were collapsed (i.e., considered to be a single variant). Only variants where the FILTER column presented a “PASS” was included. For SVIM^57^, we removed variants for which the QUAL value was less than 5.

### Variant Overlap Determination

The overlap among the SNVs/indels detected from the different methods was determined by merging the VCF files using bcftool merge^58^. Determining the overlap among the SV calls from the three SV-detection methods was more challenging, as it is not always clear whether calls with overlapping coordinates represent the same event. To address this problem, we utilized a window-based approach. Specifically, we divided GRCh38 (canonical chromosomes only) into 1 kb windows, and for each window, we determined the method(s) that reported a structural variant whose coordinates overlapped that window. Thus, SV call overlap is reported at the level of windows rather than SV calls.

### Variant Annotation

The SNVs/indels called from the three different methods were annotated with Ensembl Variant Effect Predictor^59^ (VEP) (Ensembl release 114) with reference to GRCh38.p14. We filtered for rare variants (<3% allele frequency) that intersect the coding region of genes with an associated Online Mendelian Inheritance in Man (OMIM) morbid map phenotype (https://www.omim.org). From the filtered list, we highlighted variants that were categorized as pathogenic in ClinVar^60^ or that cause gene loss of function (disrupting a canonical splice site or causing a stop gain or frameshift). These variants were then interpreted and classified according to American College of Medical Genetics and Genomics (ACMG) guidelines^61^. The SVs were annotated with TCAG CNV/SV Annotation Pipeline (v1.7.5), an internally developed R script (v4.4.0) that utilizes the GenomicRanges and data.table libraries to incorporate gene annotations, genomic features, phenotype ontologies and disease information from different online sources^36^. The annotated SVs were filtered for rarity (<1% in the population) and overlapping coding exons in genes with an OMIM morbid map phenotype.

## Data Availability

All sequencing data used in the present study are deposited under BioProjects: PRJNA1266749, PRJNA12266743, and PRJNA1268575. Assemblies HuRef2.0, JCVblood-PB, JCVcellLine-PB are deposited at NCBI Genbank under the accession JBPPLP01, JBTWEK01, and JBPQOS01, respectively. Phased heterozygous sites and associated phase blocks of HuRef2.0 (with HuRef2.0 coordinates) can be downloaded from https://doi.org/10.5281/zenodo.19861724, together with the haplotype constructions of the extended *MHC* region*, 22q11*.2, *5S rDNA* cluster, *COL4A4* and *RAD51C*.

## Author Contributions

SL, JCV, and SWS initiated the project. SL designed and directed the project, analysed results, performed genome annotation, and wrote the manuscript. TNHL performed and validated genome assembly, and analysed results. AHYT contributed to experimental design, analysis and genome annotation. TP contributed to the analysis of mitochondrial DNA. LL, TNHL, CRM, JRM, MSR, BT, and Brett T contributed to variant calling and their annotation. All authors contributed to writing and editing the manuscript.

## Acknowledgements

The work represents an ongoing collaboration between the J Craig Venter Institute and The Centre for Applied Genomics (TCAG) at the Hospital for Sick Children in Toronto. The authors wish to thank: Sachin Desai, Lan He, Karen Ho, and Sanjeev Pullenayegum for expert Illumina, PacBio and ONT sequencing; Amirhossein Hajianpour, Omar Hamdan, Edward J Higginbotham, and Joseph Whitney for technical and informatics support; Giovanna Pellecchia and Thomas Nalpathamkalam for authoring and maintaining the TCAG CNV/SV annotation pipeline; Jingle Candelario-MacDonald, Kulay Janneh, Miranda Lorenti, and Marnita Manalo in the TCAG Biobanking group for DNA purification and the development of the JCV37772 cell line and; Richard F Wintle for many helpful discussions. DNA sequencing was performed by The Centre for Applied Genomics, The Hospital for Sick Children, Toronto, Canada. Funding support is from the Canada Foundation for Innovation to the CGEn National Genome Sequencing and Informatics Centre, the Lau Family Endowment in Genome Sciences, the University of Toronto McLaughlin Centre, and Genome Canada. SWS holds the Northbridge Chair in Paediatric Research at the Hospital for Sick Children and the University of Toronto.

## Ethics Statement

Ethical approval was obtained from the Hospital for Sick Children Ethics Board (Approval No: 1000053640). All procedures involving human participants were conducted in accordance with institutional guidelines. Written informed consent was obtained from the participant prior to his inclusion in the study.

## References

1. E. S. Lander, Initial impact of the sequencing of the human genome. Nature 470, 187–197 (2011).

2. L. Hood, L. Rowen, The Human Genome Project: big science transforms biology and medicine. Genome Med 5, 79 (2013).

3. E. D. Green, J. D. Watson, F. S. Collins, Twenty-five years of big biology. Nature 526, 29–31 (2015).

4. S. Trip, M. Grueber, Economic impact of the human genome project. Battelle Memorial Institute (2011).

5. The Human Genome Sequencing Consortium, Initial sequencing and analysis of the human genome. Nature 409, 860-921 (2001).

6. J. C. Venter, et al., The sequence of the Human Genome. Science 291, 1304–1351 (2001).

7. The Human Genome Sequencing Consortium, Finishing the euchromatic sequence of the human genome. Nature 431, 931-945 (2004).

8. S. Levy, et al., The diploid genome sequence of an individual human. PLoS Biol 5, 2113–2144 (2007).

9. S. Turk, et al., The complete sequence of a human genome. Science 376, 44–53 (2022).

10. C. Yang, et al., The complete and fully phased diploid genome of a male Han Chinese. Cell Res 33, 745–761 (2023).

11. Y. He, et al., T2T-TAO: A telomere-to-telomere assembled diploid reference genome for Han Chinese. Genomics Proteomic Bioinformatics 21, 1085–1100 (2023).

12. N. F. Hansen, et al., A complete diploid human genome benchmark for personalized genome. BioRxiv (10.1101/2025.09.21.677443) (2025).

13. H. Cheng, G. T. Concepcion, X. Feng, H. Zhang, H. Li, Haplotype-resolved *de novo* assembly using phased assembly graphs with hifiasm. Nat Meth 18, 170–175 (2021).

14. H. Cheng, et al., Haplotype-resolved assembly of diploid genomes without parental data. Nat Biotech 40, 1332–1335 (2022).

15. H. Cheng, et al., Efficient near telomere-to-telomere assembly of Nanopore Simplex reads. Nature (10.1038/s41586-026-10105-6) (2026).

16. A. M. Wenger, et al., Accurate circular consensus long-read sequencing improves variant detection and assembly of a human genome. Nat Biotechnol 37, 1155–1162 (2019).

17. D. J. Taylor, et al., Beyond the human genome project: The age of complete human genome sequences and pangenome reference. Ann Rev Genom Hum Genet 25, 77–104 (2024).

18. H. Zhang, C. Jain, S. Aluru, A comprehensive evaluation of long-read error correction methods. BMC Genomics 21 (Suppl 6), 889 (2020).

19. T. J. Treangen, S. L. Salzberg, Repetitive DNA and next-generation sequencing: Computational challenges and solutions. Nat Rev Genet 13, 36–46 (2012).

20. T. Liehr, Repetitive elements in humans. Int J Mol Sci 22, 2072 (2021).

21. Y. Hori, A. Shimamoto, T. Kobayashi, The human ribosomal DNA array is composed of highly homogenized tandem clusters. Genome Res 31, 1–12 (2021).

22. A. N. Hall, E. Motron, C. Queitsch, First discovered, long out of sight, finally visible: Ribosomal DNA. Trends Genet 38, 587–597 (2022).

23. S. Agrawal, A. R. D. Ganley, The conservation landscape of the human ribosomal RNA gene repeats. PLoS ONE 13, e0207531 (2018).

24. S. Gao, et al., A global view of human centromere variation and evolution. BioRxiv (10.64898/2025.12.09.693231) (2025).

25. F. Nie, et al., *De novo* diploid genome assembly using noisy long reads. Nat Comm 15, 2964 (2024).

26. D. Stanojević, D. Lin, S. Nurk, P. F. de Sessions, M. Šikić, Telomere-to-telomere phased genome assembly using HERRO-corrected Simplex Nanopore reads. BioRxiv (10.1101/2024.05.18.594796) (2024).

27. Y. Liu, et al., Repeat and haplotype aware error-correction in nanopore sequencing reads with DeChat. Comm Biol 7, 1678 (2024).

28. G. A. V. de A. O’Connor, D. Brian, Genomics in the cloud: Using Docker, GATK, and WDL in Terra (1 ed). O’Reilly Media (2020).

29. A. Rhie, B. P. Walenz, S. Koren, A. M. Philippy, Merqury: Reference-free quality, completeness, and phasing assessment for genome assemblies. Genome Biol 21, 245 (2020).

30. M. Krzywinski, et al., Circos: An information aesthetic for comparative genomics. Genome Res 19, 1639–1645 (2009).

31. M. T. Lott, et al., mtDNA variation and analysis using MITOMAP and MITOMASTER. Curr Protoc Bioinform 1, 1.23.1–1.23.26 (2013).

32. N. Isern, F. Joaquim, V. L. de Rioja, The ancient cline of haplogroup K implies that the Neolithic transition in Europe was mainly demic. Sci Rep 7, 11229 (2017).

33. X. Chang, et al., Mitochondrial DNA haplogroups and risk of attention deficit and hyperactivity disorder in European Americans. Transl Psychiatry 10, 370 (2020).

34. X. Chang, et al., Mitochondrial DNA Haplogroup K is protective against autism spectrum disorder risk in populations of European ancestry. J Am Acad Child Adolesc Psychiatry 63, 835–844 (2024).

35. R. E. Frye, Editorial: mitochondrial gene variations increase autism risk: Uncovering the complex polygenetic landscape of autism. J Am Acad Child Adolesc Psychiatry 63, 775-777 (2024).

36. B. Trost, et al., Genomic architecture of autism from comprehensive whole-genome sequence annotation. Cell 185, 4409–4427 (2022).

37. D. Chalkia, et al., Association between mitochondrial DNA haplogroup variation and autism spectrum disorder. JAMA Psychiatry 74, 1161–1168 (2017).

38. R. Poplin, et al., A universal SNP and small-indel variant caller using deep neural networks. Nat Biotechnol 36, 983–987 (2018).

39. M. Smolka, et al., Detection of mosaic and population-level structural variants with sniffles2. Nat Biotechnol 42, 1571–1580 (2024).

40 M. Martin, et al., WhatsHap: Fast and accurate read-based phasing. 10.1101/085050 (2016).

41. R. Horton, et al., Gene map of the extended human MHC. Nat Rev Genet 5, 890–899 (2004).

42. D. M. McDonald-McGinn, et al., 22q11.2 deletion syndrome. Nat Rev Dis Primers 1, 10.1038/nrdp.2015.71 (2015).

43. Z. Motahari, S. A. Moody, T. M. Maynard, A.-S. LaMantia, In the line-up: deleted genes associated with DiGeorge/22q11.2 deletion syndrome: are they all suspects? J Neurodev Disord 11, 7 (2019).

44. Y. Ling, X. Kang, Y. Yi, S. Feng, G. Ma, H. Qu, *CLDN5*: From structure and regulation to roles in tumors and other diseases beyond CNS disorders. Pharmcol Res (10.1016/j.phrs.2024.107075 (2024).

45. A. Rhie, et al., The complete sequence of a human Y chromosome. Nature 621, 344–354 (2023).

46. P. Hallast, et al., Assembly of 43 human Y chromosomes reveals extensive complexity and variation. Nature 621, 355–364 (2023).

47. C. Krausz, C. Giachini, G. Forti, *TSPY* and male fertility. Gene 1, 308–316 (2010).

48. S. M. van de Maarel, R. Tawil, S. J. Tapscott, Facioscapulohumeral muscular dystrophy and *DUX4*: breaking the silence. Trends Mol Med 17, 252–258 (2011).

49. G. Gambardella, Joint processing of long- and short-read sequencing data with deep learning improves variant calling. Cell Reports Methods 5, 101107 (2025).

50. S. Richards, et al., Standards and guidelines for the interpretation of sequence variants: a joint consensus recommendation of the American College of Medical Genetics and Genomics and the Association for Molecular Pathology. Genet Med 17, 405–424 (2015).

51. C. Hu, et al., Functional and clinical characterization of variants of uncertain significance identifies a hotspot for inactivating missense variants in *RAD51C*. Cancer Res 83, 2557–25571 (2023).

52. R. Prakash, et al., Homologous recombination-deficient mutation cluster in tumor suppressor *RAD51C* identified by comprehensive analysis of cancer variants. Proc Natl Acad Sci (USA) 119, e2202727119 (2022).

53. T. S. T. Lim, et al., Pathogen variants in the Alport genes are prevalent in the Singapore multi-ethnic population with highest frequency in the Chinese. Sci Rep 15, 7691 (2025).

54. I. Cahyani, et al., An optimized toolkit for high-molecular-weight DNA extraction and ultra-long-read nanopore sequencing using glass beads and hexamminecobalt(III) chloride. Genome Res 35, 1154–1166 (2025).

55. H. Li, Minimap2: Pairwise alignment for nucleotide sequence. Bioinformatics 34, 3094–3100 (2018).

56. S. Behera, et al., Comprehensive genome analysis and variant detection at scale using DRAGEN. Nat Biotechnol 43, 1177–1191 (2025).

57. D. Heller, M. Vingron, SVIM: Structural variant identification using mapped long-reads. Bioinformatics 35, 2907–2915 (2019).

58. P. Danecek, et al., Twelve years of SAMtools and BCFtools. GigaScience 10, 1–4 (2021).

59. S.C. Dyer, et al., Ensembl 2025. Nuc Acids Res 53, D948–D957 (2025).

60. M. J. Landrum et al., ClinVar: public archive of relationships among sequence variation and human phenotype. Nuc Acids Res 42, (doi:10.1093/nar/gkt1113) (2014).

61. S. Richards, et al., Standards and guidelines for the interpretation of sequence variants: a joint consensus recommendation of the American College of Medical Genetics and Genomics and the Association for Molecular Pathology. Genet Med 17, 405–424 (2015).

