## Supplementary material for "The Revised Diploid Genome Sequence of an Individual Human: An Optimized Assembly Workflow for Scaling of near Telomere-to-Telomere Assemblies": SI_1.pdf

### SI-1a: Distribution of ONT Kit 14 Simplex-Reads from JCV Blood DNA

JCV Blood DNA

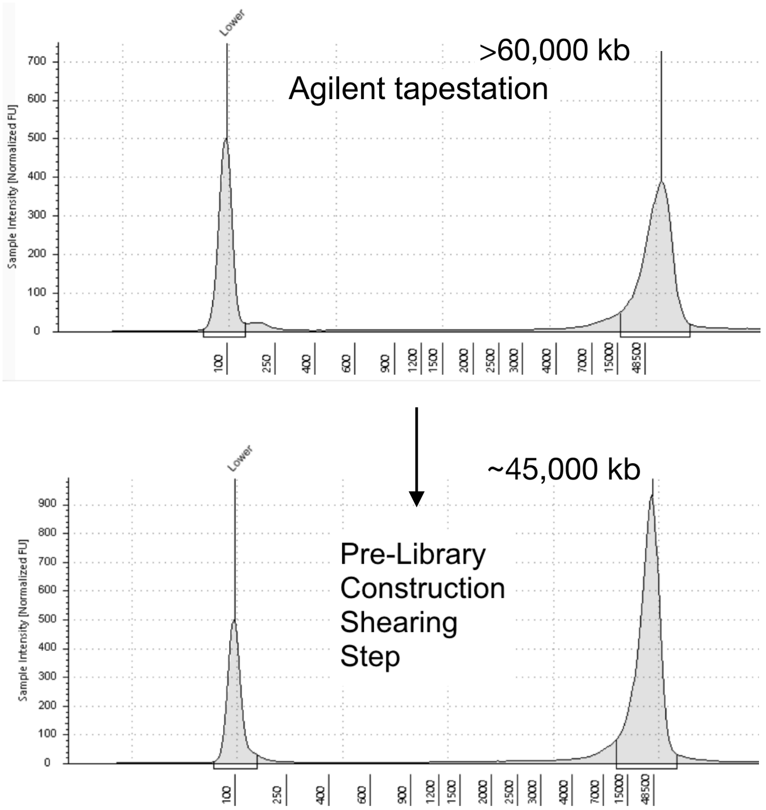


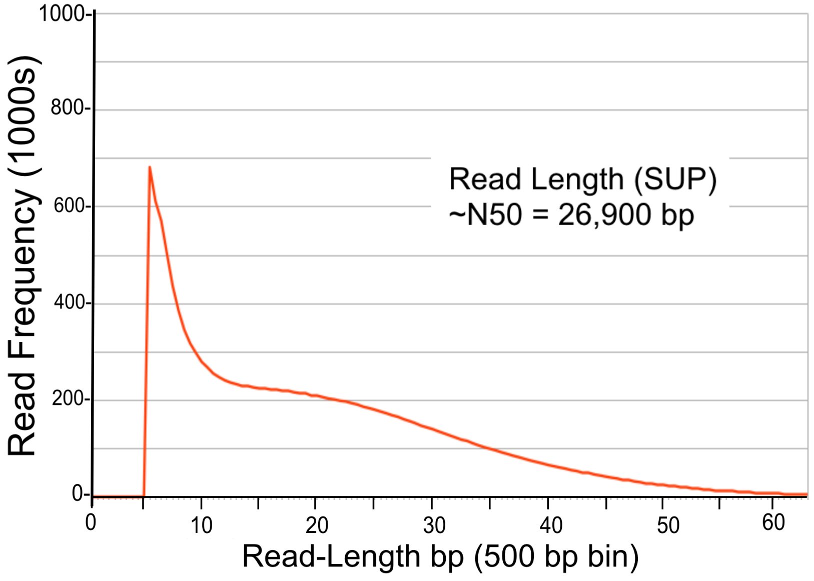


| Read-Length Category | | Super-Accuracy Basecalling (SUP): New Blood 20231027 R10 Kit14 ONT SUP (≥5kbp ≥Q9) | Coverage X (3.1 Gb) | Hi-Accuracy Basecalling (HAC): New Blood 20231027 R10 Kit14 ONT HAC (≥5kbp ≥Q9) | Coverage X (3.1 Gb) |
|---|---|---|---|---|---|
| Total | Mean Read Length (bp) | 20,219 | | 20,372 | |
| | Median Read Length | 17,427 | | 17,694 | |
| | Max Read Length | 1,174,432 | | 745,750 | |
| | Min Read Length | 5,000 | | 5,000 | |
| | **Total bases** | **319,649,797,669** | **103.113** | **308,600,830,182** | **99.549** |
| | Total Read Count | 15,809,317 | | 15,148,388 | |
| | **Read Length N50 (bp)** | **26,920** | | **26,951** | |
| | Read Length N90 | 10,118 | | 10,342 | |
| | Read Length L50 | 4,202,377 | | 4,073,363 | |
| | Read Length L90 | 11,312,872 | | 10,880,300 | |
| 2.5k (2k to <3k) | Read Count | 0 | | 0 | |
| | Bases | 0 | **0.000** | 0 | **0.000** |
| | **Read Length N50 (bp)** | 0 | | 0 | |
| 3.5k (3k to <4k) | Read Count | 0 | | 0 | |
| | Bases | 0 | **0.000** | 0 | **0.000** |
| | **Read Length N50** | 0 | | 0 | |
| 4.5k (4k to <5k) | Read Count | 0 | | 0 | |
| | Bases | 0 | **0.000** | 0 | **0.000** |
| | **Read Length N50** | 0 | | 0 | |
| 7.5k (5k to <10k) | Read Count | 4,431,632 | | 4,088,550 | |
| | Bases | 31,313,043,468 | **10.101** | 29,034,213,762 | **9.366** |
| | **Read Length N50** | **7,248** | | **7,335** | |
| 12.5k (10k to <15k) | Read Count | 2,395,385 | | 2,324,362 | |
| | Bases | 29,758,768,153 | **9.600** | 28,875,808,687 | **9.315** |
| | **Read Length N50** | **12,644** | | **12,643** | |
| 17.5k (15k to <20k) | Read Count | 2,174,790 | | 2,115,391 | |
| | Bases | 37,985,205,696 | **12.253** | 36,948,206,316 | **11.919** |
| | **Read Length N50** | **17,627** | | **17,628** | |
| 22.5k (20k to <25k) | Read Count | 1,945,833 | | 1,894,510 | |
| | Bases | 43,661,106,972 | **14.084** | 42,509,359,000 | **13.713** |
| | **Read Length N50** | **22,546** | | **22,546** | |
| 37.5k (25k to <50k) | Read Count | 4,421,780 | | 4,307,081 | |
| | Bases | 149,247,685,671 | **48.144** | 145,353,768,203 | **46.888** |
| | **Read Length N50** | **34,012** | | **34,006** | |
| 62.5k (50k to <75k) | Read Count | 380,663 | | 366,446 | |
| | Bases | 21,804,892,418 | **7.034** | 20,976,537,216 | **6.767** |
| | **Read Length N50** | **56,151** | | **56,107** | |
| 87.5k (75k to <100k) | Read Count | 44,066 | | 40,937 | |
| | Bases | 3,717,715,696 | **1.199** | 3,449,330,012 | **1.113** |
| | **Read Length N50** | **83,745** | | **83,604** | |
| 100k (>= 100k) | Read Count | 15,168 | | 11,111 | |
| | Bases | 2,161,379,595 | **0.697** | 1,453,606,986 | **0.469** |
| | **Read Length N50** | **130,768** | | **119,006** | |

### SI-1b: Distribution of PacBio HiFi-Reads from JCV Blood DNA

#### JCV Blood DNA

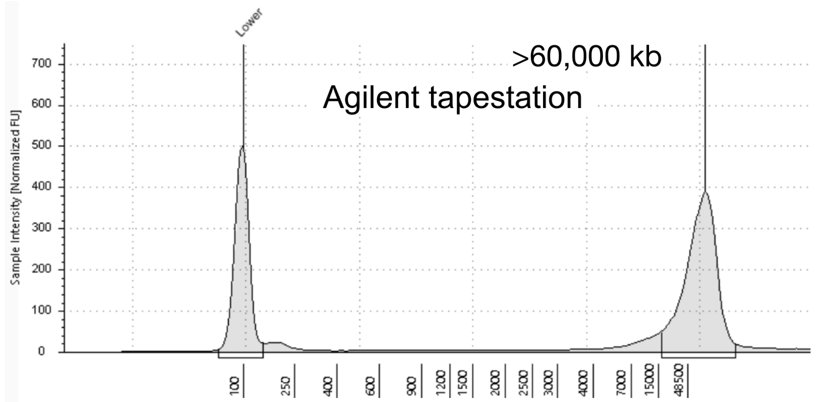


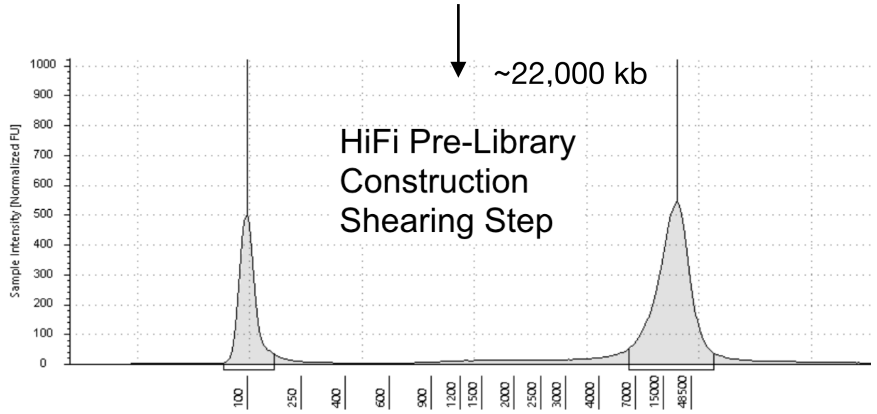


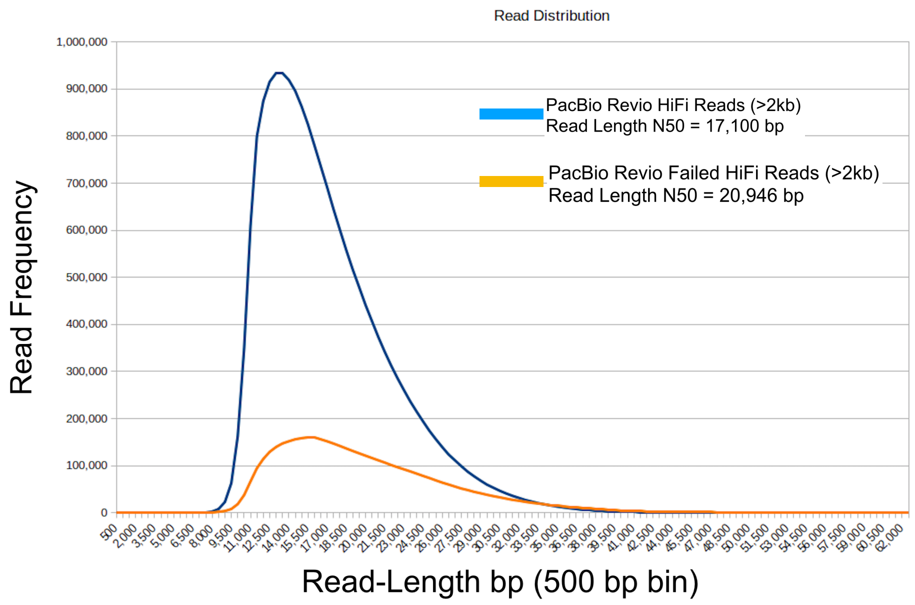


#### PacBio Revio HiFi-Reads

| | | HiFi-Reads | | Failed HiFi-Reads | |
|---|---|---|---|---|---|
| **Read-Length Category** | | **New Blood 20231027 PacBio Revio HiFi reads (≥ 2 kb)** | **Coverage X (3.1 Gb)** | **New Blood 20231027 PacBio Revio failed HiFi reads (≥2 kb)** | **Coverage X (3.1 Gb)** |
| Total | Mean Read Length (bp) | 16,694 | | 20,014 | |
| | Median Read Length | 15,618 | | 18,374 | |
| | Max Read Length | 65,378 | | 251,738 | |
| | Min Read Length | 2,001 | | 2,000 | |
| | **Total bases** | **312,794,990,694** | **100.902** | **90,029,878,770** | **29.042** |
| | Total Read Count | 18,737,091 | | 4,498,353 | |
| | **Read Length N50 (bp)** | **17,110** | | **20,946** | |
| | Read Length N90 | 11,972 | | 13,464 | |
| | Read Length L50 | 7,204,357 | | 1,621,573 | |
| | Read Length L90 | 15,894,470 | | 3,742,392 | |
| 2.5k (2k to <3k) | Read Count | 672 | | 815 | |
| | Bases | 1,696,919 | **0.001** | 2,059,955 | **0.001** |
| | **Read Length N50 (bp)** | **2,584** | | **2,577** | |
| 3.5k (3k to <4k) | Read Count | 607 | | 1,017 | |
| | Bases | 2,119,463 | **0.001** | 3,568,171 | **0.001** |
| | **Read Length N50** | **3,520** | | **3,546** | |
| 4.5k (4k to <5k) | Read Count | 667 | | 1,165 | |
| | Bases | 2,991,625 | **0.001** | 5,261,494 | **0.002** |
| | **Read Length N50** | **4,504** | | **4,536** | |
| 7.5k (5k to <10k) | Read Count | 260,011 | | 36,257 | |
| | Bases | 2,464,957,299 | **0.795** | 328,763,873 | **0.106** |
| | **Read Length N50** | **9,658** | | **9,552** | |
| 12.5k (10k to <15k) | Read Count | 8,092,406 | | 1,193,032 | |
| | Bases | 103,038,754,679 | **33.238** | 15,446,202,906 | **4.983** |
| | **Read Length N50** | **12,958** | | **13,228** | |
| 17.5k (15k to <20k) | Read Count | 6,272,595 | | 1,428,654 | |
| | Bases | 107,974,068,837 | **34.830** | 24,811,612,273 | **8.004** |
| | **Read Length N50** | **17,248** | | **17,474** | |
| 22.5k (20k to <25k) | Read Count | 2,801,261 | | 946,017 | |
| | Bases | 61,954,680,472 | **19.985** | 21,088,527,355 | **6.803** |
| | **Read Length N50** | **22,070** | | **22,331** | |
| 37.5k (25k to <50k) | Read Count | 1,308,792 | | 858,166 | |
| | Bases | 37,351,531,480 | **12.049** | 25,951,939,324 | **8.372** |
| | **Read Length N50** | **27,962** | | **29,631** | |
| 62.5k (50k to <75k) | Read Count | 80 | | 22,789 | |
| | Bases | 4,189,920 | **0.001** | 1,368,689,955 | **0.442** |
| | **Read Length N50** | **51,452** | | **60,167** | |
| 87.5k (75k to <100k) | Read Count | 0 | | 6,897 | |
| | Bases | 0 | **0.000** | 587,772,999 | **0.190** |
| | **Read Length N50** | **0** | | **85,002** | |
| 100k (>= 100k) | Read Count | 0 | | 3,544 | |
| | Bases | 0 | **0.000** | 435,480,465 | **0.140** |
| | **Read Length N50** | **0** | | **119,117** | |

### SI-1c: Distribution of PacBio HiFi-Reads and PacBio CLR from JCV37772 Cell Line DNA

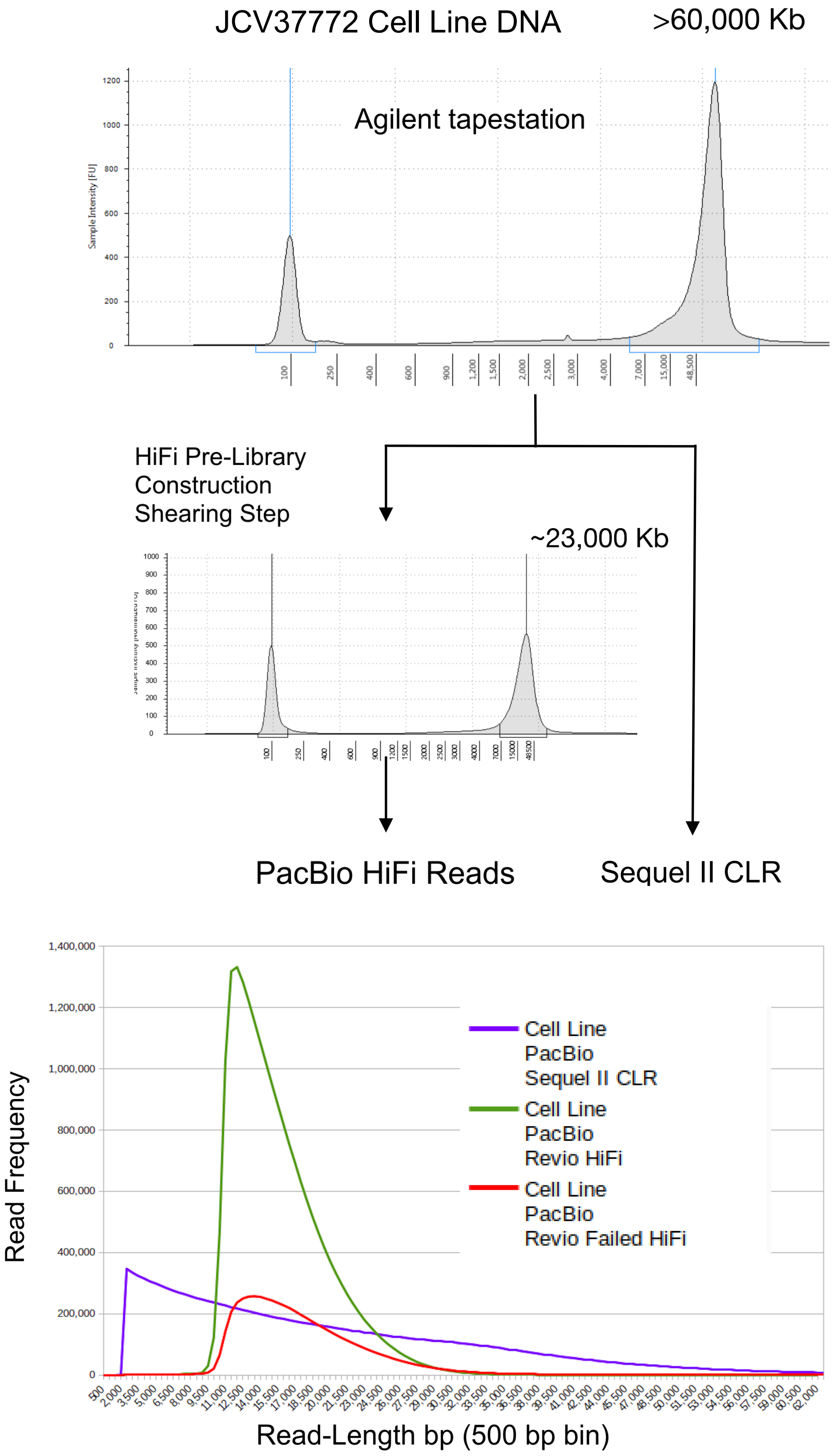


| | | PacBio Revio HiFi-Reads: HiFi-Reads | | PacBio Revio HiFi-Reads: Failed HiFi-Reads | | PacBio Sequel II CLR | |
|---|---|---|---|---|---|---|---|
| **Read-Length Category** | | **Cell Line PacBio Revio HiFi (≥ 2 kb)** | **Coverage X (3.1 Gb)** | **Cell Line Failed PacBio Revio HiFi (≥2 kb)** | **Coverage X (3.1 Gb)** | **Cell Line PacBio Sequel II CLR (≥2 kb)** | **Coverage X (3.1 Gb)** |
| Total | Mean Read Length (bp) | 15,290 | | 17,736 | | 19,384 | |
| | Median Read Length | 14,375 | | 16,240 | | 15,848 | |
| | Max Read Length | 61,359 | | 245,672 | | 348,239 | |
| | Min Read Length | 2,000 | | 2,001 | | 2,000 | |
| | **Total bases** | **300,981,218,795** | **97.091** | **98,826,236,167** | **31.879** | **269,732,456,621** | **87.01** |
| | Total Read Count | 19,685,117 | | 5,572,122 | | 13,914,885 | |
| | **Read Length N50 (bp)** | **15,460** | | **18,058** | | **28,362** | |
| | Read Length N90 | 11,426 | | 12,351 | | 10,205 | |
| | Read Length L50 | 7,829,515 | | 2,062,305 | | 3,351,638 | |
| | Read Length L90 | 16,890,551 | | 4,691,404 | | 9,280,768 | |
| 2.5k (2k to <3k) | Read Count | 995 | | 1,718 | | 680,925 | |
| | Bases | 2,531,368 | **0.001** | 4,323,804 | **0.001** | 1,697,941,714 | **0.55** |
| | **Read Length N50 (bp)** | **2,603** | | **2,573** | | **2,540** | |
| 3.5k (3k to <4k) | Read Count | 1,361 | | 1,981 | | 638,466 | |
| | Bases | 4,770,370 | **0.002** | 6,962,403 | **0.002** | 2,231,362,988 | **0.72** |
| | **Read Length N50** | **3,551** | | **3,555** | | **3,528** | |
| 4.5k (4k to <5k) | Read Count | 1,895 | | 2,625 | | 604,510 | |
| | Bases | 8,603,162 | **0.003** | 11,870,402 | **0.004** | 2,717,818,883 | **0.88** |
| | **Read Length N50** | **4,585** | | **4,570** | | **4,522** | |
| 7.5k (5k to <10k) | Read Count | 178,556 | | 49,965 | | 2,614,874 | |
| | Bases | 1,673,724,251 | **0.540** | 432,007,015 | **0.139** | 19,362,954,265 | **6.25** |
| | **Read Length N50** | **9,735** | | **9,436** | | **7,774** | |
| 12.5k (10k to <15k) | Read Count | 10,856,862 | | 2,160,052 | | 2,104,058 | |
| | Bases | 136,376,730,746 | **43.992** | 27,675,376,396 | **8.928** | 26,108,719,303 | **8.42** |
| | **Read Length N50** | **12,696** | | **13,032** | | **12,614** | |
| 17.5k (15k to <20k) | Read Count | 6,125,424 | | 1,904,578 | | 1,709,383 | |
| | Bases | 104,811,127,808 | **33.810** | 32,885,190,253 | **10.608** | 29,776,308,617 | **9.61** |
| | **Read Length N50** | **17,097** | | **17,326** | | **17,559** | |
| 22.5k (20k to <25k) | Read Count | 2,018,991 | | 927,859 | | 1,406,441 | |
| | Bases | 44,356,750,344 | **14.309** | 20,542,187,137 | **6.627** | 31,528,622,710 | **10.17** |
| | **Read Length N50** | **21,861** | | **22,102** | | **22,516** | |
| 37.5k (25k to <50k) | Read Count | 501,021 | | 481,269 | | 3,634,729 | |
| | Bases | 13,746,330,942 | **4.434** | 14,286,572,279 | **4.609** | 124,817,812,462 | **40.26** |
| | **Read Length N50** | **26,925** | | **28,515** | | **34,821** | |
| 62.5k (50k to <75k) | Read Count | 12 | | 29,655 | | 475,805 | |
| | Bases | 649,804 | **0.000** | 1,773,682,189 | **0.572** | 27,618,270,301 | **8.91** |
| | **Read Length N50** | **52,794** | | **59,734** | | **57,292** | |
| 87.5k (75k to <100k) | Read Count | 0 | | 8,336 | | 42,555 | |
| | Bases | 0 | **0.000** | 709,233,299 | **0.229** | 3,515,288,739 | **1.13** |
| | **Read Length N50** | **0** | | **84,733** | | **81,541** | |
| 100k (>= 100k) | Read Count | 0 | | 4,084 | | 3,139 | |
| | Bases | 0 | **0.000** | 498,830,990 | **0.161** | 357,356,639 | **0.12** |
| | **Read Length N50** | **0** | | **119,114** | | **108,075** | |

SI-1. (a) Length Distribution of JCV Blood DNA and its ONT-Reads. A 5 kb cut-off was applied to the ONT HAC- and SUP-reads. There was a slightly higher coverage of SUP-reads compared to HAC-reads due to a slightly greater number reads crossing the Q9 quality threshold using the SUP model. (b) Length Distribution of JCV Blood DNA and its PacBio HiFi-Reads. Failed HiFi-reads refer to HiFi-reads that have not passed the PacBio's quality threshold of Q20. Failed HiFi-reads are generally less accurate but longer. (c) Length Distribution of JCV37772 Cell Line DNA and its Pacbio HiFi-Reads and PacBio-CLR. A 2 kb cut-off was applied to the Pacbio-CLR. Though an older technology and is much less accurate, CLR has many more reads above 20 kb. The right panels denote the length profiles of input DNA and the lengths of the insuring reads.
