## Supplementary material for "The Revised Diploid Genome Sequence of an Individual Human: An Optimized Assembly Workflow for Scaling of near Telomere-to-Telomere Assemblies": SI_2.pdf

### SI-2: Errors and Read Profiles

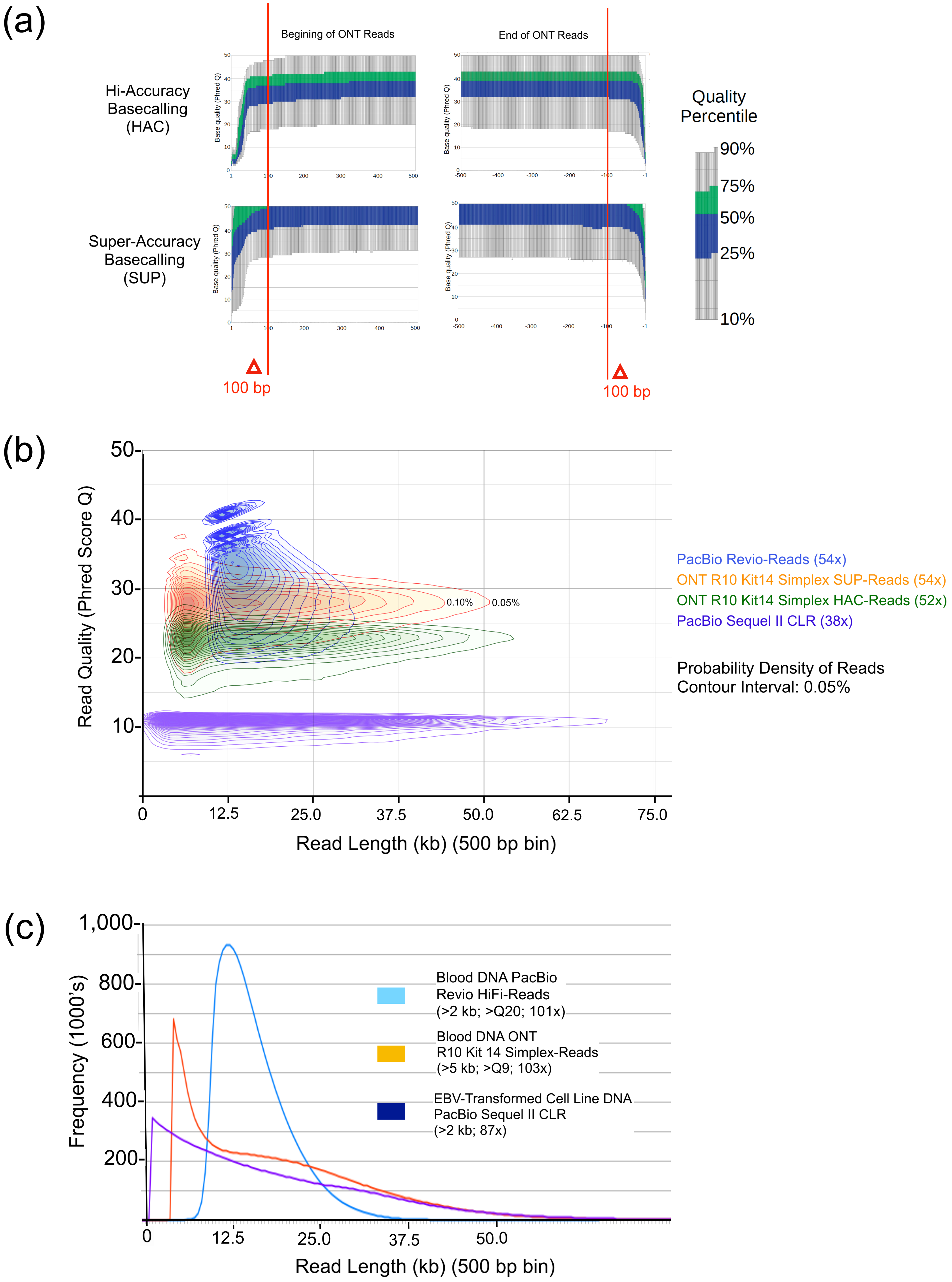

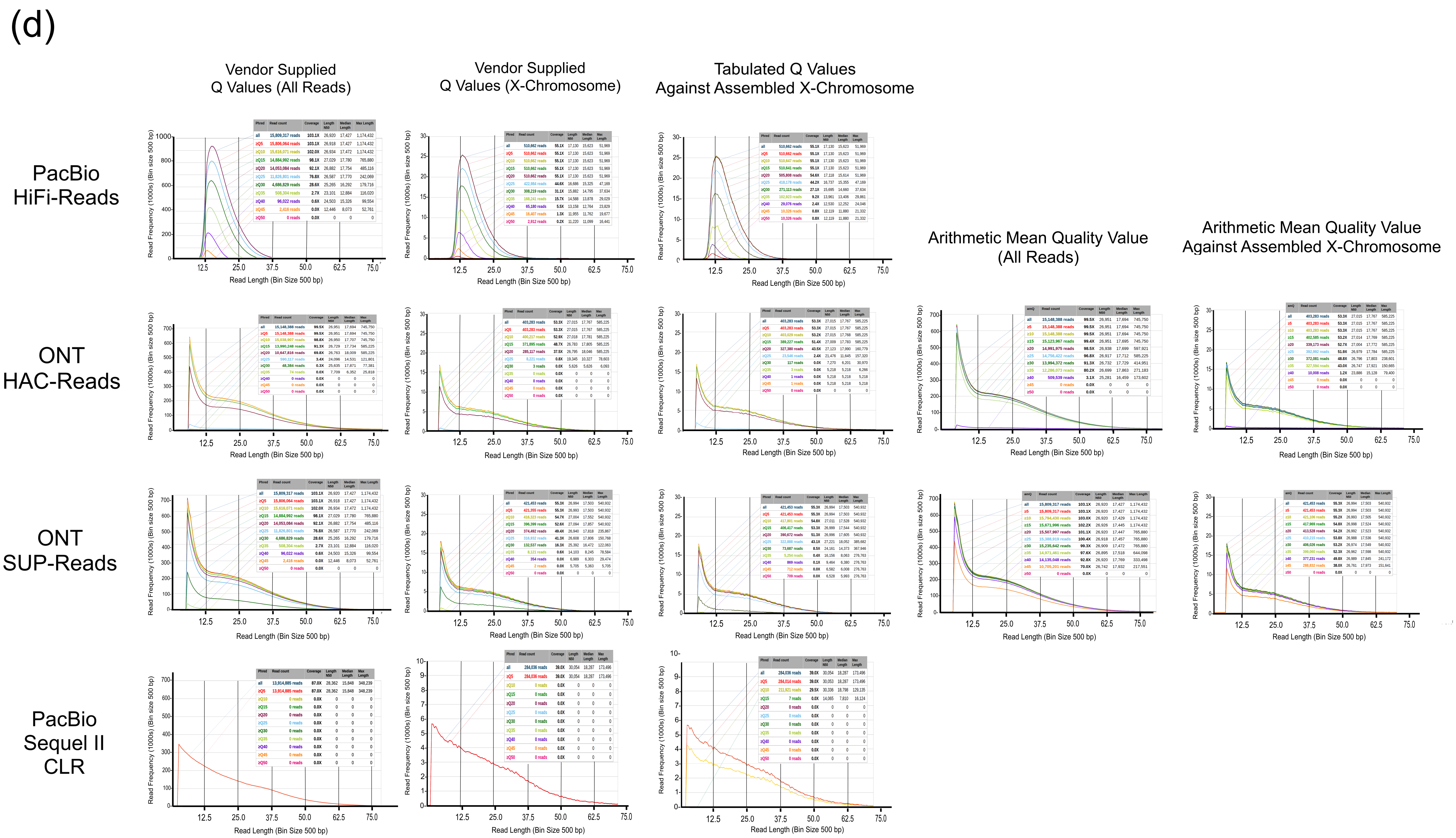
(d)
Vendor Supplied Q Values (All Reads)
Vendor Supplied Q Values (X-Chromosome)
Tabulated Q Values Against Assembled X-Chromosome
Arithmetic Mean Quality Value (All Reads)
Arithmetic Mean Quality Value Against Assembled X-Chromosome
PacBio HiFi-Reads
ONT HAC-Reads
ONT SUP-Reads
PacBio Sequel II CLR
Read Frequency (1000s) (Bin size 500 bp)
Read Length (Bin Size 500 bp)
12.5
25.0
37.5
50.0
62.5
75.0

SI-2. Length and Quality Distribution of Long-Reads.

(a) Per Base Quality Score of the First and Last 100 Bases of ONT HAC- and SUP-Reads, as Estimated by the Dorado Basecaller ( https://github.com/nanoporetech/dorado). We trimmed the less accurate 100 bp from each end of the input reads as our standard assembly condition. (b) The Tabulated Accuracy of Long-Reads in Relation to their Length. The read-accuracy was calculated by aligning the reads onto the assembled HuRef2.0 ChrX. ChrX was used to avoid mistaking allelic differences as sequencing errors. The contour plots show the probability density function of distribution, with each higher inner contour line depicting a more likely region at 0.05% increments. (c) Length Distribution of Each Type of Long-Reads. ONT HAC- and SUP-reads have the same length distribution profile. We applied a 5 kb cut-off for ONT-reads and a 2 kb cut-off for PacBio HiFi-reads and PacBio CLR. (d) Length Distribution of each Long Read Stratified by Vendor Supplied Read Q-Values, Tabulated Q-Values, and Arithmetic Mean Quality Values.
