## Supplementary material for "The Revised Diploid Genome Sequence of an Individual Human: An Optimized Assembly Workflow for Scaling of near Telomere-to-Telomere Assemblies": SI_3.pdf

### SI-3: The Contributions of Error-Correction and Coverage on Assembly Continuity

Columns 1–4 are **HAC** (Hi-Accuracy Basecalling); columns 5–8 are **SUP** (Super-Accuracy Basecalling). Column numbers 0–3 are Rounds of Pre-Assembly Error Correction.

#### 1 Flow Cell (~35x)

| Contig Stats | HAC 0: Error Correction r=0 | HAC 1: Error Correction r=1 | HAC 2: Error Correction r=2 | HAC 3: Error Correction r=3 | SUP 0: Error Correction r=0 | SUP 1: Error Correction r=1 | SUP 2: Error Correction r=2 | SUP 3: Error Correction r=3 |
|---|---|---|---|---|---|---|---|---|
| Assembly Size (bp) | 3,041,231,065 | 3,068,822,414 | 3,079,604,565 | 3,066,181,830 | 3,136,595,009 | 3,061,824,516 | 3,064,514,992 | 3,074,811,689 |
| Contig Count | 288 | 276 | 177 | 177 | 208 | 149 | 153 | 176 |
| NG50 (3.0 Gb) (bp) | 59,851,345 | 75,710,714 | 98,523,139 | 132,633,183 | 78,480,784 | 89,865,481 | 102,647,800 | 119,754,372 |
| NG90 (3.0 Gb) (bp) | 12,735,938 | 14,059,467 | 31,698,308 | 24,042,316 | 22,347,169 | 24,050,262 | 25,326,008 | 31,717,208 |
| NG99 (3.0 Gb) (bp) | 1,888,628 | 2,713,448 | 4,662,563 | 8,076,297 | 6,457,984 | 5,625,947 | 6,475,769 | 12,727,664 |
| LG50 (3.0 Gb) | 17 | 15 | 11 | 10 | 14 | 11 | 10 | 10 |
| LG90 (3.0 Gb) | 62 | 47 | 31 | 29 | 40 | 34 | 31 | 28 |
| LG99 (3.0 Gb) | 111 | 83 | 51 | 46 | 60 | 50 | 46 | 41 |
| Mean (bp) | 10,559,830 | 11,118,922 | 17,398,896 | 17,323,061 | 15,079,784 | 20,549,158 | 20,029,510 | 17,470,521 |
| Max (bp) | 138,635,340 | 182,470,112 | 190,597,440 | 190,045,885 | 171,176,277 | 203,299,583 | 248,301,786 | 202,880,204 |
| Min (bp) | 16,320 | 19,634 | 7,275 | 13,077 | 8,134 | 13,464 | 20,795 | 29,700 |
| Chromosomes (no gaps) | 0 | 0 | 2 | 5 | 1 | 4 | 3 | 4 |
| Total Gaps | >148 | >125 | >94 | >73 | >103 | >85 | >80 | >72 |
| Total Gaps (exclude Y) | 118 | 95 | 64 | 43 | 73 | 55 | 50 | 42 |
| chr1 | 10 | 7 | 5 | 3 | 3* | 4 | 0 | 2* |
| chr2 | 5 | 1 | 1 | 2 | 4 | 3 | 3 | 2 |
| chr3 | 3 | 4 | 2 | 1 | 1 | 0 | 0 | 0 |
| chr4 | 4 | 3 | 2 | 2 | 3 | 2 | 2 | 2 |
| chr5 | 4 | 4 | 0 | 0 | 1 | 0 | 1 | 0 |
| chr6 | 2 | 4* | 1 | 1 | 0 | 2 | 1 | 1 |
| chr7 | 7 | 1 | 1 | 0 | 4 | 3 | 3 | 1 |
| chr8 | 7 | 5 | 3 | 4 | 4 | 3 | 3 | 3 |
| chr9 | 6 | 6 | 3 | 1 | 5 | 5 | 4 | 2 |
| chr10 | 2 | 2 | 1 | 0 | 1 | 0 | 0 | 0 |
| chr11 | 4 | 3 | 2 | 1 | 4 | 2 | 2 | 3 |
| chr12 | 2 | 1 | 0 | 0 | 1 | 0 | 1 | 0 |
| chr13 | 4 | 4 | 1 | 2 | 3 | 1 | 1 | 1 |
| chr14 | 3* | 3 | 2 | 2 | 1 | 2 | 2 | 2 |
| chr15 | 5 | 1 | 2 | 3 | 6* | 2 | 1 | 1* |
| chr16 | 8 | 7* | 11 | 4 | 4 | 4 | 3 | 6 |
| chr17 | 5 | 4 | 3 | 2 | 4 | 3 | 3 | 1 |
| chr18 | 2 | 1 | 2 | 2 | 2 | 1 | 1 | 1 |
| chr19 | 9 | 6 | 3 | 2 | 1 | 2 | 2 | 2 |
| chr20 | 3 | 4 | 2 | 1 | 1 | 1 | 1 | 1 |
| chr21 | 3 | 4 | 2 | 0 | 3 | 1 | 3 | 2 |
| chr22 | 4* | 5 | 4 | 2 | 3 | 3 | 4 | 2 |
| chrX | 16 | 15 | 11 | 8 | 14 | 11 | 9 | 7 |
| chrY | >30 | >30 | >30 | >30 | >30 | >30 | >30 | >30 |

#### 2 Flow Cells (~67x)

| Contig Stats | HAC 0: Error Correction r=0 | HAC 1: Error Correction r=1 | HAC 2: Error Correction r=2 | HAC 3: Error Correction r=3 | SUP 0: Error Correction r=0 | SUP 1: Error Correction r=1 | SUP 2: Error Correction r=2 | SUP 3: Error Correction r=3 |
|---|---|---|---|---|---|---|---|---|
| Assembly Size (bp) | 3,052,102,655 | 3,128,872,115 | 3,064,593,368 | 3,074,258,092 | 3,163,645,556 | 3,079,761,246 | 3,071,548,880 | 3,067,174,324 |
| Contig Count | 105 | 144 | 85 | 69 | 74 | 102 | 78 | 66 |
| NG50 (3.0 Gb) (bp) | 127,147,169 | 91,918,486 | 134,718,727 | 144,806,205 | 114,534,593 | 133,897,282 | 133,991,777 | 135,164,656 |
| NG90 (3.0 Gb) (bp) | 33,419,086 | 28,103,510 | 57,936,771 | 59,877,548 | 60,948,026 | 45,306,331 | 62,322,749 | 61,011,566 |
| NG99 (3.0 Gb) (bp) | 9,321,215 | 13,711,166 | 11,235,488 | 20,464,654 | 34,805,070 | 20,969,793 | 33,729,698 | 30,834,895 |
| LG50 (3.0 Gb) | 10 | 12 | 10 | 9 | 10 | 9 | 10 | 10 |
| LG90 (3.0 Gb) | 29 | 33 | 24 | 21 | 24 | 25 | 23 | 21 |
| LG99 (3.0 Gb) | 40 | 47 | 32 | 27 | 30 | 33 | 29 | 26 |
| Mean (bp) | 29,067,644 | 21,728,279 | 36,054,040 | 44,554,465 | 42,751,967 | 30,193,738 | 39,378,832 | 46,472,338 |
| Max (bp) | 184,454,705 | 202,895,237 | 192,406,875 | 248,435,310 | 192,064,441 | 242,580,376 | 202,393,462 | 202,385,309 |
| Min (bp) | 14,325 | 17,340 | 16,791 | 6,431 | 9,114 | 18,527 | 7,575 | 37,057 |
| Chromosomes (no gaps) | 7 | 6 | 9 | 14 | 10 | 13 | 11 | 12 |
| Total Gaps | 36 | 56 | 25 | 21 | 26 | 25 | 25 | 20 |
| Total Gaps (exclude Y) | 30 | 49 | 19 | 15 | 19 | 20 | 17 | 14 |
| chr1 | 4 | 4 | 1 | 0 | 3 | 2 | 1 | 1 |
| chr2 | 2* | 6 | 1 | 3 | 1* | 0 | 3 | 2 |
| chr3 | 1 | 0 | 1 | 0 | 2 | 0 | 0 | 0 |
| chr4 | 3 | 2 | 0 | 0 | 1 | 0 | 1 | 0 |
| chr5 | 0 | 0 | 1 | 0 | 0 | 1 | 1 | 0 |
| chr6 | 1* | 0 | 0 | 0 | 0 | 0 | 0 | 0 |
| chr7 | 0 | 2 | 0 | 0 | 0 | 0 | 1 | 0 |
| chr8 | 0 | 2* | 1 | 0 | 1* | 0 | 0 | 0 |
| chr9 | 0 | 6 | 1 | 2 | 3* | 6 | 1 | 0 |
| chr10 | 0 | 1 | 0 | 0 | 0 | 0 | 0 | 0 |
| chr11 | 1 | 0 | 0 | 0 | 0 | 1 | 0 | 0 |
| chr12 | 0 | 0 | 0 | 0 | 0 | 0 | 0 | 0 |
| chr13 | 1 | 3* | 1 | 2 | 2 | 0 | 1 | 1 |
| chr14 | 2 | 2 | 1 | 1 | 0 | 2 | 1 | 1 |
| chr15 | 1 | 4 | 1 | 0 | 1 | 2 | 2 | 2 |
| chr16 | 1 | 5 | 5 | 1 | 1 | 3 | 2 | 2 |
| chr17 | 2 | 1 | 2 | 3 | 1 | 1 | 0 | 1 |
| chr18 | 2 | 1 | 1 | 0 | 1 | 0 | 0 | 0 |
| chr19 | 1 | 1 | 0 | 0 | 0 | 0 | 0 | 0 |
| chr20 | 0 | 0 | 0 | 0 | 0 | 0 | 0 | 1 |
| chr21 | 1 | 3 | 1 | 1 | 0 | 1 | 0 | 1 |
| chr22 | 2 | 2 | 1 | 1 | 1 | 1 | 1 | 1 |
| chrX | 5 | 4 | 0 | 1 | 1 | 0 | 2 | 1 |
| chrY | 6 | 7 | 6 | 6 | 7 | 5 | 8 | 6 |

#### 3 Flow Cells (~103x)

| Contig Stats | HAC 0: Error Correction r=0 | HAC 1: Error Correction r=1 | HAC 2: Error Correction r=2 | HAC 3: Error Correction r=3 | SUP 0: Error Correction r=0 | SUP 1: Error Correction r=1 | SUP 2: Error Correction r=2 | SUP 3: Error Correction r=3 |
|---|---|---|---|---|---|---|---|---|
| Assembly Size (bp) | 3,199,004,399 | 3,120,162,269 | 3,078,139,492 | 3,022,294,422 | 3,099,752,068 | 3,092,461,989 | 3,060,121,034 | 3,068,164,550 |
| Contig Count | 171 | 184 | 104 | 72 | 84 | 88 | 76 | 128 |
| NG50 (3.0 Gb) (bp) | 87,947,079 | 91,924,179 | 133,925,827 | 144,857,908 | 134,729,557 | 133,929,771 | 134,741,182 | 135,308,536 |
| NG90 (3.0 Gb) (bp) | 30,679,604 | 31,230,152 | 57,202,429 | 61,074,292 | 56,898,033 | 53,841,491 | 74,694,085 | 58,622,792 |
| NG99 (3.0 Gb) (bp) | 16,658,125 | 10,139,935 | 25,567,020 | 20,103,542 | 20,455,170 | 23,937,415 | 43,194,962 | 25,191,235 |
| LG50 (3.0 Gb) | 14 | 13 | 10 | 9 | 9 | 10 | 10 | 9 |
| LG90 (3.0 Gb) | 35 | 35 | 24 | 20 | 22 | 24 | 22 | 21 |
| LG99 (3.0 Gb) | 46 | 50 | 31 | 27 | 29 | 31 | 27 | 27 |
| Mean (bp) | 18,707,628 | 16,957,404 | 29,597,495 | 41,976,311 | 36,901,810 | 35,141,614 | 40,264,750 | 23,970,036 |
| Max (bp) | 168,711,437 | 145,983,597 | 203,224,926 | 248,800,294 | 248,601,805 | 202,904,933 | 202,730,645 | 242,590,848 |
| Min (bp) | 7,770 | 10,801 | 14,302 | 10,817 | 17,340 | 26,529 | 13,989 | 7,295 |
| Chromosomes (no gaps) | 2 | 2 | 11 | 14 | 10 | 11 | 12 | 14 |
| Total Gaps | 54 | 55 | 28 | 14 | 19 | 22 | 15 | 12 |
| Total Gaps (exclude Y) | 47 | 46 | 22 | 11 | 15 | 19 | 13 | 11 |
| chr1 | 4* | 2 | 2 | 0 | 0 | 1 | 1 | 3 |
| chr2 | 5 | 2 | 2 | 1 | 0 | 2 | 1 | 0 |
| chr3 | 2 | 1 | 0 | 0 | 0 | 0 | 1 | 0 |
| chr4 | 3 | 2 | 0 | 0 | 1 | 0 | 0 | 0 |
| chr5 | 1 | 2 | 1 | 0 | 1 | 0 | 0 | 0 |
| chr6 | 1 | 3 | 0 | 0 | 0 | 0 | 0 | 0 |
| chr7 | 1 | 1 | 0 | 0 | 1 | 2 | 1 | 0 |
| chr8 | 1* | 0 | 0 | 0 | 0 | 0 | 0 | 1 |
| chr9 | 2 | 3 | 1 | 0 | 1 | 3 | 1 | 0 |
| chr10 | 1 | 2 | 0 | 0 | 0 | 0 | 0 | 0 |
| chr11 | 1 | 2 | 0 | 0 | 0 | 0 | 0 | 0 |
| chr12 | 4 | 1 | 1 | 0 | 0 | 0 | 0 | 0 |
| chr13 | 3 | 3 | 4* | 1 | 1 | 1 | 2* | 1 |
| chr14 | 4 | 1 | 3 | 1 | 0 | 2 | 1 | 1 |
| chr15 | 1 | 4 | 1 | 1 | 1 | 2 | 1* | 1 |
| chr16 | 2 | 2 | 2 | 2 | 1 | 1 | 1 | 1 |
| chr17 | 4 | 4 | 2* | 1 | 1 | 1 | 0 | 1 |
| chr18 | 1 | 2 | 0 | 0 | 1 | 0 | 0 | 0 |
| chr19 | 1 | 2 | 0 | 0 | 1 | 0 | 0 | 0 |
| chr20 | 0 | 0 | 0 | 0 | 0 | 0 | 0 | 0 |
| chr21 | 0 | 1 | 1 | 1 | 2 | 1 | 2* | 1 |
| chr22 | 1 | 2 | 2 | 1 | 1 | 2 | 1* | 1 |
| chrX | 4 | 4 | 0 | 2 | 2 | 1 | 0 | 0 |
| chrY | 7 | 9 | 6 | 3 | 4 | 3 | 2 | 1 |

Comparison to T2T-CHM13v2

- **0** Chromosome - 0 Gaps (T2T)
- **1** Chromosome - 1 Gap
- **2** Chromosome - 2 Gaps
- **3** Chromosome - 3 Gaps
- **4+** Chromosome - > 3 Gaps
- Chromosome - 1 or more miss-assembly

SI-3. Base Calling Accuracy (HAC- vs SUP-Reads), Coverage, and the Number of Error-Correction Rounds on Assembly Contiguity. Contiguity of each chromosome was determined by aligning assembled contigs to T2T-CHM13v2.0. Assembled chromosomes with 0, 1, 2, or 3 gaps were highlighted in green, dark blue, light blue and yellow, respectively. Chromosomes with mis-assemblies, usually a chimera between different chromosomes, were highlighted in red. Assemblies using SUP-reads were consistently more contiguous than those using HAC-reads. The increase in assembly contiguity with more rounds of error correction was most pronounced at low coverage, with diminishing return going from two to three rounds of correction and with 2 to 3 flowcells.
