## Supplementary material for "The Revised Diploid Genome Sequence of an Individual Human: An Optimized Assembly Workflow for Scaling of near Telomere-to-Telomere Assemblies": SI_4.pdf

### SI-4: hifiasm Parameters for Sub-Assemblies A to N

**A**

**Standard ONT input for assembly:**

- 3 ONT R10 flow cells, K14 chemistry
- **SUP** reads (99x)
- Reads **>Q9** and **> 5 kb**; **trim = 100**
- 2 rounds of pre-assembly error-correction **$r$ = 2**

**Standard hifiasm.v24 assembly condition:**

- k-mer Length, **$k$ = 51**
- Minimizer Window Size, **$w$ = 51**
- Expected Error Rate, **$bw$ = 0.05**
- Upper Frequency Limit, **$D$ = 5**

| | |
|---|---|
| A | **Standard condition: standard assembly condition using standard error-corrected reads** |
| B | **Standard condition using HAC reads** |
| C | **Standard condition with $r$ = 0 (instead of $r$ = 2)** |
| D | **Standard condition with $r$ = 1 (instead of $r$ = 2)** |
| E | **Standard condition with $bw$ = 0.02 (instead of $bw$ = 0.05)** |
| F | **Standard condition with $D$ = 10 (instead of $D$ = 5)** |
| G | **Standard condition with $k$ = 63; $w$ = 63 (instead of $k$ = 51; $w$ = 51)** |
| H | **Standard condition with *trim* = 0 bp (instead *trim* = 100 bp)** |
| I | **Standard condition with $r$ = 3 (instead of $r$ = 2)** |
| J | **Standard condition with *trim* = 500 bp; $r$ = 3 (instead *trim* = 100 bp; $r$ = 2)** |
| K | **Standard condition using longer reads with greater acuracy: >= amQ15; >= 10 kb; $r$ = 3** |
| L | **Standard condition with >= amQ40 as primary reads; and reads >= 15 kb and < amQ40 as secondary reads** |
| M | **Standard condition with >= amQ40 as primary reads; and reads >= 15 kb and < amQ40 as secondary reads; $bw$ = 0.02** |
| N | **Standard condition with >= amQ40 as primary reads; and reads >= 15 kb and < amQ40 as secondary reads; bw = 0.02; $r$ = 3** |

SI-4. Conditions and hifiasm v0.24.0 Parameters used to Generate the 14 Sub-Assemblies (A-N) from ONT-Reads. Condition A represents the standard baseline, and uses default hifiasm parameters except for r=2. When reads were filtered according to their quality score, we used the Arithmetic mean Quality score (amQ), defined as the arithmetic mean of the quality score of each base in the read, as output in the FASTQ file.
