## Supplementary material for "The Revised Diploid Genome Sequence of an Individual Human: An Optimized Assembly Workflow for Scaling of near Telomere-to-Telomere Assemblies": SI_5.pdf

### SI-5: JCV Sub-Assemblies at Different ONT Sequence Coverage

#### JCV Sub-Assemblies: Assembly Conditions A to N

#### JCV Composite-Assembly

##### 1 Flowcell (~35x)

| Contig Stats | A | B | C | D | E | F | G | H | I | J | K | L | M | N |
|---|---|---|---|---|---|---|---|---|---|---|---|---|---|---|
| Assembly Size (bp) | 3,058,338,596 | 3,080,122,575 | 3,160,361,862 | 3,045,813,863 | 3,056,788,388 | 3,075,002,198 | 3,073,196,623 | 3,064,514,992 | 3,063,706,546 | 3,091,085,362 | 3,083,147,597 | 3,065,866,867 | 3,069,858,780 | 3,093,153,866 |
| Contig Count | 161 | 200 | 264 | 146 | 153 | 165 | 163 | 153 | 166 | 218 | 230 | 167 | 160 | 220 |
| NG50 (3.0 Gb) (bp) | 102,296,622 | 101,389,781 | 78,249,222 | 85,184,942 | 99,757,501 | 98,795,403 | 102,184,375 | 102,647,800 | 132,788,159 | 106,377,225 | 88,209,368 | 96,815,599 | 90,729,230 | 91,635,406 |
| NG90 (3.0 Gb) (bp) | 25,572,982 | 24,050,917 | 19,760,677 | 23,440,231 | 25,311,022 | 25,114,990 | 25,720,871 | 25,326,008 | 25,082,321 | 27,637,374 | 19,563,642 | 21,628,322 | 25,028,253 | 15,254,842 |
| NG99 (3.0 Gb) (bp) | 5,468,645 | 4,968,718 | 5,022,508 | 4,835,092 | 6,610,508 | 11,496,755 | 5,359,724 | 6,475,769 | 9,497,351 | 8,914,334 | 4,818,170 | 5,913,501 | 6,273,375 | 5,617,134 |
| LG50 (3.0 Gb) | 11 | 11 | 15 | 12 | 11 | 10 | 10 | 10 | 9 | 10 | 12 | 12 | 11 | 12 |
| LG90 (3.0 Gb) | 31 | 32 | 47 | 36 | 31 | 31 | 30 | 31 | 28 | 29 | 41 | 35 | 31 | 38 |
| LG99 (3.0 Gb) | 47 | 52 | 75 | 55 | 47 | 46 | 49 | 46 | 43 | 44 | 66 | 55 | 50 | 64 |
| Mean (bp) | 18,995,892 | 15,400,613 | 11,971,068 | 20,861,739 | 19,979,009 | 18,636,377 | 18,853,967 | 20,029,510 | 18,456,064 | 14,179,291 | 13,404,990 | 18,358,484 | 19,186,617 | 14,059,790 |
| Max (bp) | 202,863,574 | 190,687,658 | 148,530,779 | 203,301,561 | 202,866,167 | 203,171,428 | 202,868,490 | 248,301,786 | 248,371,667 | 232,975,508 | 192,053,714 | 191,038,787 | 191,051,765 | 182,693,416 |
| Min (bp) | 8,289 | 16,252 | 15,738 | 31,857 | 8,289 | 8,289 | 8,289 | 20,795 | 8,289 | 10,270 | 17,140 | 16,607 | 16,607 | 28,014 |
| T2T Chromosomes | 3 | 2 | 2 | 5 | 2 | 2 | 6 | 3 | 3 | 4 | 2 | 2 | 2 | 2 |
| Total Gaps | >79 | >92 | >113 | >90 | >81 | >80 | >83 | >80 | >74 | >81 | >110 | >99 | >92 | >119 |
| Total Gaps (exclude Y) | 49 | 62 | 83 | 60 | 51 | 50 | 53 | 50 | 44 | 51 | 80 | 69 | 62 | 89 |
| chr1 | 2 | 3 | 5 | 5 | 2 | 1 | 2 | 0 | **0** | 1 | 2 | 3 | 3 | 2 |
| chr2 | 3 | 4 | 3 | 3 | 4 | 5 (Chimeric) | 4 | 3 | **2** | 2 | 4 | 2 | 3 | 3 |
| chr3 | 0 | 1 | 1 | **0** | 0 | 0 | 0 | 0 | 1 | 2 | 3 | 2 | 1 | 3 |
| chr4 | 2 | 2 | 4 | 3 | 1 | 2 | 2 | 2 | 2 | 2 | **0** | 2 | 1 | 2 |
| chr5 | 1 | 2 | 3 | 2 | 1 | 2 | 0 | 1 | **0** | 0 | 3 | 1 | 1 | 0 |
| chr6 | 1 | 2 | 2 | 1 | 1 | 1 | 1 | 1 | **1** | 2 | 2 | 1 | 1 | 1 |
| chr7 | 2 | 0 | 3 | 4 | 3 | 2 | 2 | 3 | 3 | **1** | 6 | 4 | 4 | 4 |
| chr8 | 3 | 4 | 5 | 5 | **1** | 2 | 2 | 3 | 3 | 2 | 5 | 7 | 5 | 5 |
| chr9 | 2 | 5 | 5 | 4 | 4 | 4 (Chimeric) | 3 | 4 | **1** | 4 | 3 | 4 | 3 | 4 |
| chr10 | 0 | 0 | 1 | **0** | 1 | 0 | 0 | 0 | 1 | 0 | 0 | 0 | 1 | 0 |
| chr11 | 1 | 1 | 3 | **0** | 1 | 1 | 1 | 2 | 2 | 2 | 4 | 1 | 1 | 6 |
| chr12 | 1 | 3 | 0 | **0** | 1 | 1 | 0 | 1 | 0 | 0 | 2 | 3 | 0 | 2 |
| chr13 | 1 | 2 | 2 | 2 | 2 | 1 | 3 | 1 | 2 | 2 | 2 | 2 | 2 | **1** |
| chr14 | 2 | 2 | 4 | 4 | 2 (Chimeric) | 3 | 3 | 2 | 2 | 4 | **1** | 2 | 2 | 2 |
| chr15 | 2 | 2 | 5 | 2 | 2 | 1 | 0 | 1 | 3 | **0** | 3 | 1 | 1 | 2 |
| chr16 | 4 | 3 | 9 | 4 | 6 | 5 | 5 | **3** | 5 | 4 | 6 | 8 | 7 | 5 |
| chr17 | 4 | 6 | 4 | 5 | 4 | 3 | 5 | 3 | 2 | **1** | 5 | 4 | 4 | 8 |
| chr18 | 0 | 2 | 1 | **0** | 0 | 1 | 0 | 1 | 1 | 1 | 2 | 2 | 1 | 1 |
| chr19 | 3 | 4 | 4 | 2 | 2 | 3 | 3 | 2 | **2** | 5 | 4 | 2 | 2 | 7 |
| chr20 | 2 | 1 | 0 | 2 | 2 | 2 | 2 | 1 | 1 | 1 | 2 | **0** | 0 | 1 |
| chr21 | 3 | 1 | 4 | **1** | 2 (Chimeric) | 1 | 3 | 3 | 2 | 1 | 2 | 2 | 1 | 2 |
| chr22 | 2 | 1 | 4 | 2 | 2 | 2 | 1 | 4 | **1** | 1 | 1 | 1 | 3 | 2 |
| chrX | 8 | 11 | 11 | 9 | **7** | 7 | 11 | 9 | 7 | 13 | 18 | 15 | 15 | 26 |
| chrY | >30 | >30 | >30 | >30 | >30 | >30 | **>30** | >30 | >30 | >30 | >30 | >30 | >30 | >30 |

→

| | |
|---|---|
| chr1 | 0 |
| chr2 | 2 |
| chr3 | 0 |
| chr4 | 0 |
| chr5 | 0 |
| chr6 | 1 |
| chr7 | 1 |
| chr8 | 1 |
| chr9 | 1 |
| chr10 | 0 |
| chr11 | 0 |
| chr12 | 0 |
| chr13 | 1 |
| chr14 | 1 |
| chr15 | 0 |
| chr16 | 3 |
| chr17 | 1 |
| chr18 | 0 |
| chr19 | 2 |
| chr20 | 0 |
| chr21 | 1 |
| chr22 | 1 |
| chrX | 7 |
| chrY | >30 |

D E H I J K L N

| | Composite Assembly ONT Contig (1 FC) | Composite Assembly ONT Scaffold (1 FC) |
|---|---|---|
| Assembly Size (bp) | 3,037,022,651 | 3,037,028,851 |
| Contig/ Scaffold Count | 86 | 24 |
| Complete Chromosomes | 10 | 10 |
| Gaps | 62 | 62 |
| NG50 (3.0 Gb) (bp) | 133,718,319 | 153,026,018 |
| NG90 (3.0 Gb) (bp) | 35,303,050 | 83,144,720 |
| NG99 (3.0 Gb) (bp) | 17,193,060 | 49,908,900 |
| LG50 (3.0 Gb) | 9 | 8 |
| LG90 (3.0 Gb) | 24 | 18 |
| LG99 (3.0 Gb) | 35 | 22 |
| Mean (bp) | 35,314,217 | 126,542,869 |
| Max (bp) | 248,371,667 | 248,371,667 |
| Min (bp) | 31,861 | 18,530,514 |
| Phase Block Count | 3,659 | |
| Phase Block NG50 (bp) | 1,490,519 | |

Comparison to T2T-CHM13v2

| | |
|---|---|
| 0 | Chromosome - 0 Gaps (T2T) |
| 1 | Chromosome - 1 Gap |
| 2 | Chromosome - 2 Gaps |
| 3 | Chromosome - 3 Gaps |
| 4+ | Chromosome - > 3 Gaps |
| | Chromosome - 1 or more miss-assembly |
| **N** | Selected for composite assembly |

##### 2 Flowcells (~67x)

| Contig Stats | A | B | C | D | E | F | G | H | I | J | K | L | M | N |
|---|---|---|---|---|---|---|---|---|---|---|---|---|---|---|
| Assembly Size (bp) | 3,067,510,349 | 3,083,123,677 | 3,113,041,308 | 3,105,785,406 | 3,067,899,359 | 3,075,301,841 | 3,068,067,509 | 3,071,548,880 | 3,076,180,198 | 3,083,023,025 | 3,064,532,066 | 3,063,893,003 | 3,062,283,949 | 3,052,056,569 |
| Contig Count | 69 | 77 | 99 | 92 | 74 | 69 | 67 | 78 | 77 | 92 | 82 | 66 | 67 | 63 |
| NG50 (3.0 Gb) (bp) | 135,159,782 | 136,304,727 | 102,926,441 | 134,755,481 | 134,637,139 | 133,989,483 | 134,153,889 | 133,991,777 | 133,889,243 | 124,154,236 | 135,160,607 | 146,304,026 | 135,151,193 | 138,680,322 |
| NG90 (3.0 Gb) (bp) | 60,999,483 | 60,958,124 | 46,602,497 | 49,485,710 | 46,249,048 | 60,931,782 | 58,392,270 | 62,322,749 | 58,226,014 | 58,266,643 | 61,050,079 | 60,942,101 | 66,791,705 | 80,436,116 |
| NG99 (3.0 Gb) (bp) | 12,826,016 | 25,111,777 | 11,556,980 | 20,791,519 | 12,577,537 | 25,168,951 | 15,422,927 | 33,729,698 | 25,472,188 | 22,380,979 | 21,609,262 | 25,275,548 | 25,120,560 | 42,329,395 |
| LG50 (3.0 Gb) | 9 | 9 | 11 | 10 | 10 | 10 | 9 | 10 | 10 | 10 | 9 | 8 | 8 | 9 |
| LG90 (3.0 Gb) | 21 | 21 | 26 | 23 | 24 | 22 | 22 | 23 | 23 | 23 | 21 | 20 | 21 | 19 |
| LG99 (3.0 Gb) | 28 | 27 | 35 | 31 | 33 | 29 | 30 | 29 | 29 | 30 | 28 | 27 | 27 | 24 |
| Mean (bp) | 44,456,672 | 40,040,567 | 31,444,862 | 33,758,537 | 41,458,099 | 44,569,592 | 45,792,052 | 39,378,832 | 39,950,392 | 33,511,120 | 37,372,342 | 46,422,621 | 45,705,731 | 48,445,342 |
| Max (bp) | 218,917,890 | 248,402,561 | 185,010,707 | 202,266,880 | 202,401,636 | 218,917,483 | 242,172,192 | 202,393,462 | 202,393,106 | 201,531,567 | 248,401,894 | 275,254,159 | 309,502,901 | 242,616,368 |
| Min (bp) | 12,824 | 10,617 | 6,912 | 11,283 | 12,824 | 17,080 | 17,080 | 7,575 | 7,639 | 13,961 | 28,981 | 16,607 | 16,607 | 31,014 |
| T2T Chromosomes | 10 | 13 | 7 | 11 | 9 | 10 | 11 | 11 | 11 | 10 | 12 | 14 | 11 | 18 |
| Total Gaps | 22 | 18 | 32 | 33 | 27 | 26 | 24 | 25 | 24 | 18 | 21 | 19 | 18 | 16 |
| Total Gaps (exclude Y) | 17 | 13 | 25 | 26 | 22 | 20 | 18 | 17 | 16 | 15 | 14 | 13 | 13 | 6 |
| chr1 | 3 | **0** | 2 | 4 | 1 | 2 (Chimeric) | 3 | 1 | 2 | 1 | **0** | 1 (Chimeric) | 1 (Chimeric) | 1 |
| chr2 | 1 | 2 | 1 | 1 | 2 | 2 (Chimeric) | 0 | 3 | 3 | 3 | 2 | 0 | 2 | **0** |
| chr3 | 0 | 0 | 1 | 0 | 0 | 0 | 0 | 0 | 0 | 0 | 0 | 0 | 0 | **0** |
| chr4 | 1 | 1 | 3 | 2 | 0 | 1 | 0 | 1 | 1 | 1 | 1 | 1 | 1 | **0** |
| chr5 | 0 | 1 | 0 | 0 | 0 | 1 | 0 | 1 | 0 | 0 | 0 | 0 | 1 | **0** |
| chr6 | 1 (Chimeric) | 1 | 1 | 0 | 1 (Chimeric) | 1 (Chimeric) | 0 | 0 | 0 | 0 | 0 | 0 | 1 (Chimeric) | **0** |
| chr7 | 0 | 0 | 0 | 0 | 1 | 0 | 1 | 1 | 1 | 1 | 1 | 0 | 1 | **0** |
| chr8 | 1 | 1 | 1 | 0 | 1 | 0 | 1 | 0 | 0 | 1 | 0 | **0** | 0 | 2 |
| chr9 | **0** | 0 | 2 | 2 | 3 | 2 | 1 | 1 | 1 | 1 | 1 | 2 (Chimeric) | 1 (Chimeric) | 0 |
| chr10 | 0 | 0 | 0 | 0 | 0 | 0 | 0 | 0 | 0 | 1 | 0 | 0 | 0 | **0** |
| chr11 | 0 | 0 | 1 | 0 | 1 | 0 | 0 | 0 | 0 | 0 | 0 | 0 | 0 | **0** |
| chr12 | 0 | 0 | 0 | 0 | 0 | 0 | 0 | 0 | 0 | 0 | 0 | 0 | 0 | **0** |
| chr13 | 1 | 1 | 3 | 2 | 1 | 2 (Chimeric) | 1 | 1 | 1 | 0 | 1 | 1 | 1 | **0** |
| chr14 | 1 | 0 | 2 | 1 | 1 | **0** | 2 | 1 | 0 | 1 | 1 (Chimeric) | 1 | 0 | 1 |
| chr15 | 2 | 1 | 1 | 2 | 2 | 2 (Chimeric) | 2 | 2 | 2 | 1 | **1** | 2 | 1 | 0 (shorter) |
| chr16 | 2 | 2 | 1 | 4 | 4 | 2 | 3 | 2 | 1 | 1 | 2 | 0 | **0** | 0 |
| chr17 | 1 (Chimeric) | 2 | 0 | 3 | 2 (Chimeric) | 1 (Chimeric) | 1 | **0** | 1 | 1 | 1 | 3 | 1 (Chimeric) | 1 |
| chr18 | 0 | 0 | 1 | 0 | 0 | 0 | 0 | 0 | 0 | 0 | 0 | 0 | 0 | **0** |
| chr19 | 0 | 0 | 0 | 0 | 0 | 0 | 0 | 0 | 0 | 0 | 0 | 0 | 0 | **0** |
| chr20 | 0 | 0 | 0 | 0 | 0 | 0 | 0 | 0 | 0 | 0 | 0 | 0 | 0 | **0** |
| chr21 | 1 | 0 | 1 | 1 | 0 | 2 (Chimeric) | 1 | **0** | 1 | 1 | 2 (Chimeric) | 1 | 1 | 0 |
| chr22 | 1 | 0 | 3 | 3 | 1 | 1 | 1 | 1 | 1 | 1 | 1 | **0** | 1 | 1 |
| chrX | 1 | 1 | 2 | 1 | 1 | 1 | 1 | 2 | 1 | 0 | 0 | 1 | **0** | 0 |
| chrY | 5 | 5 | 7 | 7 | 5 | 6 | 6 | 8 | 8 | 3 | 7 | 6 | **5** | 10 |

→

| | |
|---|---|
| chr1 | 0 |
| chr2 | 0 |
| chr3 | 0 |
| chr4 | 0 |
| chr5 | 0 |
| chr6 | 0 |
| chr7 | 0 |
| chr8 | 0 |
| chr9 | 0 |
| chr10 | 0 |
| chr11 | 0 |
| chr12 | 0 |
| chr13 | 0 |
| chr14 | 0 |
| chr15 | 1 |
| chr16 | 0 |
| chr17 | 0 |
| chr18 | 0 |
| chr19 | 0 |
| chr20 | 0 |
| chr21 | 0 |
| chr22 | 0 |
| chrX | 0 |
| chrY | 5 |

A F H K L M N

| | Composite Assembly ONT Contig (2 FC) | Composite Assembly ONT Scaffold (2 FC) |
|---|---|---|
| Assembly Size (bp) | 3,076,200,123 | 3,076,200,623 |
| Contig/ Scaffold Count | 29 | 24 |
| Complete Chromosomes | 22 | 22 |
| Gaps | 6 | 6 |
| NG50 (3.0 Gb) (bp) | 154,727,717 | 154,727,717 |
| NG90 (3.0 Gb) (bp) | 82,023,092 | 83,047,129 |
| NG99 (3.0 Gb) (bp) | 43,113,097 | 51,062,433 |
| LG50 (3.0 Gb) | 8 | 8 |
| LG90 (3.0 Gb) | 18 | 18 |
| LG99 (3.0 Gb) | 23 | 22 |
| Mean (bp) | 106,075,866 | 128,175,026 |
| Max (bp) | 248,401,894 | 248,401,894 |
| Min (bp) | 718,255 | 43,113,097 |
| Phase Block Count | 2,707 | |
| Phase Block NG50 (bp) | 2,044,076 | |

The final composite assembly was compiled from the best assembled chromosomes from sub-assemblies (A-N) indicted by pairs of double asterisks (** **)

##### 3 Flowcells (~103x)

| Contig Stats | A | B | C | D | E | F | G | H | I | J | K | L | M | N | F (2FC) |
|---|---|---|---|---|---|---|---|---|---|---|---|---|---|---|---|
| Assembly Size (bp) | 3,066,381,738 | 3,085,091,674 | 3,129,759,164 | 3,084,453,945 | 3,058,487,834 | 3,065,533,249 | 3,058,850,696 | 3,060,121,034 | 3,055,308,173 | 3,077,367,467 | 3,073,213,406 | 3,064,531,315 | 3,043,384,535 | 3,058,363,874 | 3,075,301,841 |
| Contig Count | 78 | 88 | 112 | 105 | 83 | 76 | 75 | 76 | 82 | 104 | 86 | 58 | 68 | 107 | 69 |
| NG50 (3.0 Gb) (bp) | 135,218,378 | 133,926,235 | 133,919,696 | 104,771,396 | 153,434,071 | 134,029,922 | 134,737,267 | 134,741,182 | 136,356,532 | 139,940,519 | 153,436,580 | 146,885,042 | 146,870,284 | 152,687,993 | 133,989,483 |
| NG90 (3.0 Gb) (bp) | 58,075,424 | 48,884,555 | 59,633,553 | 56,581,185 | 57,799,507 | 57,942,162 | 61,065,964 | 74,694,085 | 61,074,363 | 58,688,349 | 66,898,730 | 66,882,975 | 66,789,406 | 60,950,207 | 60,931,782 |
| NG99 (3.0 Gb) (bp) | 30,072,951 | 22,416,017 | 26,462,186 | 15,238,881 | 25,297,957 | 30,062,211 | 22,368,280 | 43,194,962 | 25,283,055 | 29,795,806 | 29,775,994 | 39,497,594 | 21,490,388 | 25,676,612 | 25,168,951 |
| LG50 (3.0 Gb) | 9 | 9 | 9 | 10 | 8 | 10 | 9 | 10 | 9 | 9 | 8 | 8 | 8 | 8 | 10 |
| LG90 (3.0 Gb) | 21 | 24 | 22 | 25 | 20 | 23 | 20 | 22 | 21 | 21 | 19 | 20 | 20 | 20 | 22 |
| LG99 (3.0 Gb) | 27 | 31 | 28 | 35 | 26 | 29 | 27 | 27 | 26 | 27 | 25 | 26 | 25 | 25 | 29 |
| Mean (bp) | 39,312,586 | 35,057,860 | 27,944,278 | 29,375,752 | 36,849,251 | 40,335,964 | 40,784,676 | 40,264,750 | 37,259,856 | 29,590,072 | 35,735,040 | 52,836,747 | 44,755,655 | 28,582,840 | 44,569,592 |
| Max (bp) | 242,149,584 | 242,859,312 | 241,076,672 | 201,236,838 | 242,149,755 | 202,383,115 | 242,162,689 | 202,730,645 | 242,584,874 | 242,571,736 | 248,541,081 | 248,076,779 | 247,915,876 | 242,583,836 | 218,917,483 |
| Min (bp) | 10,559 | 14,574 | 16,501 | 13,108 | 8,434 | 10,554 | 14,969 | 13,989 | 7,969 | 7,763 | 15,879 | 12,353 | 15,581 | 27,024 | 17,080 |
| T2T Chromosomes | 14 | 12 | 11 | 9 | 15 | 11 | 16 | 12 | 15 | 12 | 16 | 15 | 16 | 17 | 10 |
| Total Gaps | 13 | 20 | 19 | 26 | 14 | 19 | 11 | 15 | 13 | 14 | 8 | 12 | 11 | 10 | 26 |
| Total Gaps (exclude Y) | 11 | 16 | 15 | 21 | 9 | 15 | 8 | 13 | 9 | 12 | 7 | 9 | 9 | 7 | 20 |
| chr1 | 1 | 3 | 2 (Chimeric) | 2 | 0 | 1 | 2 | 1 | 1 | 1 | **0** | 0 | 0 | 0 | 2 (Chimeric) |
| chr2 | 0 | 0 | 0 | 2 | 0 | 2 | 0 | 1 | 0 | 0 | 0 | 0 | 0 | **0** | 2 (Chimeric) |
| chr3 | 0 | 0 | 0 | 0 | 0 | 0 | 0 | 1 | 0 | 0 | 0 | 0 | 0 | **0** | 0 |
| chr4 | 0 | 0 | 1 | 0 | 0 | 0 | 0 | 0 | 0 | 0 | 0 | 0 | 0 | **0** | 1 |
| chr5 | 0 | 0 | 0 | 0 | 0 | 1 | 0 | 0 | 0 | 0 | **0** | 1 | 0 | 2 | 1 |
| chr6 | 0 | 1 | 1 | 0 | 0 | 0 | 0 | 0 | 0 | 1 | 0 | 0 | 0 | **0** | 1 (Chimeric) |
| chr7 | 0 | 1 | 0 | 1 | 0 | 0 | 0 | 1 | 0 | 0 | 0 | 0 | 1 | **0** | 0 |
| chr8 | 0 | 0 | 0 | 0 | 0 | 0 | 1 | 0 | 0 | 1 | 1 | 0 | 0 | **0** | 0 |
| chr9 | 1 | 1 | 1 (Chimeric) | 2 | 1 | 1 | 0 | 1 | **0** | 1 | 1 | 2 | 0 | 1 | 2 |
| chr10 | 0 | 1 | 1 (Chimeric) | 1 | 0 | 0 | 0 | 0 | 0 | 0 | 0 | 0 | 0 | **0** | 0 |
| chr11 | 0 | 0 | 0 | 0 | 0 | 0 | 0 | 0 | 0 | 0 | 0 | 0 | 0 | **0** | 0 |
| chr12 | 0 | 0 | 0 | 0 | 0 | 0 | 0 | 0 | 0 | 0 | 0 | 0 | 0 | **0** | 0 |
| chr13 | 2 | 1 (Chimeric) | 1 | 1 | 1 | 2 | 1 (Chimeric) | 2 (Chimeric) | 0 | **0** | 1 | 1 | 1 | 1 | 2 (Chimeric) |
| chr14 | 1 | 1 (Chimeric) | 1 (Chimeric) | 1 | 2 | 1 | 0 | 1 | 1 | 1 | 1 | 1 | 2 (Chimeric) | 1 | **0** |
| chr15 | 1 | 2 | 1 | 1 | 1 | 1 | 0 | 1 (Chimeric) | 1 | 1 | 1 | 0 | 1 (Chimeric) | **0** | 2 (Chimeric) |
| chr16 | 1 | 3 | 1 | 2 | 1 | 1 | 1 | 1 | 1 | 1 | **0** | 1 | 1 | 1 | 2 |
| chr17 | 2 | 0 | 0 | 2 | 1 | 1 | 1 | 0 | 1 | 1 | 1 | 1 | **0** | 1 | 1 (Chimeric) |
| chr18 | 0 | 0 | 0 | 1 | 0 | 0 | 0 | 0 | 0 | 0 | 0 | 0 | 0 | **0** | 0 |
| chr19 | 0 | 0 | 0 | 0 | 0 | 0 | 0 | 0 | 0 | 0 | 0 | 0 | 0 | **0** | 0 |
| chr20 | 0 | 0 | 0 | 0 | 0 | 0 | 0 | 0 | 0 | 0 | 0 | 0 | 0 | **0** | 0 |
| chr21 | 1 | 1 | 2 (Chimeric) | 1 | 1 | 1 | 1* | 2 (Chimeric) | 1 | 1 | 0 | 1 | 1 (Chimeric) | **0** | 2 (Chimeric) |
| chr22 | 1 | 1 | 1 (Chimeric) | 2 | 1 | 1 | 1 | 1 (Chimeric) | 1 | 1 | 1 | 1 | 2 (Chimeric) | **0** | 1 |
| chrX | 0 | 0 | 2 | 2 | 0 | 2 | 0 | **0** | 2 | 2 | 0 | 0 | 0 | 0 | 1 |
| chrY | 2 | 4 | 4 | 5 | 5 | 4 | 3 | **2** | 4 | 2 | 1 | 3 | 2 | 3 | 6 |

→

| | |
|---|---|
| chr1 | 0 |
| chr2 | 0 |
| chr3 | 0 |
| chr4 | 0 |
| chr5 | 0 |
| chr6 | 0 |
| chr7 | 0 |
| chr8 | 0 |
| chr9 | 0 |
| chr10 | 0 |
| chr11 | 0 |
| chr12 | 0 |
| chr13 | 0 |
| chr14 | 0 |
| chr15 | 0 |
| chr16 | 0 |
| chr17 | 0 |
| chr18 | 0 |
| chr19 | 0 |
| chr20 | 0 |
| chr21 | 0 |
| chr22 | 0 |
| chrX | 0 |
| chrY | 2 |

H I J K M N F (2FC)

| | Composite Assembly ONT Contig (3 FC) | Composite Assembly ONT Scaffold (3 FC) |
|---|---|---|
| Assembly Size (bp) | 3,077,076,224 | 3,077,076,424 |
| Contig/ Scaffold Count | 26 | 24 |
| Complete Chromosomes | 23 | 23 |
| Gaps | 2 | 2 |
| NG50 (3.0 Gb) (bp) | 154,728,733 | 154,728,733 |
| NG90 (3.0 Gb) (bp) | 82,612,722 | 82,612,722 |
| NG99 (3.0 Gb) (bp) | 49,587,772 | 52,137,724 |
| LG50 (3.0 Gb) | 8 | 8 |
| LG90 (3.0 Gb) | 18 | 18 |
| LG99 (3.0 Gb) | 23 | 22 |
| Mean (bp) | 118,349,086 | 128,211,518 |
| Max (bp) | 248,541,081 | 248,541,081 |
| Min (bp) | 6,443,951 | 43,357,947 |
| Phase Block Count | 2,393 | |
| Phase Block NG50 (bp) | 2,360,941 | |

SI-5. Assembly Contiguity and Metrics for the 14 Sub-Assemblies (A-N) and the Final Composite-Assembly at Different Coverages. A final composite-assembly was created by selecting the best sub-assembly for each chromosome. In most cases, the sub-assembly with the least number of gaps for that chromosome was selected. In a few rare cases, the sub-assembly with an assembled chromosome length significantly longer than others was selected albeit having 1 or 2 more gaps. Overall, higher coverage generated more contiguous assemblies. Condition N proved to be suitable for most chromosomes at high-coverage. The final one-flowcell composite-assembly made use of 8 sub-assemblies, resulting in 62 gaps in total. The two-flowcell composite-assembly made use of 7 sub-assemblies, resulting in all but 2 gapless chromosomes, with the remaining 6 gaps residing on Chr15 and ChrY. The three-flowcell composite-assembly drew from 7 sub-assemblies, resulting in all chromosomes being gapless except for ChrY, which had 2 gaps.
