## Supplementary material for "The Revised Diploid Genome Sequence of an Individual Human: An Optimized Assembly Workflow for Scaling of near Telomere-to-Telomere Assemblies": SI_6.pdf

**QC/ Gap Filling**

→

Sub-assembly H: 2 Gaps on Y

Assembly Gap 1
351,481 bp

Assembly Gap 2
76,423 bp

### JCV-T2T Assembly

| | |
|---|---|
| chr1 | 0 |
| chr2 | 0 |
| chr3 | 0 |
| chr4 | 0 |
| chr5 | 0 |
| chr6 | 0 |
| chr7 | 0 |
| chr8 | 0 |
| chr9 | 0 |
| chr10 | 0 |
| chr11 | 0 |
| chr12 | 0 |
| chr13 | 0 |
| chr14 | 0 |
| chr15 | 0 |
| chr16 | 0 |
| chr17 | 0 |
| chr18 | 0 |
| chr19 | 0 |
| chr20 | 0 |
| chr21 | 0 |
| chr22 | 0 |
| chrX | 0 |
| chrY | 0 |

| | T2T-Assembly JCV |
|---|---|
| Assembly Size (bp) | 3,077,495,244 |
| Contig/ Scaffold Count | **24** |
| Complete Chromosomes | **24** |
| Gaps | **0** |
| NG50 (3.0 Gb) (bp) | 154,728,740 |
| NG90 (3.0 Gb) (bp) | 82,612,577 |
| NG99 (3.0 Gb) (bp) | 52,565,427 |
| LG50 (3.0 Gb) (bp) | 8 |
| LG90 (3.0 Gb) (bp) | 18 |
| LG99 (3.0 Gb) (bp) | 22 |
| Mean (bp) | 128,228,968 |
| Max (bp) | 248,539,509 |
| Min (bp) | 43,357,947 |
| Phase Block Count | **2,393** |
| Phase Block NG50 (bp) | **2,360,941** |

SI-6. Metrics of the Three-Flowcell Composite-Assembly and the Subsequent Gapless JCV-T2T Assembly. The composite-assembly comprised the best chromosomes from sub-assemblies H, I, J, K, M, N, and F, with only two gaps on ChrY. The two gaps were filled by manual curation to produce the JCV-T2T assembly, with a contig NG50 (3.0 Gb) of 154.7 Mb.
