## Supplementary material for "The Revised Diploid Genome Sequence of an Individual Human: An Optimized Assembly Workflow for Scaling of near Telomere-to-Telomere Assemblies": SI_7.pdf

### SI-7: Homopolymer Tracks in Composite-Assembly

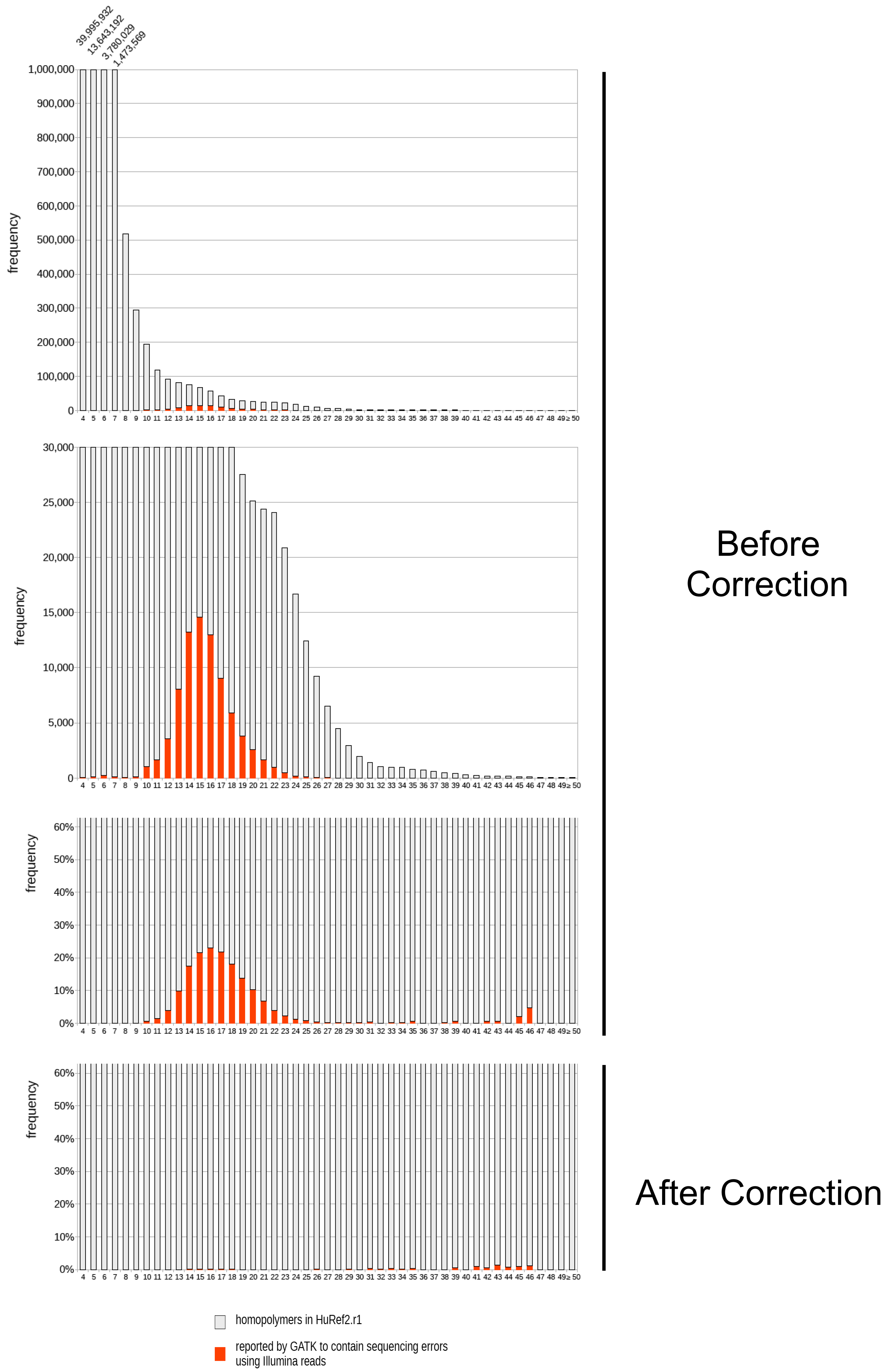


SI-7. Homopolymer Track Length Distribution in HuRef2.0 Before and After Correction. Red bars represent the portion of homopolymer tracks containing sequencing errors reported by GATK using Illumina reads.
