## Supplementary material for "The Revised Diploid Genome Sequence of an Individual Human: An Optimized Assembly Workflow for Scaling of near Telomere-to-Telomere Assemblies": SI_8.pdf

### SI-8: Venter Mitochondria (16,569 bp) : Haplogroup K1a3a (95% hit)

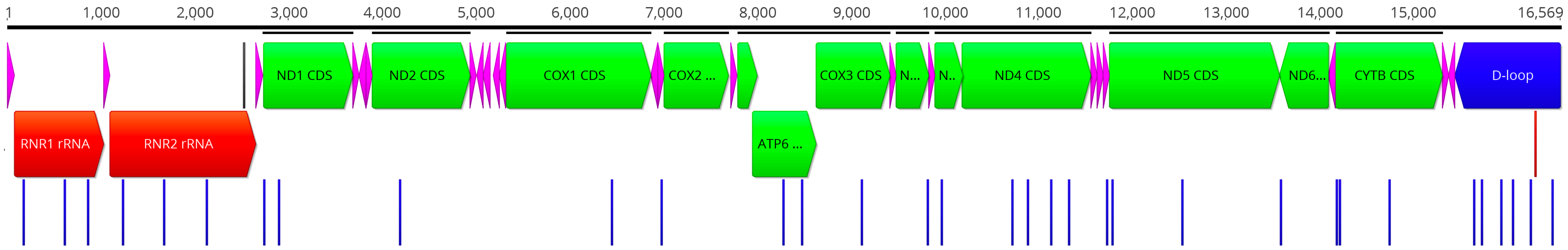


#### 34 Variants (1 insertion; 33 Transitions)

| rCRS Position | rCRS NT | Query NT | Mut type | Locus | Other |
|---|---|---|---|---|---|
| 73 | A | G | transition | CR:HVS2ATTCR: | non-coding |
| 263 | A | G | transition | CR:HVS2CR:OH/ | non-coding |
| 315 | C | CC | insertion | CR:HVS2CR:OH/ | non-coding |
| 497 | C | T | transition | ATTCR:Control I | non-coding |
| 750 | A | G | transition | 12S | rRNA |
| 1189 | T | C | transition | 12S | rRNA |
| 1438 | A | G | transition | 12S | rRNA |
| 1811 | A | G | transition | 16SSHLP3 | rRNA |
| 2251 | A | G | transition | 16S | rRNA |
| 2706 | A | G | transition | 16SHumanin | rRNA |
| 3316 | G | A | transition | ND1 | ND1:A4T |
| 3480 | A | G | transition | ND1 | ND1:K58K |
| 4769 | A | G | transition | ND2 | ND2:M100M |
| 7028 | C | T | transition | COI | COI:A375A |
| 7559 | A | G | transition | D | MitoTIP7.70% |
| 8860 | A | G | transition | ATPase6 | ATPase6:T112A |
| 9055 | G | A | transition | ATPase6 | ATPase6:A177T |
| 9698 | T | C | transition | COIII | COIII:L164L |
| 10398 | A | G | transition | ND3 | ND3:T114A |
| 10550 | A | G | transition | ND4L | ND4L:M27M |
| 11299 | T | C | transition | ND4 | ND4:T180T |
| 11467 | A | G | transition | ND4 | ND4:L236L |
| 11719 | G | A | transition | ND4 | ND4:G320G |
| 11902 | G | A | transition | ND4 | ND4:V381V |
| 12308 | A | G | transition | L(CUN) | MitoTIP42.00% |
| 12372 | G | A | transition | ND5 | ND5:L12L |
| 13117 | A | G | transition | ND5 | ND5:I261V |
| 14167 | C | T | transition | ND6 | ND6:E169E |
| 14766 | C | T | transition | Cytb | Cytb:T7I |
| 14798 | T | C | transition | Cytb | Cytb:F18L |
| 15326 | A | G | transition | Cytb | Cytb:T194A |
| 16224 | T | C | transition | ATTCR:Control I | non-coding |
| 16311 | T | C | transition | ATTCR:Control I | non-coding |
| 16519 | T | C | transition | ATTCR:Control I | non-coding |

##### JCVenter_MT

**Top Hit**

K1a3a (95%)

**Expected Mutations**

73G 263G 497T 750G 1189C 1438G 1811G 2706G 3480G 4769G 7028T 7559G 8860G 9055A 9698C 10398G 10550G 11299C 11467G 11719A 12308G 12372A 13117G 14167T 14766T 14798C 15326G 16093C 16224C 16311C

**Remaining Mutations**

315.1C 2251G 3107d 3316A 11902A 16519C

**Additional Hits**

**Ranges**

577-16569 1-576

**Expected But Not Included** 73G!

Mutations displayed in red are mutations that are **expected by the tophit but not included** in the input sample.

**Remaining Mutations**

Remaining mutations of the input sample are separated by its type.

**Hotspots** 315.1C

Hotspots mutations are defined by a high number of occurrences in the used phylogenetic tree.

**Local Private Mutations** 73G

Local private mutations are mutations that are not associated with the tophit but are included in the phylogenetic tree for other haplogroups.


Haplotype assignment:
Lott, M.T., Leipzig, J.N., Derbeneva, O., Xie, H.M., Chalkia, D., Sarmady, M., Procaccio, V., and Wallace, D.C. 2013. mtDNA variation and analysis using MITOMAP and MITOMASTER. Current Protocols in Bioinformatics 1(123):1.23.1-26. PMID: 25489354 URL: http://www.mitomap.org

Haplotype map: https://blog.23andme.com/articles/haplogroups-explained
