## Supplementary material for "The Revised Diploid Genome Sequence of an Individual Human: An Optimized Assembly Workflow for Scaling of near Telomere-to-Telomere Assemblies": SI_9.pdf

### SI-9: HuRef2.0 Phase Blocks and Switch-Over Regions

(a) HuRef2.0 Phase Blocks

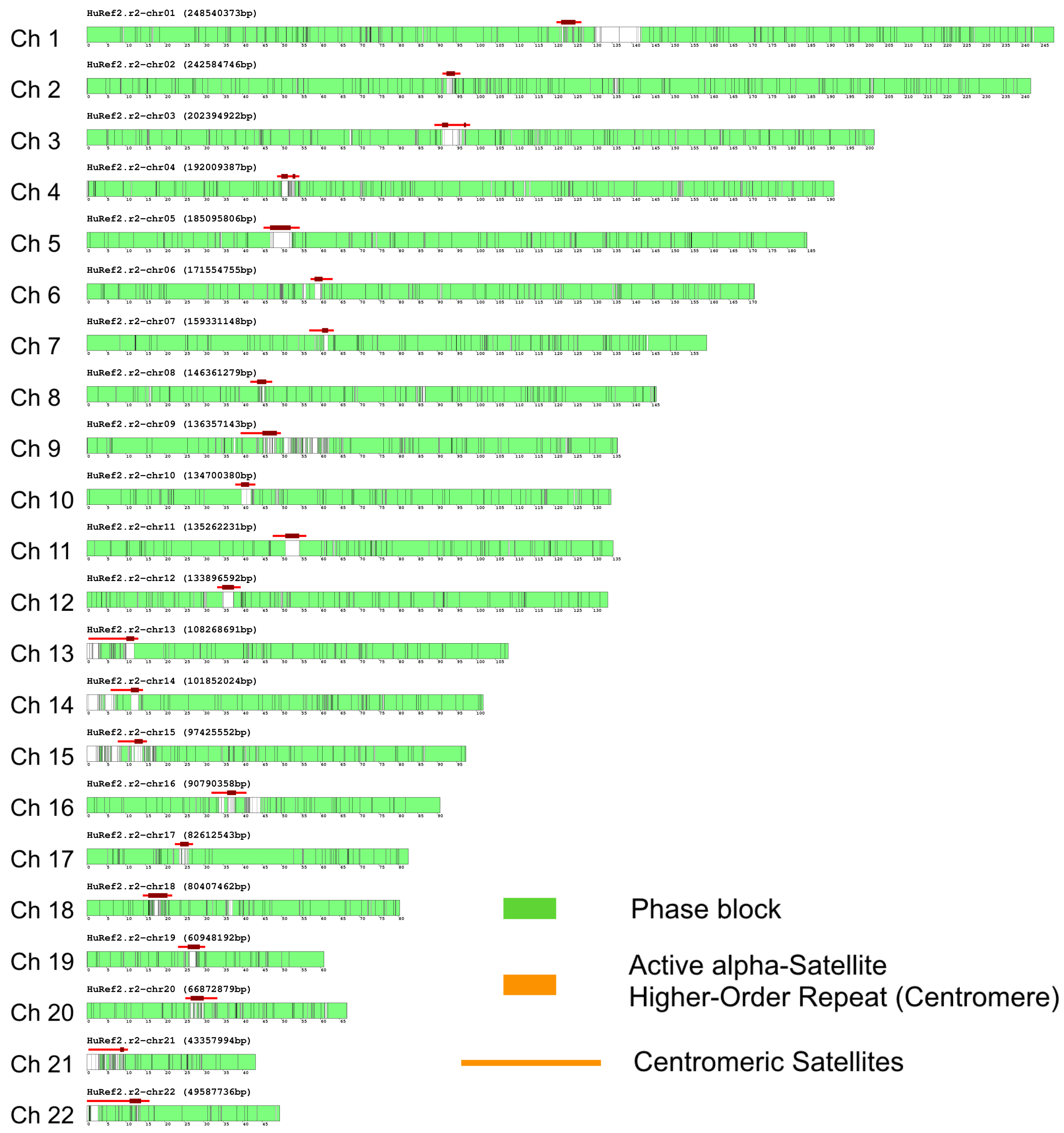


(b) HuRef2.0 Switch-Over Regions

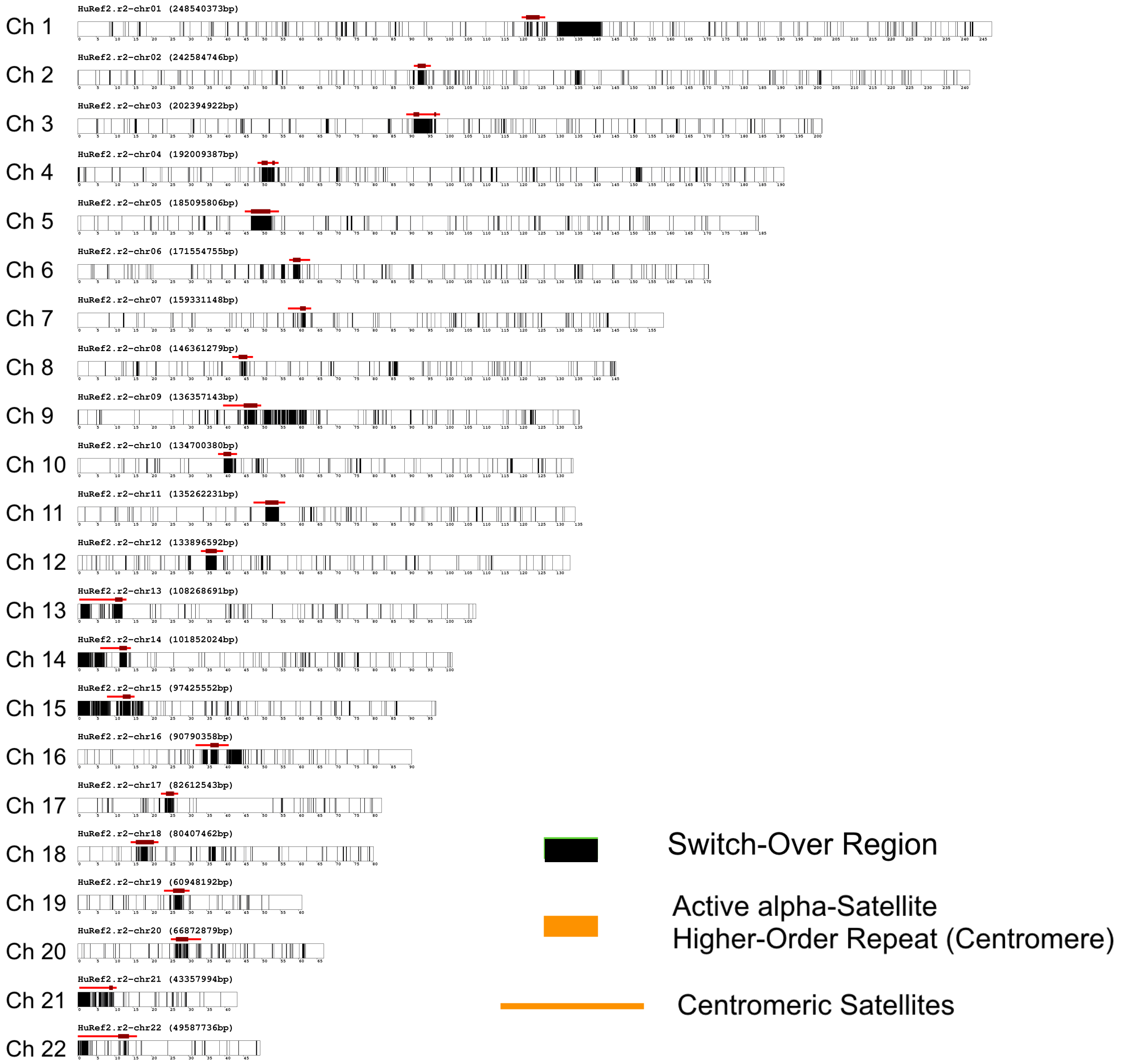


Phase Blocks:
Count = 2,393
NG50 (3.0 Gb) = 2.36 Mbp
Combined Length = 2,615,977,996 bp
Proportion of Autosomes = 91.1%

Switch-Over Regions:
Count = 2,371
N50 = 122,756 bp
Combined Length = 253,065,573 bp
Proportion of Autosomes = 8.8%

SI-9. Phase Block Boundaries for HuRef2.0 Autosomes. (a) Assembly Phased by Aligning ONT SUP-Reads. Heterozygous sites were detected and phased with the aligned reads using WhatsHap[40]. Phase blocks are highlighted in green. Centromeric satellites and active alpha-satellite higher-order-repeats are demarcated using CenMAPv1.2.0[24], and are denoted by red lines and deep red thicker lines, respectively. (b) Switch-Over Regions Between Phase Blocks. Switch-over regions are highlighted in black.
