## Supplementary material for "The Revised Diploid Genome Sequence of an Individual Human: An Optimized Assembly Workflow for Scaling of near Telomere-to-Telomere Assemblies": SI_10.pdf

Chromosome Y Gene Families

| Family | Gene | Chromosome | CDS_Start | CDS_End | Strand |
|---|---|---|---|---|---|
| *BPY2* | *BPY2A* | HuRef2.0-chrY | 7,634,488 | 7,639,131 | - |
| *BPY2* | *BPY2B* | HuRef2.0-chrY | 23,977,119 | 23,983,043 | - |
| *BPY2* | *BPY2C* | HuRef2.0-chrY | 25,405,276 | 25,411,201 | + |
| *BPY2* | *BPY2D* | HuRef2.0-chrY | 25,824,371 | 25,830,295 | - |
| *CDY* | *CDY1* | HuRef2.0-chrY | 18,557,304 | 18,558,929 | - |
| *CDY* | *CDY2* | HuRef2.0-chrY | 18,705,162 | 18,706,787 | + |
| *CDY* | *CDY3* | HuRef2.0-chrY | 24,825,254 | 24,826,876 | - |
| *CDY* | *CDY4* | HuRef2.0-chrY | 26,408,645 | 26,410,267 | + |
| *DAZ* | *DAZ1* | HuRef2.0-chrY | 23,689,946 | 23,755,907 | - |
| *DAZ* | *DAZ2* | HuRef2.0-chrY | 23,776,433 | 23,840,222 | + |
| *DAZ* | *DAZ3* | HuRef2.0-chrY | 25,548,096 | 25,597,330 | - |
| *DAZ* | *DAZ4* | HuRef2.0-chrY | 25,617,856 | 25,687,474 | + |
| *HSFY* | *HSFY1* | HuRef2.0-chrY | 19,225,768 | 19,227,451 | + |
| *HSFY* | *HSFY2* | HuRef2.0-chrY | 19,410,733 | 19,417,371 | - |
| *HSFY* | *HSFY3* | HuRef2.0-chrY | 19,450,915 | 19,452,598 | - |
| *PRY* | *PRY1* | HuRef2.0-chrY | 22,850,940 | 22,875,176 | - |
| *PRY* | *PRY2* | HuRef2.0-chrY | 23,280,812 | 23,293,981 | + |
| *RBMY* | *RBMY1* | HuRef2.0-chrY | 22,308,701 | 22,320,858 | + |
| *RBMY* | *RBMY2* | HuRef2.0-chrY | 22,332,241 | 22,344,398 | + |
| *RBMY* | *RBMY3* | HuRef2.0-chrY | 22,355,781 | 22,367,937 | + |
| *RBMY* | *RBMY4* | HuRef2.0-chrY | 22,659,541 | 22,671,699 | - |
| *RBMY* | *RBMY5* | HuRef2.0-chrY | 22,683,082 | 22,695,238 | - |
| *RBMY* | *RBMY6* | HuRef2.0-chrY | 22,947,975 | 22,960,127 | - |
| *RBMY* | *RBMY7* | HuRef2.0-chrY | 23,184,731 | 23,196,880 | + |
| *TSPY* | *TSPY* | HuRef2.0-chrY | 5,947,222 | 5,949,778 | + |
| *TSPY* | *TSPY1* | HuRef2.0-chrY | 8,989,081 | 8,991,643 | + |
| *TSPY* | *TSPY2* | HuRef2.0-chrY | 9,009,393 | 9,011,955 | + |
| *TSPY* | *TSPY3* | HuRef2.0-chrY | 9,029,717 | 9,032,297 | + |
| *TSPY* | *TSPY4* | HuRef2.0-chrY | 9,050,010 | 9,052,572 | + |
| *TSPY* | *TSPY6* | HuRef2.0-chrY | 9,090,626 | 9,093,188 | + |
| *TSPY* | *TSPY5* | HuRef2.0-chrY | 9,070,317 | 9,072,897 | + |
| *TSPY* | *TSPY7* | HuRef2.0-chrY | 9,110,922 | 9,113,484 | + |
| *TSPY* | *TSPY8* | HuRef2.0-chrY | 9,131,203 | 9,133,765 | + |
| *TSPY* | *TSPY9* | HuRef2.0-chrY | 9,151,482 | 9,154,044 | + |
| *TSPY* | *TSPY10* | HuRef2.0-chrY | 9,171,771 | 9,174,333 | + |
| *TSPY* | *TSPY11* | HuRef2.0-chrY | 9,192,034 | 9,194,614 | + |
| *TSPY* | *TSPY12* | HuRef2.0-chrY | 9,212,352 | 9,214,932 | + |
| *TSPY* | *TSPY13* | HuRef2.0-chrY | 9,232,682 | 9,235,244 | + |
| *TSPY* | *TSPY15* | HuRef2.0-chrY | 9,273,301 | 9,275,881 | + |
| *TSPY* | *TSPY16* | HuRef2.0-chrY | 9,293,621 | 9,296,201 | + |
| *TSPY* | *TSPY17* | HuRef2.0-chrY | 9,313,926 | 9,316,488 | + |
| *TSPY* | *TSPY18* | HuRef2.0-chrY | 9,334,203 | 9,336,765 | + |
| *TSPY* | *TSPY19* | HuRef2.0-chrY | 9,354,511 | 9,357,073 | + |
| *TSPY* | *TSPY20* | HuRef2.0-chrY | 9,374,835 | 9,377,397 | + |
| *TSPY* | *TSPY21* | HuRef2.0-chrY | 9,395,124 | 9,397,686 | + |
| *TSPY* | *TSPY22* | HuRef2.0-chrY | 9,415,406 | 9,417,968 | + |
| *TSPY* | *TSPY23* | HuRef2.0-chrY | 9,435,698 | 9,438,260 | + |
| *TSPY* | *TSPY24* | HuRef2.0-chrY | 9,456,015 | 9,458,577 | + |
| *TSPY* | *TSPY25* | HuRef2.0-chrY | 9,476,296 | 9,478,876 | + |
| *TSPY* | *TSPY26* | HuRef2.0-chrY | 9,496,595 | 9,499,175 | + |
| *TSPY* | *TSPY27* | HuRef2.0-chrY | 9,516,868 | 9,519,448 | + |
| *TSPY* | *TSPY28* | HuRef2.0-chrY | 9,537,170 | 9,539,732 | + |
| *TSPY* | *TSPY29* | HuRef2.0-chrY | 9,557,449 | 9,560,027 | + |
| *VCY* | *VCY1* | HuRef2.0-chrY | 14,665,060 | 14,665,629 | - |
| *VCY* | *VCY2* | HuRef2.0-chrY | 14,735,457 | 14,736,026 | + |
| *XKRY* | *XKRYP1* | HuRef2.0-chrY | 18,448,014 | 18,449,594 | - |
| *XKRY* | *XKRYP2* | HuRef2.0-chrY | 18,814,504 | 18,816,084 | + |
| *XKRY* | *XKRYP3* | HuRef2.0-chrY | 19,186,795 | 19,188,591 | + |
| *XKRY* | *XKRYP4* | HuRef2.0-chrY | 19,489,778 | 19,491,574 | - |
| *XKRY* | *XKRYP5* | HuRef2.0-chrY | 24,494,873 | 24,496,671 | - |
| *XKRY* | *XKRYP6* | HuRef2.0-chrY | 24,695,878 | 24,697,676 | - |
| *XKRY* | *XKRYP7* | HuRef2.0-chrY | 26,537,823 | 26,539,621 | + |
| *XKRY* | *XKRYP8* | HuRef2.0-chrY | 26,738,806 | 26,740,603 | + |

SI-10. Ampliconic Gene Clusters on ChrY.
