## Supplementary material for "The Revised Diploid Genome Sequence of an Individual Human: An Optimized Assembly Workflow for Scaling of near Telomere-to-Telomere Assemblies": SI_11.pdf

(a)

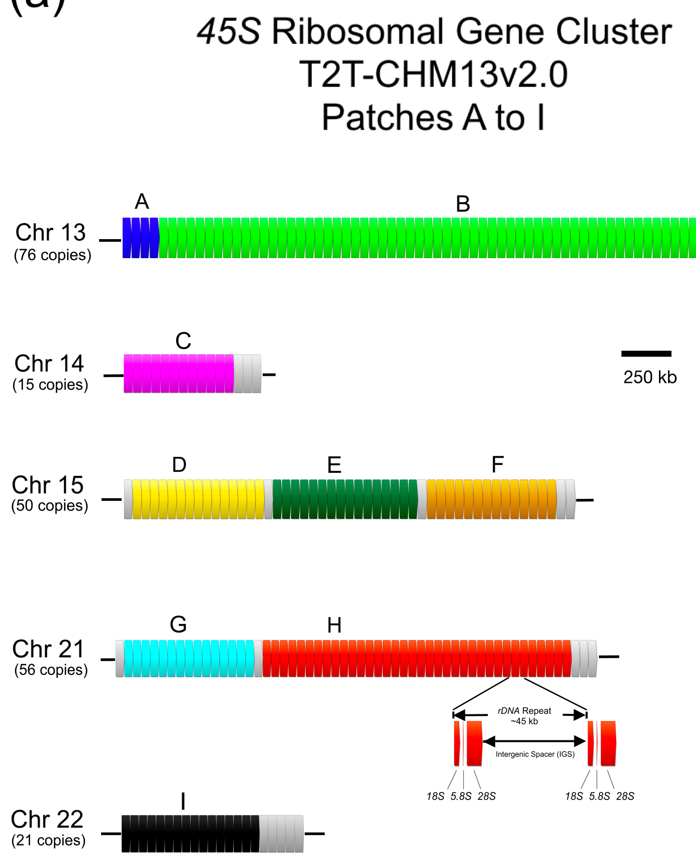


(b)

Sequence Identity of Patches (%)

| | Chr 13 ribo-A | Chr 13 ribo-B | Chr 14 ribo-C | Chr 15 ribo-D | Chr 15 ribo-E | Chr 15 ribo-F | Chr 21 ribo-G | Chr 21 ribo-H | Chr 22 ribo-I |
|---|---|---|---|---|---|---|---|---|---|
| Chr 13 ribo-A | | 99.998 | 93.072 | 89.451 | 92.524 | 94.513 | 92.774 | 94.476 | 91.433 |
| Chr 13 ribo-B | 99.998 | | 93.074 | 89.454 | 92.526 | 94.515 | 92.776 | 94.478 | 91.435 |
| Chr 14 ribo-C | 93.072 | 93.074 | | 95.363 | 95.393 | 97.624 | 95.515 | 97.388 | 97.16 |
| Chr 15 ribo-D | 89.451 | 89.454 | 95.363 | | 96.355 | 94.082 | 92.041 | 93.402 | 96.545 |
| Chr 15 ribo-E | 92.524 | 92.526 | 95.393 | 96.355 | | 97.605 | 95.19 | 96.566 | 93.298 |
| Chr 15 ribo-F | 94.513 | 94.515 | 97.624 | 94.082 | 97.605 | | 96.875 | 98.697 | 95.489 |
| Chr 21 ribo-G | 92.774 | 92.776 | 95.515 | 92.041 | 95.19 | 96.875 | | 97.537 | 93.478 |
| Chr 21 ribo-H | 94.476 | 94.478 | 97.388 | 93.402 | 96.566 | 98.697 | 97.537 | | 95.28 |
| Chr 22 ribo-I | 91.433 | 91.435 | 97.16 | 96.545 | 93.298 | 95.489 | 93.478 | 95.28 | |

(c)

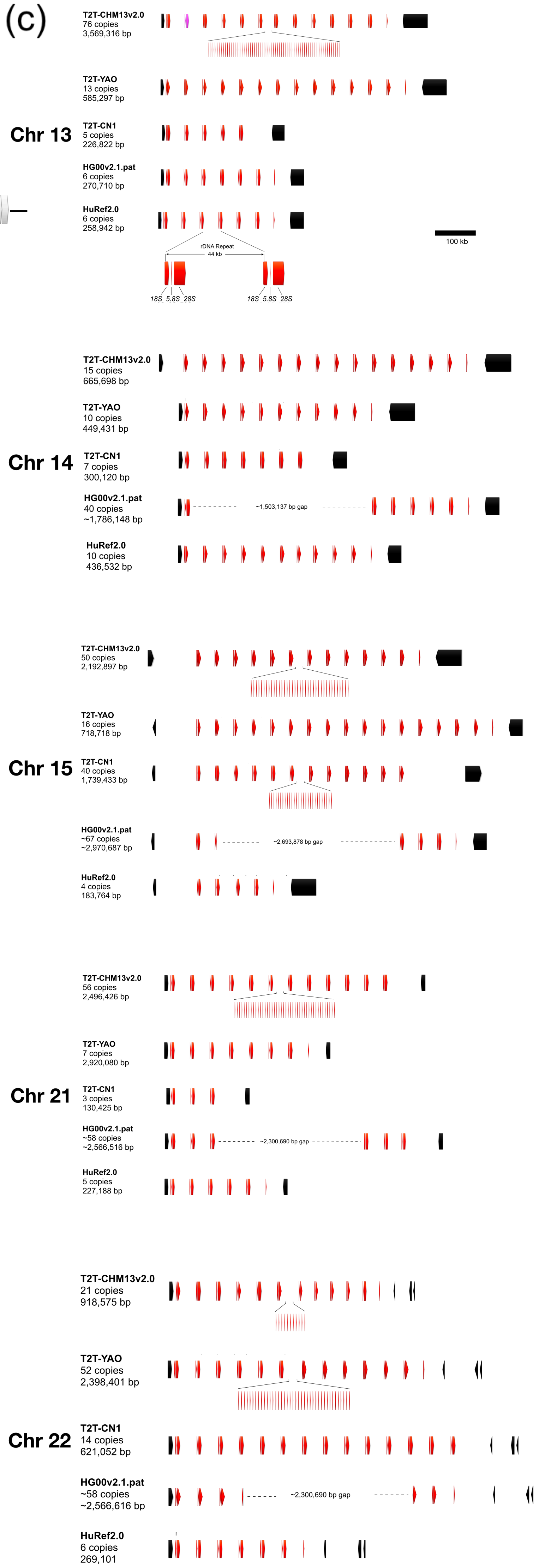

SI-11: *45S* Ribosomal Gene Clusters in T2T-Assemblies. (a) *45S* Ribosomal Gene Clusters in T2T-CHM13v2.0. The nine *45S* patches used to reconstruct the cluster in T2T-CHM13v2.0 in accordance to scaffolding information are designated “A” to “I” and are shaded in the colour as indicated. (b) Sequence identity of the nine patches. (c) The *45S* Ribosomal Gene Clusters in the Recent T2T-Assemblies[9-12].
