## Supplementary material for "The Revised Diploid Genome Sequence of an Individual Human: An Optimized Assembly Workflow for Scaling of near Telomere-to-Telomere Assemblies": SI_12.pdf

SI-12. Three Hundred and Eighty-Two Rare OMIM Phenotype-Associated Coding Variants (SNV/indels). The variants listed are rare (<3% population frequency) and occur in the CDS of OMIM phenotype-associated genes. This list corresponds to the blue numbers in the Venn diagram in Figure 4a, with the calling method column denoting which Venn diagram region the variant belongs to.

| Gene | Calling method (V1=Illumina short reads, V2= ONT long reads, V3=assembly, V4=HiFi long reads) | Location (GRCh38) | Consequence | MANE transcript | Genomic change | Transcript / Protein Change | Zygosity | OMIM Phenotype |
|---|---|---|---|---|---|---|---|---|
| *C1orf127* | V1,V2,V3,V4 | chr1:10949633 | synonymous_variant | NM_001170754.2 | g.10949633T>C | c.1281A>G (p.Ala427Ala) | heterozygous | Heterotaxy, visceral, 14, autosomal, 621080 (3) |
| *EPHA2* | V1,V2,V3,V4 | chr1:16148487 | synonymous_variant | NM_004431.5 | g.16148487C>G | c.714G>C (p.Pro238Pro) | heterozygous | Cataract 6, multiple types, 116600 (3) |
| *RHD* | V1,V2,V4 | chr1:25321889 | missense_variant,splice_region_variant | NM_016124.6 | g.25321889G>C | c.1154G>C (p.Gly385Ala) | heterozygous | {Hemolytic disease of fetus and newborn, RH-induced}, 619462 (3) |
| *RHCE* | V1,V2,V3,V4 | chr1:25408711 | missense_variant | NM_020485.8 | g.25408711G>A | c.307C>T (p.Pro103Ser) | heterozygous | Rh-null disease, amorph type, 617970 (3) |
| *RHCE* | V1,V2,V3,V4 | chr1:25408815 | missense_variant | NM_020485.8 | g.25408815T>C | c.203A>G (p.Asn68Ser) | heterozygous | Rh-null disease, amorph type, 617970 (3) |
| *RHCE* | V1,V2,V3,V4 | chr1:25408817 | synonymous_variant | NM_020485.8 | g.25408817T>C | c.201A>G (p.Ser67Ser) | heterozygous | Rh-null disease, amorph type, 617970 (3) |
| *RHCE* | V1,V2,V3,V4 | chr1:25408840 | missense_variant | NM_020485.8 | g.25408840G>T | c.178C>A (p.Leu60Ile) | heterozygous | Rh-null disease, amorph type, 617970 (3) |
| *RHCE* | V1,V2,V3,V4 | chr1:25408868 | splice_region_variant,synonymous_variant | NM_020485.8 | g.25408868G>A | c.150C>T (p.Val50Val) | heterozygous | Rh-null disease, amorph type, 617970 (3) |
| *SZT2* | V1,V2,V3,V4 | chr1:43448343 | synonymous_variant | NM_001365999.1 | g.43448343C>T | c.9828C>T (p.Asp3276Asp) | heterozygous | Developmental and epileptic encephalopathy 18, 615476 (3) |
| *HFM1* | V1,V2,V3,V4 | chr1:91394241 | missense_variant | NM_001017975.6 | g.91394241G>A | c.346C>T (p.His116Tyr) | heterozygous | Premature ovarian failure 9, 615724 (3) |
| *RPL5* | V1,V2,V3,V4 | chr1:92837557 | missense_variant | NM_000969.5 | g.92837557A>G | c.629A>G (p.Tyr210Cys) | heterozygous | Diamond-Blackfan anemia 6, 612561 (3) |
| *SPAG17* | V1,V2,V3,V4 | chr1:118041837 | missense_variant | NM_206996.4 | g.118041837G>C | c.3020C>G (p.Ser1007Cys) | heterozygous | ?Spermatogenic failure 55, 619380 (3) |
| *FLG* | V1,V2,V4 | chr1:152304286 | missense_variant | NM_002016.2 | g.152304286T>A | c.10600A>T (p.Asn3534Tyr) | heterozygous | {Dermatitis, atopic, susceptibility to, 2}, 605803 (3) |
| *FLG* | V1,V2,V4 | chr1:152304306 | missense_variant | NM_002016.2 | g.152304306G>T | c.10580C>A (p.Ala3527Glu) | heterozygous | {Dermatitis, atopic, susceptibility to, 2}, 605803 (3) |
| *FLG* | V1,V2,V4 | chr1:152304321 | missense_variant | NM_002016.2 | g.152304321C>A | c.10565G>T (p.Gly3522Val) | heterozygous | {Dermatitis, atopic, susceptibility to, 2}, 605803 (3) |
| *FLG* | V1,V2,V4 | chr1:152304323 | synonymous_variant | NM_002016.2 | g.152304323G>T | c.10563C>A (p.Ser3521Ser) | heterozygous | {Dermatitis, atopic, susceptibility to, 2}, 605803 (3) |
| *FLG* | V1,V2,V4 | chr1:152304325 | missense_variant | NM_002016.2 | g.152304325A>C | c.10561T>G (p.Ser3521Ala) | heterozygous | {Dermatitis, atopic, susceptibility to, 2}, 605803 (3) |
| *FLG* | V1 | chr1:152304368 | synonymous_variant | NM_002016.2 | g.152304368G>A | c.10518C>T (p.His3506His) | heterozygous | {Dermatitis, atopic, susceptibility to, 2}, 605803 (3) |
| *FLG* | V1,V2 | chr1:152304372 | missense_variant | NM_002016.2 | g.152304372T>C | c.10514A>G (p.His3505Arg) | heterozygous | {Dermatitis, atopic, susceptibility to, 2}, 605803 (3) |
| *FLG* | V1,V2 | chr1:152304379 | missense_variant | NM_002016.2 | g.152304379A>C | c.10507T>G (p.Trp3503Gly) | heterozygous | {Dermatitis, atopic, susceptibility to, 2}, 605803 (3) |
| *FLG* | V1,V2,V4 | chr1:152304389 | synonymous_variant | NM_002016.2 | g.152304389G>A | c.10497C>T (p.Ser3499Ser) | heterozygous | {Dermatitis, atopic, susceptibility to, 2}, 605803 (3) |
| *FLG* | V1,V2 | chr1:152304409-152304410 | inframe_insertion | NM_002016.2 | g.152304409_152304410insCTT | c.10476_10477insAAG (p.3492_3493insLys) | heterozygous | {Dermatitis, atopic, susceptibility to, 2}, 605803 (3) |
| *FLG* | V4 | chr1:152304410 | missense_variant | NM_002016.2 | g.152304410G>C | c.10476C>G (p.Asp3492Glu) | heterozygous | {Dermatitis, atopic, susceptibility to, 2}, 605803 (3) |
| *FLG* | V4 | chr1:152304412 | missense_variant | NM_002016.2 | g.152304412C>T | c.10474G>A (p.Asp3492Asn) | heterozygous | {Dermatitis, atopic, susceptibility to, 2}, 605803 (3) |
| *FLG* | V1,V2 | chr1:152304415-152304417 | inframe_deletion | NM_002016.2 | g.152304415_152304417del | c.10469_10471del (p.3490_3491delinsHis) | heterozygous | {Dermatitis, atopic, susceptibility to, 2}, 605803 (3) |
| *FLG* | V4 | chr1:152304417 | missense_variant | NM_002016.2 | g.152304417C>T | c.10469G>A (p.Arg3490His) | heterozygous | {Dermatitis, atopic, susceptibility to, 2}, 605803 (3) |
| *FLG* | V1,V2,V4 | chr1:152304419 | synonymous_variant | NM_002016.2 | g.152304419A>T | c.10467T>A (p.Thr3489Thr) | heterozygous | {Dermatitis, atopic, susceptibility to, 2}, 605803 (3) |
| *FLG* | V1,V2,V4 | chr1:152304431 | synonymous_variant | NM_002016.2 | g.152304431G>T | c.10455C>A (p.Ala3485Ala) | heterozygous | {Dermatitis, atopic, susceptibility to, 2}, 605803 (3) |
| *FLG* | V1,V2,V4 | chr1:152304538 | missense_variant | NM_002016.2 | g.152304538A>G | c.10348T>C (p.Tyr3450His) | heterozygous | {Dermatitis, atopic, susceptibility to, 2}, 605803 (3) |
| *FLG* | V1,V2,V4 | chr1:152304545 | synonymous_variant | NM_002016.2 | g.152304545T>C | c.10341A>G (p.Gly3447Gly) | heterozygous | {Dermatitis, atopic, susceptibility to, 2}, 605803 (3) |
| *FLG* | V1,V2,V4 | chr1:152304551 | synonymous_variant | NM_002016.2 | g.152304551C>T | c.10335G>A (p.Arg3445Arg) | heterozygous | {Dermatitis, atopic, susceptibility to, 2}, 605803 (3) |
| *FLG* | V2,V4 | chr1:152304559 | missense_variant | NM_002016.2 | g.152304559T>C | c.10327A>G (p.Arg3443Gly) | heterozygous | {Dermatitis, atopic, susceptibility to, 2}, 605803 (3) |
| *FLG* | V1,V2,V4 | chr1:152304568 | missense_variant | NM_002016.2 | g.152304568A>G | c.10318T>C (p.Ser3440Pro) | heterozygous | {Dermatitis, atopic, susceptibility to, 2}, 605803 (3) |
| *FLG* | V1,V2,V4 | chr1:152304645 | missense_variant | NM_002016.2 | g.152304645T>C | c.10241A>G (p.His3414Arg) | heterozygous | {Dermatitis, atopic, susceptibility to, 2}, 605803 (3) |
| *FLG* | V1,V2,V4 | chr1:152304652 | missense_variant | NM_002016.2 | g.152304652C>T | c.10234G>A (p.Gly3412Arg) | heterozygous | {Dermatitis, atopic, susceptibility to, 2}, 605803 (3) |
| *FLG* | V1,V4 | chr1:152304661 | missense_variant | NM_002016.2 | g.152304661G>C | c.10225C>G (p.Arg3409Gly) | heterozygous | {Dermatitis, atopic, susceptibility to, 2}, 605803 (3) |
| *FLG* | V1,V4 | chr1:152304673 | missense_variant | NM_002016.2 | g.152304673T>G | c.10213A>C (p.Thr3405Pro) | heterozygous | {Dermatitis, atopic, susceptibility to, 2}, 605803 (3) |
| *FLG* | V1,V4 | chr1:152304676 | missense_variant | NM_002016.2 | g.152304676T>C | c.10210A>G (p.Arg3404Gly) | heterozygous | {Dermatitis, atopic, susceptibility to, 2}, 605803 (3) |
| *FLG* | V1,V4 | chr1:152304677 | synonymous_variant | NM_002016.2 | g.152304677G>T | c.10209C>A (p.Thr3403Thr) | heterozygous | {Dermatitis, atopic, susceptibility to, 2}, 605803 (3) |
| *FLG* | V1,V2,V4 | chr1:152304680 | synonymous_variant | NM_002016.2 | g.152304680C>G | c.10206G>C (p.Arg3402Arg) | heterozygous | {Dermatitis, atopic, susceptibility to, 2}, 605803 (3) |
| *FLG* | V2,V4 | chr1:152304683 | synonymous_variant | NM_002016.2 | g.152304683C>T | c.10203G>A (p.Gly3401Gly) | heterozygous | {Dermatitis, atopic, susceptibility to, 2}, 605803 (3) |
| *FLG* | V1,V2,V4 | chr1:152304833 | synonymous_variant | NM_002016.2 | g.152304833G>A | c.10053C>T (p.Asp3351Asp) | heterozygous | {Dermatitis, atopic, susceptibility to, 2}, 605803 (3) |
| *FLG* | V1,V2,V4 | chr1:152305276 | missense_variant | NM_002016.2 | g.152305276C>T | c.9610G>A (p.Gly3204Arg) | heterozygous | {Dermatitis, atopic, susceptibility to, 2}, 605803 (3) |
| *FLG* | V1,V2,V3,V4 | chr1:152310830 | missense_variant | NM_002016.2 | g.152310830A>T | c.4056T>A (p.Ser1352Arg) | heterozygous | {Dermatitis, atopic, susceptibility to, 2}, 605803 (3) |
| *PPOX* | V1,V2,V3,V4 | chr1:161169143 | missense_variant | NM_001122764.3 | g.161169143C>G | c.767C>G (p.Pro256Arg) | heterozygous | Variegate porphyria, childhood-onset, 620483 (3) |
| *HMCN1* | V1,V2,V4 | chr1:186182258 | missense_variant | NM_031935.3 | g.186182258A>G | c.16385A>G (p.Gln5462Arg) | heterozygous | {Macular degeneration, age-related, 1}, 603075 (3) |
| *PRG4* | V1,V2,V3,V4 | chr1:186307343-186307344 | inframe_insertion | NM_005807.6 | g.186307343_186307344insCCAAGTCTGCACCCACCACTC | c.1624_1625insCCAAGTCTGCACCCACCACTC (p.Thr542delinsThrLysSerAlaProThrThrPro) | heterozygous | Camptodactyly-arthropathy-coxa vara-pericarditis syndrome, 208250 (3) |
| *CR1* | V4 | chr1:207545375 | synonymous_variant | NM_000651.6 | g.207545375C>T | c.2304C>T (p.Pro768Pro) | heterozygous | {Malaria, severe, resistance to}, 611162 (3) |
| *CR1* | V1,V2,V4 | chr1:207560952 | missense_variant | NM_000651.6 | g.207560952T>C | c.3401T>C (p.Ile1134Thr) | heterozygous | {Malaria, severe, resistance to}, 611162 (3) |
| *CR1* | V1,V2,V3,V4 | chr1:207569911 | missense_variant | NM_000651.6 | g.207569911G>T | c.4416G>T (p.Gln1472His) | heterozygous | {Malaria, severe, resistance to}, 611162 (3) |
| *SYT14* | V1,V2,V4 | chr1:210021220 | synonymous_variant | NM_001146262.4 | g.210021220T>C | c.408T>C (p.Tyr136Tyr) | heterozygous | ?Spinocerebellar ataxia, autosomal recessive 11, 614229 (3) |
| *OBSCN* | V1,V2,V4 | chr1:228371741 | frameshift_variant | NM_001386125.1 | g.228371741del | c.408T>C (p.Tyr136Tyr) | heterozygous | {Rhabdomyolysis, susceptibility to, 1}, 620235 (3) |
| *COG2* | V1,V2,V4 | chr1:230669469 | synonymous_variant | NM_007357.3 | g.230669469G>A | c.708G>A (p.Thr236Thr) | heterozygous | ?Congenital disorder of glycosylation, type IIq, 617395 (3) |
| *MTR* | V1,V2,V3,V4 | chr1:236803548 | missense_variant | NM_000254.3 | g.236803548G>A | c.155G>A (p.Arg52Gln) | heterozygous | {Neural tube defects, folate-sensitive, susceptibility to}, 601634 (3) |
| *FMN2* | V2 | chr1:240207872 | synonymous_variant | NM_020066.5 | g.240207872T>C | c.3060T>C (p.Pro1020Pro) | heterozygous | Intellectual developmental disorder, autosomal recessive 47, 616193 (3) |
| *FMN2* | V2 | chr1:240207884 | synonymous_variant | NM_020066.5 | g.240207884T>A | c.3072T>A (p.Pro1024Pro) | heterozygous | Intellectual developmental disorder, autosomal recessive 47, 616193 (3) |
| *FMN2* | V2 | chr1:240207887 | synonymous_variant | NM_020066.5 | g.240207887A>T | c.3075A>T (p.Leu1025Leu) | heterozygous | Intellectual developmental disorder, autosomal recessive 47, 616193 (3) |
| *FMN2* | V2 | chr1:240207905 | synonymous_variant | NM_020066.5 | g.240207905C>T | c.3093C>T (p.Pro1031Pro) | heterozygous | Intellectual developmental disorder, autosomal recessive 47, 616193 (3) |
| *FMN2* | V2 | chr1:240207911 | synonymous_variant | NM_020066.5 | g.240207911G>A | c.3099G>A (p.Pro1033Pro) | heterozygous | Intellectual developmental disorder, autosomal recessive 47, 616193 (3) |
| *FMN2* | V2 | chr1:240207917 | synonymous_variant | NM_020066.5 | g.240207917A>T | c.3105A>T (p.Pro1035Pro) | heterozygous | Intellectual developmental disorder, autosomal recessive 47, 616193 (3) |
| *FMN2* | V2,V4 | chr1:240207920 | synonymous_variant | NM_020066.5 | g.240207920T>A | c.3108T>A (p.Leu1036Leu) | heterozygous | Intellectual developmental disorder, autosomal recessive 47, 616193 (3) |
| *FMN2* | V2,V4 | chr1:240207986 | synonymous_variant | NM_020066.5 | g.240207986T>A | c.3174T>A (p.Leu1058Leu) | heterozygous | Intellectual developmental disorder, autosomal recessive 47, 616193 (3) |
| *HADHA* | V1,V2,V4 | chr2:26230216 | missense_variant | NM_000182.5 | g.26230216C>G | c.652G>C (p.Val218Leu) | heterozygous | Mitochondrial trifunctional protein deficiency 1, 609015 (3) |
| *GCKR* | V1,V2,V4 | chr2:27523329 | missense_variant | NM_001486.4 | g.27523329C>T | c.1768C>T (p.His590Tyr) | heterozygous | [Fasting plasma glucose level QTL 5], 613463 (3) |
| *ZFP36L2* | V1,V2,V3,V4 | chr2:43225375 | synonymous_variant | NM_006887.5 | g.43225375C>G | c.429G>C (p.Gly143Gly) | heterozygous | Oocyte/zygote/embryo maturation arrest 13, 620154 (3) |
| *MSH6* | V1,V2,V3,V4 | chr2:47799169 | missense_variant | NM_000179.3 | g.47799169C>G | c.1186C>G (p.Leu396Val) | heterozygous | {Endometrial cancer, familial}, 608089 (3) |
| *DYSF* | V1,V2,V3,V4 | chr2:71564125 | missense_variant | NM_001130987.2 | g.71564125G>A | c.2477G>A (p.Arg826Gln) | heterozygous | Myopathy, distal, with anterior tibial onset, 606768 (3) |
| *ALMS1* | V1,V2,V4 | chr2:73572505 | missense_variant | NM_001378454.1 | g.73572505C>G | c.10628C>G (p.Thr3543Ser) | heterozygous | Alstrom syndrome, 203800 (3) |
| *NEB* | V1,V2,V3,V4 | chr2:151602679 | missense_variant | NM_001164508.2 | g.151602679C>T | c.13276G>A (p.Asp4426Asn) | homozygous | Nemaline myopathy 2, autosomal recessive, 256030 (3) |
| *SCN9A* | V1,V2,V4 | chr2:166304242 | missense_variant | NM_001365536.1 | g.166304242G>C | c.684C>G (p.Ile228Met) | heterozygous | Small fiber neuropathy, 133020 (3) |
| *ATP5MC3* | V1,V2,V4 | chr2:175179178 | missense_variant | NM_001689.5 | g.175179178T>C | c.193A>G (p.Ile65Val) | heterozygous | Dystonia, early-onset, and/or spastic paraplegia, 619681 (3) |
| *TTN* | V1,V2,V4 | chr2:178534950 | missense_variant | NM_001267550.2 | g.178534950C>T | c.101665G>A (p.Val33889Ile) | heterozygous | Tibial muscular dystrophy, tardive, 600334 (3) |
| *TTN* | V1,V2,V3,V4 | chr2:178662395 | missense_variant | NM_001267550.2 | g.178662395T>C | c.36982A>G (p.Thr12328Ala) | heterozygous | Tibial muscular dystrophy, tardive, 600334 (3) |
| *TTN* | V1,V2,V4 | chr2:178722825 | synonymous_variant | NM_001267550.2 | g.178722825T>C | c.22074A>G (p.Lys7358Lys) | heterozygous | Tibial muscular dystrophy, tardive, 600334 (3) |
| *TTN* | V1,V2,V4 | chr2:178741264 | missense_variant | NM_001267550.2 | g.178741264G>A | c.11969C>T (p.Pro3990Leu) | heterozygous | Tibial muscular dystrophy, tardive, 600334 (3) |
| *DNAH7* | V1,V2,V4 | chr2:195884691 | missense_variant | NM_018897.3 | g.195884691C>T | c.5657G>A (p.Arg1886Gln) | heterozygous | Ciliary dyskinesia, primary, 50, 620356 (3) |
| *DNAH7* | V1,V2,V3,V4 | chr2:195923649-195923650 | frameshift_variant | NM_018897.3 | g.195923649_195923650insT | c.3770_3771insA (p.Asn1257LysTer) | heterozygous | Ciliary dyskinesia, primary, 50, 620356 (3) |
| *PGAP1* | V1,V2,V4 | chr2:196844038 | missense_variant | NM_024989.4 | g.196844038T>C | c.2375A>G (p.Asn792Ser) | heterozygous | Neurodevelopmental disorder with dysmorphic features, spasticity, and brain abnormalities, 615802 (3) |
| *SF3B1* | V1,V2,V4 | chr2:197398499 | synonymous_variant | NM_012433.4 | g.197398499T>C | c.3096A>G (p.Gln1032Gln) | heterozygous | Myelodysplastic syndrome, somatic, 614286 (3) |
| *SF3B1* | V1,V2,V4 | chr2:197403027 | synonymous_variant | NM_012433.4 | g.197403027C>T | c.1728G>A (p.Val576Val) | heterozygous | Myelodysplastic syndrome, somatic, 614286 (3) |
| *SF3B1* | V1,V2,V3,V4 | chr2:197408043 | synonymous_variant | NM_012433.4 | g.197408043T>G | c.1194A>C (p.Pro398Pro) | heterozygous | Myelodysplastic syndrome, somatic, 614286 (3) |
| *SF3B1* | V1,V2,V4 | chr2:197409843 | synonymous_variant | NM_012433.4 | g.197409843C>T | c.831G>A (p.Ala277Ala) | heterozygous | Myelodysplastic syndrome, somatic, 614286 (3) |
| *SATB2* | V1,V2,V3,V4 | chr2:199433474 | synonymous_variant | NM_001172509.2 | g.199433474G>A | c.210C>T (p.Asp70Asp) | heterozygous | Glass syndrome, 612313 (3) |
| *ALS2* | V1,V2,V3,V4 | chr2:201753242 | splice_region_variant,synonymous_variant | NM_020919.4 | g.201753242C>T | c.1641G>A (p.Arg547Arg) | heterozygous | Spastic paralysis, infantile onset ascending, 607225 (3) |
| *STK36* | V1,V2,V4 | chr2:218696531 | missense_variant | NM_015690.5 | g.218696531G>A | c.2516G>A (p.Arg839Gln) | heterozygous | ?Ciliary dyskinesia, primary, 46, 619436 (3) |
| *SPEG* | V1,V2,V4 | chr2:219467189 | missense_variant | NM_005876.5 | g.219467189G>A | c.2897G>A (p.Arg966Gln) | heterozygous | Centronuclear myopathy 5, 615959 (3) |
| *COL4A4* | V1,V2,V4 | chr2:227052367 | stop_gained | NM_000092.5 | g.227052367G>C | c.2906C>G (p.Ser969*) | heterozygous | Hematuria, familial benign, 1, 141200 (3) |
| *CHRNG* | V1,V2,V3,V4 | chr2:232545678 | missense_variant | NM_005199.5 | g.232545678C>T | c.1516C>T (p.Pro506Ser) | heterozygous | Multiple pterygium syndrome, lethal type, 253290 (3) |
| *NDUFA10* | V1,V2,V4 | chr2:240011654 | missense_variant | NM_004544.4 | g.240011654C>T | c.712G>A (p.Glu238Lys) | heterozygous | Mitochondrial complex I deficiency, nuclear type 22, 618243 (3) |
| *BTD* | V1,V2,V4 | chr3:15644367 | missense_variant | NM_001370658.1 | g.15644367G>A | c.451G>A (p.Ala151Thr) | heterozygous | Biotinidase deficiency, 253260 (3) |
| *TRAK1* | V1,V2,V4 | chr3:42200929 | missense_variant | NM_001042646.3 | g.42200929G>C | c.1302G>C (p.Met434Ile) | heterozygous | Developmental and epileptic encephalopathy 68, 618201 (3) |
| *CCR5* | V1,V2,V4 | chr3:46373293 | missense_variant | NM_001394783.1 | g.46373293G>T | c.391G>T (p.Val131Phe) | heterozygous | {West nile virus, susceptibility to}, 610379 (3) |
| *COL7A1* | V1,V2,V4 | chr3:48586607 | missense_variant | NM_000094.4 | g.48586607C>T | c.3359G>A (p.Arg1120Lys) | heterozygous | Transient bullous of the newborn, 131705 (3) |
| *COL7A1* | V1,V2,V4 | chr3:48592819 | missense_variant | NM_000094.4 | g.48592819G>A | c.802C>T (p.Pro268Ser) | heterozygous | Transient bullous of the newborn, 131705 (3) |
| *LAMB2* | V1,V2,V4 | chr3:49123880 | synonymous_variant | NM_002292.4 | g.49123880C>T | c.3645G>A (p.Ala1215Ala) | heterozygous | Pierson syndrome, 609049 (3) |
| *MST1R* | V1,V2,V4 | chr3:49895841 | missense_variant | NM_002447.4 | g.49895841C>T | c.2836G>A (p.Val946Met) | heterozygous | {Nasopharyngeal carcinoma, susceptibility to, 3}, 617075 (3) |
| *CACNA2D2* | V1,V2,V3,V4 | chr3:50503236 | missense_variant | NM_006030.4 | g.50503236C>T | c.188G>A (p.Ser63Asn) | heterozygous | Cerebellar atrophy with seizures and variable developmental delay, 618501 (3) |
| *ACOX2* | V1,V2,V4 | chr3:58534005-58534008 | frameshift_variant | NM_003500.4 | g.58534005_58534008del | c.188G>A (p.Ser63Asn) | heterozygous | Bile acid synthesis defect, congenital, 6, 617308 (3) |
| *ZBTB11* | V1,V2,V3,V4 | chr3:101665538 | missense_variant | NM_014415.4 | g.101665538G>T | c.1049C>A (p.Thr350Asn) | heterozygous | Intellectual developmental disorder, autosomal recessive 69, 618383 (3) |

| | | | | | | | | |
|---|---|---|---|---|---|---|---|---|
| *MCM2* | V1,V2,V4 | chr3:127615934 | missense_variant | NM_004526.4 | g.127615934G>A | c.1501G>A (p.Gly501Arg) | heterozygous | ?Deafness, autosomal dominant 70, 616968 (3) |
| *MSL2* | V1,V2,V4 | chr3:136151441 | synonymous_variant | NM_018133.4 | g.136151441G>A | c.1440C>T (p.Arg480Arg) | heterozygous | Karayol-Borroto-Haghshenas neurodevelopmental syndrome, 620985 (3) |
| *PIK3CA* | V1,V2,V3,V4 | chr3:179199003 | missense_variant | NM_006218.4 | g.179199003C>A | c.178C>A (p.Gln60Lys) | heterozygous | Ovarian cancer, somatic, 167000 (3) |
| *CLCN2* | V1,V2,V3,V4 | chr3:184357688 | missense_variant | NM_004366.6 | g.184357688C>T | c.704G>A (p.Arg235Gln) | heterozygous | {Epilepsy, juvenile myoclonic, susceptibility to, 8}, 607628 (3) |
| *NRROS* | V1,V2,V3,V4 | chr3:196661530 | synonymous_variant | NM_198565.3 | g.196661530C>T | c.1887C>T (p.Arg629Arg) | heterozygous | Seizures, early-onset, with neurodegeneration and brain calcification, 618875 (3) |
| *CORIN* | V1,V2,V3,V4 | chr4:47674419 | missense_variant | NM_006587.4 | g.47674419T>C | c.1331A>G (p.Asp444Gly) | heterozygous | Preeclampsia/eclampsia 5, 614595 (3) |
| *KDR* | V1,V2,V4 | chr4:55094919 | missense_variant | NM_002253.4 | g.55094919C>T | c.2854G>A (p.Val952Ile) | heterozygous | {Hemangioma, capillary infantile, susceptibility to}, 602089 (3) |
| *ENAM* | V1,V2,V4 | chr4:70644746 | missense_variant | NM_031889.3 | g.70644746G>A | c.3320G>A (p.Ser1107Asn) | heterozygous | Amelogenesis imperfecta, type IC, 204650 (3) |
| *DSPP* | V2,V3,V4 | chr4:87615730-87615765 | inframe_deletion | NM_014208.3 | g.87615730_87615765del | c.3068_3103del (p.1023_1035delinsAsn) | heterozygous | Dentinogenesis imperfecta, Shields type III, 125500 (3) |
| *DSPP* | V2,V4 | chr4:87616091 | missense_variant | NM_014208.3 | g.87616091C>A | c.3429C>A (p.Asp1143Glu) | heterozygous | Dentinogenesis imperfecta, Shields type III, 125500 (3) |
| *DSPP* | V2,V4 | chr4:87616097 | synonymous_variant | NM_014208.3 | g.87616097T>C | c.3435T>C (p.Ser1145Ser) | heterozygous | Dentinogenesis imperfecta, Shields type III, 125500 (3) |
| *DSPP* | V2,V4 | chr4:87616106 | synonymous_variant | NM_014208.3 | g.87616106T>C | c.3444T>C (p.Ser1148Ser) | heterozygous | Dentinogenesis imperfecta, Shields type III, 125500 (3) |
| *ADH1C* | V1,V2,V3,V4 | chr4:99342945 | synonymous_variant | NM_000669.5 | g.99342945G>A | c.678C>T (p.Asn226Asn) | heterozygous | {Parkinson disease, susceptibility to}, 168600 (3) |
| *SLC9A3* | V1,V2,V4 | chr5:482093 | missense_variant | NM_004174.4 | g.482093C>T | c.1421G>A (p.Arg474Gln) | heterozygous | Diarrhea 8, secretory sodium, congenital, 616868 (3) |
| *IL7R* | V1,V2,V3,V4 | chr5:35876149 | missense_variant | NM_002185.5 | g.35876149A>C | c.1043A>C (p.Asn348Thr) | heterozygous | Immunodeficiency 104, severe combined, 608971 (3) |
| *GHR* | V1,V2,V4 | chr5:42565884 | missense_variant | NM_000163.5 | g.42565884T>C | c.10T>C (p.Trp4Arg) | heterozygous | {Hypercholesterolemia, familial, modifier of}, 143890 (3) |
| *MAP1B* | V1,V2,V3,V4 | chr5:72195741 | missense_variant | NM_005909.5 | g.72195741G>A | c.2386G>A (p.Ala796Thr) | heterozygous | Periventricular nodular heterotopia 9, 618918 (3) |
| *MSH3* | V3,V4 | chr5:80654888-80654914 | inframe_deletion | NM_002439.5 | g.80654888_80654914del | c.161_187del (p.54_63delinsAla) | heterozygous | Familial adenomatous polyposis 4, 617100 (3) |
| *MSH3* | V1 | chr5:80654889-80654890 | inframe_insertion | NM_002439.5 | g.80654889_80654890insGCAGCGGCC | c.162_163insGCAGCGGCC (p.54_55insAlaAlaAla) | heterozygous | Familial adenomatous polyposis 4, 617100 (3) |
| *MSH3* | V3,V4 | chr5:80654917 | missense_variant | NM_002439.5 | g.80654917C>G | c.190C>G (p.Pro64Ala) | heterozygous | Familial adenomatous polyposis 4, 617100 (3) |
| *MSH3* | V3,V4 | chr5:80654931 | synonymous_variant | NM_002439.5 | g.80654931T>G | c.204T>G (p.Ala68Ala) | heterozygous | Familial adenomatous polyposis 4, 617100 (3) |
| *ADGRV1* | V1,V2,V4 | chr5:90745757 | missense_variant | NM_032119.4 | g.90745757T>C | c.10936T>C (p.Ser3646Pro) | heterozygous | Usher syndrome, type 2C, GPR98/PDZD7 digenic, 605472 (3) |
| *APC* | V1,V2,V3,V4 | chr5:112839920 | synonymous_variant | NM_000038.6 | g.112839920T>A | c.4326T>A (p.Pro1442Pro) | heterozygous | Hepatoblastoma, somatic, 114550 (3) |
| *APC* | V1,V2,V4 | chr5:112842515 | synonymous_variant | NM_000038.6 | g.112842515G>A | c.6921G>A (p.Ser2307Ser) | heterozygous | Hepatoblastoma, somatic, 114550 (3) |
| *GRXCR2* | V1,V2,V3,V4 | chr5:145872797 | missense_variant | NM_001080516.2 | g.145872797G>A | c.172C>T (p.Leu58Phe) | heterozygous | ?Deafness, autosomal recessive 101, 615837 (3) |
| *F12* | V1,V2,V3,V4 | chr5:177405165 | missense_variant | NM_000505.4 | g.177405165G>C | c.418C>G (p.Leu140Val) | heterozygous | Factor XII deficiency, 234000 (3) |
| *DSP* | V1,V2,V4 | chr6:7541993 | synonymous_variant | NM_004415.4 | g.7541993G>T | c.78G>T (p.Leu26Leu) | heterozygous | Keratosis palmoplantaris striata II, 612908 (3) |
| *DSP* | V1,V2,V3,V4 | chr6:7581514 | missense_variant | NM_004415.4 | g.7581514G>T | c.5324G>T (p.Arg1775Ile) | heterozygous | Keratosis palmoplantaris striata II, 612908 (3) |
| *ATXN1* | V3 | chr6:16327672-16327673 | inframe_insertion | NM_001128164.2 | g.16327672_16327673insTGCTGCTGCTGA | c.638_639insTCAGCAGCAGCA (p.Gln213delinsHisGlnGlnGlnGln) | heterozygous | Spinocerebellar ataxia 1, 164400 (3) |
| *ALDH5A1* | V1,V2,V3,V4 | chr6:24533516 | missense_variant | NM_001080.3 | g.24533516A>C | c.1412A>C (p.Tyr471Ser) | heterozygous | Succinic semialdehyde dehydrogenase deficiency, 271980 (3) |
| *HLA-A* | V1,V2,V4 | chr6:29942761 | synonymous_variant | NM_002116.8 | g.29942761C>T | c.78C>T (p.Ser26Ser) | heterozygous | {Hypersensitivity syndrome, carbamazepine-induced, susceptibility to}, 608579 (3) |
| *HLA-A* | V1,V2,V4 | chr6:29944124 | missense_variant,splice_region_va riant | NM_002116.8 | g.29944124C>G | c.622C>G (p.Pro208Ala) | heterozygous | {Hypersensitivity syndrome, carbamazepine-induced, susceptibility to}, 608579 (3) |
| *HLA-A* | V1,V2,V4 | chr6:29944144 | synonymous_variant | NM_002116.8 | g.29944144C>T | c.642C>T (p.Thr214Thr) | heterozygous | {Hypersensitivity syndrome, carbamazepine-induced, susceptibility to}, 608579 (3) |
| *HLA-A* | V1,V2,V4 | chr6:29944151 | missense_variant | NM_002116.8 | g.29944151C>G | c.649C>G (p.Pro217Ala) | heterozygous | {Hypersensitivity syndrome, carbamazepine-induced, susceptibility to}, 608579 (3) |
| *HLA-A* | V1,V2,V4 | chr6:29944153 | synonymous_variant | NM_002116.8 | g.29944153C>T | c.651C>T (p.Pro217Pro) | heterozygous | {Hypersensitivity syndrome, carbamazepine-induced, susceptibility to}, 608579 (3) |
| *HLA-A* | V1,V2,V4 | chr6:29944154 | missense_variant | NM_002116.8 | g.29944154A>G | c.652A>G (p.Ile218Val) | heterozygous | {Hypersensitivity syndrome, carbamazepine-induced, susceptibility to}, 608579 (3) |
| *HLA-A* | V1,V2,V4 | chr6:29944168 | synonymous_variant | NM_002116.8 | g.29944168G>A | c.666G>A (p.Glu222Glu) | heterozygous | {Hypersensitivity syndrome, carbamazepine-induced, susceptibility to}, 608579 (3) |
| *DHX16* | V1,V2,V3,V4 | chr6:30665484 | missense_variant | NM_003587.5 | g.30665484C>T | c.916G>A (p.Gly306Arg) | heterozygous | Neuromuscular disease and ocular or auditory anomalies with or without seizures, 618733 (3) |
| *HLA-C* | V1,V2,V3,V4 | chr6:31270009 | synonymous_variant | NM_002117.6 | g.31270009A>T | c.972T>A (p.Leu324Leu) | homozygous | {Psoriasis susceptibility 1}, 177900 (3) |
| *HLA-C* | V1,V2,V3,V4 | chr6:31270025 | missense_variant | NM_002117.6 | g.31270025A>G | c.956T>C (p.Val319Ala) | homozygous | {Psoriasis susceptibility 1}, 177900 (3) |
| *HLA-C* | V1,V2,V3,V4 | chr6:31270276 | missense_variant | NM_002117.6 | g.31270276G>C | c.829C>G (p.Gln277Glu) | homozygous | {Psoriasis susceptibility 1}, 177900 (3) |
| *HLA-B* | V1,V2,V4 | chr6:31356280 | missense_variant | NM_005514.8 | g.31356280C>A | c.506G>T (p.Arg169Leu) | heterozygous | {Toxic epidermal necrolysis, susceptibility to}, 608579 (3) |
| *HLA-B* | V1,V2,V4 | chr6:31356418 | missense_variant | NM_005514.8 | g.31356418T>G | c.368A>C (p.Tyr123Ser) | heterozygous | {Toxic epidermal necrolysis, susceptibility to}, 608579 (3) |
| *HLA-B* | V1 | chr6:31356711-31356712 | frameshift_variant | NM_005514.8 | g.31356711_31356712insGG | c.319_320insCC (p.Gly107AlaTer) | heterozygous | {Toxic epidermal necrolysis, susceptibility to}, 608579 (3) |
| *HLA-B* | V3 | chr6:31356825 | missense_variant | NM_005514.8 | g.31356825T>A | c.206A>T (p.Glu69Val) | heterozygous | {Toxic epidermal necrolysis, susceptibility to}, 608579 (3) |
| *C4B* | V1,V2,V3,V4 | chr6:32027362 | missense_variant | NM_001002029.4 | g.32027362A>G | c.2719A>G (p.Thr907Ala) | homozygous | C4B deficiency, 614379 (3) |
| *HLA-DRB1* | V2,V4 | chr6:32580255 | missense_variant | NM_002124.4 | g.32580255T>G | c.779A>C (p.Gln260Pro) | heterozygous | {Sarcoidosis, susceptibility to, 1}, 181000 (3) |
| *HLA-DRB1* | V4 | chr6:32581804 | synonymous_variant | NM_002124.4 | g.32581804G>A | c.405C>T (p.Thr135Thr) | heterozygous | {Sarcoidosis, susceptibility to, 1}, 181000 (3) |
| *HLA-DRB1* | V2,V4 | chr6:32584176 | synonymous_variant | NM_002124.4 | g.32584176C>A | c.303G>T (p.Arg101Arg) | homozygous | {Sarcoidosis, susceptibility to, 1}, 181000 (3) |
| *HLA-DRB1* | V2,V4 | chr6:32584183 | missense_variant | NM_002124.4 | g.32584183T>C | c.296A>G (p.Gln99Arg) | homozygous | {Sarcoidosis, susceptibility to, 1}, 181000 (3) |
| *HLA-DRB1* | V1,V2,V4 | chr6:32584301 | missense_variant | NM_002124.4 | g.32584301A>C | c.178T>G (p.Phe60Val) | heterozygous | {Sarcoidosis, susceptibility to, 1}, 181000 (3) |
| *HLA-DRB1* | V1 | chr6:32584352-32584353 | frameshift_variant | NM_002124.4 | g.32584352_32584353insAAACTTAA | c.126_127insTTAAGTTT (p.42LeuSerTer) | heterozygous | {Sarcoidosis, susceptibility to, 1}, 181000 (3) |
| *HLA-DRB1* | V4 | chr6:32584364 | missense_variant | NM_002124.4 | g.32584364G>C | c.115C>G (p.Gln39Glu) | homozygous | {Sarcoidosis, susceptibility to, 1}, 181000 (3) |
| *HLA-DRB1* | V1 | chr6:32589688 | synonymous_variant | NM_002124.4 | g.32589688G>A | c.55C>T (p.Leu19Leu) | heterozygous | {Sarcoidosis, susceptibility to, 1}, 181000 (3) |
| *HLA-DQA1* | V1,V4 | chr6:32641487 | missense_variant | NM_002122.5 | g.32641487G>C | c.260G>C (p.Arg87Thr) | heterozygous | {Celiac disease, susceptibility to}, 212750 (3) |
| *HLA-DQA1* | V1,V2,V4 | chr6:32641494 | missense_variant | NM_002122.5 | g.32641494G>C | c.267G>C (p.Met89Ile) | heterozygous | {Celiac disease, susceptibility to}, 212750 (3) |
| *HLA-DQA1* | V1,V2,V4 | chr6:32641509 | synonymous_variant | NM_002122.5 | g.32641509C>T | c.282C>T (p.His94His) | heterozygous | {Celiac disease, susceptibility to}, 212750 (3) |
| *HLA-DQA1* | V1,V2,V4 | chr6:32641522 | missense_variant | NM_002122.5 | g.32641522A>C | c.295A>C (p.Met99Leu) | heterozygous | {Celiac disease, susceptibility to}, 212750 (3) |
| *HLA-DQB1* | V1,V2,V4 | chr6:32664868 | synonymous_variant | NM_002123.5 | g.32664868G>T | c.309C>A (p.Thr103Thr) | heterozygous | {Multiple sclerosis, susceptibility to, 1}, 126200 (3) |
| *HLA-DQB1* | V1,V2,V4 | chr6:32664869 | missense_variant | NM_002123.5 | g.32664869G>T | c.308C>A (p.Thr103Asn) | heterozygous | {Multiple sclerosis, susceptibility to, 1}, 126200 (3) |
| *HLA-DQB1* | V1,V2,V4 | chr6:32664917 | missense_variant | NM_002123.5 | g.32664917C>A | c.260G>T (p.Arg87Leu) | heterozygous | {Multiple sclerosis, susceptibility to, 1}, 126200 (3) |
| *HLA-DQB1* | V1,V2,V4 | chr6:32664926 | missense_variant | NM_002123.5 | g.32664926G>A | c.251C>T (p.Pro84Leu) | heterozygous | {Multiple sclerosis, susceptibility to, 1}, 126200 (3) |
| *HLA-DQB1* | V1,V2,V4 | chr6:32664971 | missense_variant | NM_002123.5 | g.32664971T>A | c.206A>T (p.Tyr69Phe) | heterozygous | {Multiple sclerosis, susceptibility to, 1}, 126200 (3) |
| *HLA-DQB1* | V1,V2,V4 | chr6:32664976 | synonymous_variant | NM_002123.5 | g.32664976C>T | c.201G>A (p.Glu67Glu) | heterozygous | {Multiple sclerosis, susceptibility to, 1}, 126200 (3) |
| *HLA-DQB1* | V1,V2,V4 | chr6:32664992 | missense_variant | NM_002123.5 | g.32664992T>C | c.185A>G (p.Tyr62Cys) | heterozygous | {Multiple sclerosis, susceptibility to, 1}, 126200 (3) |
| *HLA-DQB1* | V1,V2,V4 | chr6:32664993 | missense_variant | NM_002123.5 | g.32664993A>T | c.184T>A (p.Tyr62Asn) | heterozygous | {Multiple sclerosis, susceptibility to, 1}, 126200 (3) |
| *HLA-DQB1* | V1,V2,V4 | chr6:32664998 | missense_variant | NM_002123.5 | g.32664998G>C | c.179C>G (p.Thr60Ser) | heterozygous | {Multiple sclerosis, susceptibility to, 1}, 126200 (3) |
| *HLA-DQB1* | V1,V2,V4 | chr6:32665018 | synonymous_variant | NM_002123.5 | g.32665018C>T | c.159G>A (p.Thr53Thr) | heterozygous | {Multiple sclerosis, susceptibility to, 1}, 126200 (3) |
| *SCUBE3* | V1,V2,V3,V4 | chr6:35214472 | synonymous_variant | NM_152753.4 | g.35214472C>T | c.54C>T (p.Ala18Ala) | heterozygous | Short stature, facial dysmorphism, and skeletal anomalies with or without cardiac anomalies 2, 619184 (3) |
| *EYS* | V1,V2,V3,V4 | chr6:63726672 | missense_variant | NM_001142800.2 | g.63726672T>C | c.8080A>G (p.Ile2694Val) | heterozygous | Retinitis pigmentosa 25, 602772 (3) |
| *LMBRD1* | V1,V2,V3,V4 | chr6:69796818-69796819 | frameshift_variant | NM_018368.4 | g.69796818_69796819insT | c.63_64insA (p.21Ter) | heterozygous | Methylmalonic aciduria and homocystinuria, cblF type, 277380 (3) |
| *FILIP1* | V1,V2,V3,V4 | chr6:75315194 | missense_variant | NM_015687.5 | g.75315194T>G | c.638A>C (p.Lys213Thr) | heterozygous | Neuromuscular disorder, congenital, with dysmorphic facies, 620775 (3) |
| *SYNE1* | V1,V2,V3,V4 | chr6:152436089 | missense_variant | NM_182961.4 | g.152436089G>A | c.4162C>T (p.Arg1388Trp) | heterozygous | Spinocerebellar ataxia, autosomal recessive 8, 610743 (3) |
| *LPA* | V3 | chr6:160633872 | synonymous_variant | NM_005577.4 | g.160633872G>A | c.1116C>T (p.His372His) | heterozygous | {Coronary artery disease, susceptibility to}, 618807 (3) |
| *LPA* | V2,V3 | chr6:160633899 | synonymous_variant | NM_005577.4 | g.160633899C>T | c.1089G>A (p.Gln363Gln) | heterozygous | {Coronary artery disease, susceptibility to}, 618807 (3) |
| *PDE10A* | V1,V2,V3,V4 | chr6:165336194 | synonymous_variant | NM_001385079.1 | g.165336194G>A | c.2994C>T (p.Ala998Ala) | heterozygous | Striatal degeneration, autosomal dominant, 616922 (3) |
| *TBP* | V3,V4 | chr6:170561959-170561961, chr6:170561959-170561967 | inframe_deletion | NM_003194.5 | g.170561959_170561961del, g.170561959_170561967del | c.223_225del (p.75del), c.223_231del (p.75_77del) | heterozygous | {Parkinson disease, susceptibility to}, 168600 (3) |
| *DNAH11* | V1,V2,V3,V4 | chr7:21765477 | missense_variant | NM_001277115.2 | g.21765477G>A | c.8990G>A (p.Arg2997Gln) | heterozygous | Ciliary dyskinesia, primary, 7, with or without situs inversus, 611884 (3) |
| *DNAH11* | V1,V2,V3,V4 | chr7:21842591 | missense_variant | NM_001277115.2 | g.21842591G>A | c.10739G>A (p.Arg3580His) | heterozygous | Ciliary dyskinesia, primary, 7, with or without situs inversus, 611884 (3) |
| *OGDH* | V1,V2,V3,V4 | chr7:44624382 | synonymous_variant | NM_002541.4 | g.44624382A>C | c.39A>C (p.Pro13Pro) | heterozygous | Oxoglutarate dehydrogenase deficiency, 203740 (3) |
| *GNAI1* | V1,V2,V3,V4 | chr7:80211068 | synonymous_variant | NM_002069.6 | g.80211068C>T | c.690C>T (p.Tyr230Tyr) | heterozygous | Neurodevelopmental disorder with hypotonia, impaired speech, and behavioral abnormalities, 619854 (3) |
| *PEX1* | V1,V2,V3,V4 | chr7:92519022 | synonymous_variant | NM_000466.3 | g.92519022G>C | c.330C>G (p.Pro110Pro) | heterozygous | Peroxisome biogenesis disorder 1B (NALD/IRD), 601539 (3) |
| *TRRAP* | V1,V2,V3,V4 | chr7:98910604 | synonymous_variant | NM_001375524.1 | g.98910604T>C | c.1809T>C (p.Tyr603Tyr) | heterozygous | Developmental delay with or without dysmorphic facies and autism, 618454 (3) |
| *ARPC1B* | V1,V2,V3,V4 | chr7:99390998 | synonymous_variant | NM_005720.4 | g.99390998C>T | c.606C>T (p.Cys202Cys) | heterozygous | Immunodeficiency 71 with inflammatory disease and congenital thrombocytopenia, 617718 (3) |
| *CYP3A5* | V1,V2,V3,V4 | chr7:99648330 | missense_variant | NM_000777.5 | g.99648330C>G | c.1484G>C (p.Arg495Thr) | heterozygous | {Hypertension, salt-sensitive essential, susceptibility to}, 145500 (3) |
| *CUX1* | V1,V2,V4 | chr7:102248541 | synonymous_variant | NM_181552.4 | g.102248541C>T | c.4017C>T (p.Asp1339Asp) | heterozygous | Neurodevelopmental disorder with developmental delay and with or without motor or speech delay, 618330 (3) |
| *SMO* | V1,V2,V3,V4 | chr7:129212026 | missense_variant,splice_region_va riant | NM_005631.5 | g.129212026C>T | c.1939C>T (p.Pro647Ser) | heterozygous | Pallister-Hall-like syndrome, 241800 (3) |
| *NUP205* | V1,V2,V3,V4 | chr7:135577933 | synonymous_variant | NM_015135.3 | g.135577933C>T | c.786C>T (p.Gly262Gly) | heterozygous | ?Nephrotic syndrome, type 13, 616893 (3) |
| *IFT56* | V1,V2,V3,V4 | chr7:139166872 | missense_variant | NM_024926.4 | g.139166872G>A | c.928G>A (p.Asp310Asn) | heterozygous | Biliary, renal, neurologic, and skeletal syndrome, 619534 (3) |
| *FGF17* | V1,V2,V4 | chr8:22046236 | synonymous_variant | NM_003867.4 | g.22046236C>T | c.195C>T (p.His65His) | heterozygous | Hypogonadotropic hypogonadism 20 with or without anosmia, 615270 (3) |
| *ANK1* | V1,V2,V3,V4 | chr8:41692865 | missense_variant | NM_000037.4 | g.41692865G>A | c.3641C>T (p.Ser1214Leu) | heterozygous | Spherocytosis, type 1, 182900 (3) |
| *CHD7* | V1,V2,V4 | chr8:60781401 | synonymous_variant | NM_017780.4 | g.60781401G>A | c.2067G>A (p.Thr689Thr) | heterozygous | Hypogonadotropic hypogonadism 5 with or without anosmia, 612370 (3) |
| *PKHD1L1* | V1,V2,V3,V4 | chr8:109523346 | synonymous_variant | NM_177531.6 | g.109523346C>T | c.12444C>T (p.Ser4148Ser) | heterozygous | Deafness, autosomal recessive 124, 620794 (3) |
| *TG* | V1,V2,V4 | chr8:132871398 | missense_variant | NM_003235.5 | g.132871398A>G | c.325A>G (p.Ile109Val) | heterozygous | {Autoimmune thyroid disease, susceptibility to, 3}, 608175 (3) |
| *GPAA1* | V1,V2,V4 | chr8:144086122 | missense_variant | NM_003801.4 | g.144086122G>T | c.1863G>T (p.Lys621Asn) | heterozygous | Glycosylphosphatidylinositol biosynthesis defect 15, 617810 (3) |
| *RIC1* | V1,V2,V4 | chr9:5747475 | synonymous_variant | NM_020829.4 | g.5747475A>G | c.1422A>G (p.Leu474Leu) | heterozygous | CATIFA syndrome, 618761 (3) |
| *NANS* | V1,V2,V3,V4 | chr9:98076920 | missense_variant,splice_region_va riant | NM_018946.4 | g.98076920G>A | c.351G>A (p.Met117Ile) | heterozygous | Spondyloepimetaphyseal dysplasia, Genevieve type, 610442 (3) |
| *GABBR2* | V1,V2,V4 | chr9:98708441 | synonymous_variant | NM_005458.8 | g.98708441G>A | c.297C>T (p.Leu99Leu) | heterozygous | {Nicotine dependence, susceptibility to}, 188890 (3) |
| *CRB2* | V1,V2,V4 | chr9:123373659 | missense_variant | NM_173689.7 | g.123373659C>T | c.3128C>T (p.Ser1043Phe) | heterozygous | Ventriculomegaly with cystic kidney disease, 219730 (3) |
| *CEL* | V2,V4 | chr9:133071434 | synonymous_variant | NM_001807.6 | g.133071434G>C | c.1932G>C (p.Gly644Gly) | heterozygous | Maturity-onset diabetes of the young, type VIII, 609812 (3) |

| | | | | | | | | |
|---|---|---|---|---|---|---|---|---|
| *CEL* | V1,V2,V4 | chr9:133071468 | missense_variant | NM_001807.6 | g.133071468G>C | c.1966G>C (p.Ala656Pro) | heterozygous | Maturity-onset diabetes of the young, type VIII, 609812 (3) |
| *CEL* | V1 | chr9:133071566 | synonymous_variant | NM_001807.6 | g.133071566C>G | c.2064C>G (p.Gly688Gly) | heterozygous | Maturity-onset diabetes of the young, type VIII, 609812 (3) |
| *CEL* | V1 | chr9:133071567 | missense_variant | NM_001807.6 | g.133071567G>C | c.2065G>C (p.Ala689Pro) | heterozygous | Maturity-onset diabetes of the young, type VIII, 609812 (3) |
| *DBH* | V1,V2,V3,V4 | chr9:133643415 | splice_region_variant,synonymous_variant | NM_000787.4 | g.133643415C>T | c.747C>T (p.Tyr249Tyr) | heterozygous | Orthostatic hypotension 1, due to DBH deficiency, 223360 (3) |
| *LHX3* | V1,V2,V4 | chr9:136200725 | synonymous_variant | NM_178138.6 | g.136200725C>T | c.108G>A (p.Gln36Gln) | heterozygous | Pituitary hormone deficiency, combined, 3, 221750 (3) |
| *NOTCH1* | V1,V2,V4 | chr9:136503225 | synonymous_variant | NM_017617.5 | g.136503225C>A | c.5124G>T (p.Ser1708Ser) | heterozygous | Aortic valve disease 1, 109730 (3) |
| *ABCA2* | V1,V2,V3,V4 | chr9:137011443 | synonymous_variant | NM_001606.5 | g.137011443C>A | c.5763G>T (p.Val1921Val) | heterozygous | Intellectual developmental disorder with poor growth and with or without seizures or ataxia, 618808 (3) |
| *MAN1B1* | V1,V2,V4 | chr9:137108423 | synonymous_variant | NM_016219.5 | g.137108423C>T | c.1932C>T (p.Val644Val) | heterozygous | Rafiq syndrome, 614202 (3) |
| *WDR37* | V1,V2,V3,V4 | chr10:1129240 | missense_variant | NM_014023.4 | g.1129240G>T | c.1381G>T (p.Ala461Ser) | heterozygous | Neurooculocardiogenitourinary syndrome, 618652 (3) |
| *PITRM1* | V1,V2,V4 | chr10:3140811 | missense_variant,splice_region_variant | NM_014889.4 | g.3140811G>A | c.2647C>T (p.Leu883Phe) | heterozygous | Spinocerebellar ataxia, autosomal recessive 30, 619405 (3) |
| *CACNB2* | V1,V2,V4 | chr10:18539557 | missense_variant | NM_201596.3 | g.18539557C>G | c.1816C>G (p.Arg606Gly) | heterozygous | Brugada syndrome 4, 611876 (3) |
| *ODAD2* | V1,V2,V4 | chr10:27985204 | synonymous_variant | NM_018076.5 | g.27985204T>C | c.390A>G (p.Arg130Arg) | heterozygous | Ciliary dyskinesia, primary, 23, 615451 (3) |
| *ANK3* | V1,V2,V3,V4 | chr10:60071176 | synonymous_variant | NM_020987.5 | g.60071176G>A | c.9705C>T (p.Asp3235Asp) | heterozygous | Intellectual developmental disorder, autosomal recessive 37, 615493 (3) |
| *PLAU* | V1,V2,V3,V4 | chr10:73914791 | missense_variant | NM_002658.6 | g.73914791G>A | c.845G>A (p.Arg282His) | heterozygous | {Alzheimer disease, late-onset, susceptibility to}, 104300 (3) |
| *POLR3A* | V1,V2,V4 | chr10:78022289 | synonymous_variant | NM_007055.4 | g.78022289C>T | c.741G>A (p.Pro247Pro) | heterozygous | Wiedemann-Rautenstrauch syndrome, 264090 (3) |
| *LDB3* | V1,V2,V3,V4 | chr10:86687270 | splice_region_variant,synonymous_variant | NM_001368067.1 | g.86687270T>C | c.546T>C (p.Ser182Ser) | heterozygous | Myopathy, myofibrillar, 4, 609452 (3) |
| *DEAF1* | V1,V2,V4 | chr11:686935 | missense_variant | NM_021008.4 | g.686935T>C | c.727A>G (p.Met243Val) | heterozygous | Vulto-van Silfout-de Vries syndrome, 615828 (3) |
| *MUC5B* | V1,V2,V4 | chr11:1250617 | synonymous_variant | NM_002458.3 | g.1250617C>G | c.13737C>G (p.Ala4579Ala) | heterozygous | {Pulmonary fibrosis, idiopathic, susceptibility to}, 178500 (3) |
| *FAR1* | V1,V2,V3,V4 | chr11:13700413 | missense_variant | NM_032228.6 | g.13700413G>A | c.286G>A (p.Glu96Lys) | heterozygous | Peroxisomal fatty acyl-CoA reductase 1 disorder, 616154 (3) |
| *PIK3C2A* | V1,V2,V4 | chr11:17091430 | synonymous_variant | NM_002645.4 | g.17091430T>C | c.4782A>G (p.Pro1594Pro) | heterozygous | Oculoskeletodental syndrome, 618440 (3) |
| *TENM4* | V1,V2,V3,V4 | chr11:78658358 | synonymous_variant | NM_001098816.3 | g.78658358G>A | c.8010C>T (p.Tyr2670Tyr) | heterozygous | Essential tremor, hereditary, 5, 616736 (3) |
| *TENM4* | V1,V2,V3,V4 | chr11:78812231 | synonymous_variant | NM_001098816.3 | g.78812231G>A | c.1869C>T (p.Cys623Cys) | heterozygous | Essential tremor, hereditary, 5, 616736 (3) |
| *CEP295* | V1,V2,V4 | chr11:93696992 | missense_variant | NM_033395.2 | g.93696992C>A | c.2080C>A (p.Arg694Ser) | heterozygous | Seckel syndrome 11, 620767 (3) |
| *CACNA2D4* | V1,V2,V3,V4 | chr12:1844466 | stop_gained | NM_172364.5 | g.1844466G>T | c.2406C>A (p.Tyr802*) | heterozygous | Retinal cone dystrophy 4, 610478 (3) |
| *VWF* | V1,V2,V3,V4 | chr12:5993928 | missense_variant | NM_000552.5 | g.5993928C>A | c.6532G>T (p.Ala2178Ser) | heterozygous | von Willebrand disease, types 2A, 2B, 2M, and 2N, 613554 (3) |
| *VWF* | V1,V2,V3,V4 | chr12:6031493 | missense_variant | NM_000552.5 | g.6031493C>T | c.2771G>A (p.Arg924Gln) | heterozygous | von Willebrand disease, types 2A, 2B, 2M, and 2N, 613554 (3) |
| *ATN1* | V1,V2,V4 | chr12:6936729-6936743 | inframe_insertion | NM_001940.4 | g.6936729_6936743delinsCAGCAGCAGCAGCAGCAGCAGCAGCAGCAG | c.1462_1476delinsCAGCAGCAGCAGCAGCAGCAGCAGCAGCAG (p.488_492delinsGlnGlnGlnGlnGlnGlnGlnGlnGlnGln | heterozygous | Dentatorubral-pallidoluysian atrophy, 125370 (3) |
| *A2ML1* | V1,V2,V3,V4 | chr12:8868565 | missense_variant | NM_144670.6 | g.8868565A>G | c.4090A>G (p.Met1364Val) | heterozygous | {Otitis media, susceptibility to}, 166760 (3) |
| *PHC1* | V1,V2,V3,V4 | chr12:8932856 | missense_variant | NM_004426.3 | g.8932856C>A | c.1399C>A (p.Gln467Lys) | heterozygous | ?Microcephaly 11, primary, autosomal recessive, 615414 (3) |
| *PTPRO* | V1,V2,V4 | chr12:15508630 | missense_variant | NM_030667.3 | g.15508630G>A | c.1327G>A (p.Val443Ile) | heterozygous | Nephrotic syndrome, type 6, 614196 (3) |
| *COL2A1* | V1,V2,V3,V4 | chr12:47974302 | synonymous_variant | NM_001844.5 | g.47974302G>A | c.4104C>T (p.Pro1368Pro) | heterozygous | Stickler syndrome, type I, nonsyndromic ocular, 609508 (3) |
| *WNT10B* | V1,V2,V4 | chr12:48966364 | missense_variant | NM_003394.4 | g.48966364G>A | c.901C>T (p.Pro301Ser) | heterozygous | Tooth agenesis, selective, 8, 617073 (3) |
| *KMT2D* | V1,V2,V4 | chr12:49034855 | missense_variant | NM_003482.4 | g.49034855C>T | c.10312G>A (p.Val3438Met) | heterozygous | Kabuki syndrome 1, 147920 (3) |
| *KMT2D* | V1,V2,V3,V4 | chr12:49040572 | missense_variant | NM_003482.4 | g.49040572G>C | c.7198C>G (p.Pro2400Ala) | heterozygous | Kabuki syndrome 1, 147920 (3) |
| *KRT81* | V1,V2,V4 | chr12:52287624 | missense_variant | NM_002281.4 | g.52287624G>A | c.998C>T (p.Thr333Met) | heterozygous | Monilethrix 2, 621169 (3) |
| *OTOGL* | V1,V2,V3,V4 | chr12:80358705 | synonymous_variant | NM_001378609.3 | g.80358705C>T | c.6156C>T (p.Leu2052Leu) | heterozygous | Deafness, autosomal recessive 84B, 614944 (3) |
| *PTPRQ* | V1,V2,V3,V4 | chr12:80541660 | missense_variant | NM_001145026.2 | g.80541660A>G | c.3260A>G (p.Tyr1087Cys) | heterozygous | Deafness, autosomal recessive 84A, 613391 (3) |
| *POLE* | V1,V2,V4 | chr12:132668439 | missense_variant | NM_006231.4 | g.132668439G>C | c.2090C>G (p.Pro697Arg) | heterozygous | {Colorectal cancer, susceptibility to, 12}, 615083 (3) |
| *MIPEP* | V1,V2,V4 | chr13:23837737 | missense_variant | NM_005932.4 | g.23837737C>T | c.1358G>A (p.Arg453His) | heterozygous | Combined oxidative phosphorylation deficiency 31, 617228 (3) |
| *LACC1* | V1,V2,V3,V4 | chr13:43881097 | missense_variant | NM_153218.4 | g.43881097A>G | c.112A>G (p.Lys38Glu) | heterozygous | Juvenile arthritis, 618795 (3) |
| *SLITRK6* | V1,V2,V4 | chr13:85796436 | missense_variant | NM_032229.3 | g.85796436G>A | c.73C>T (p.Leu25Phe) | heterozygous | Deafness and myopia, 221200 (3) |
| *ZIC2* | V1,V2,V3,V4 | chr13:99985026 | missense_variant | NM_007129.5 | g.99985026G>A | c.1156G>A (p.Val386Ile) | heterozygous | Holoprosencephaly 5, 609637 (3) |
| *IRS2* | V1,V2,V3,V4 | chr13:109783949-109783950 | inframe_insertion | NM_003749.3 | g.109783949_109783950insCGG | c.2104_2105insCCG (p.Val702delinsAlaVal) | heterozygous | {Diabetes mellitus, noninsulin-dependent}, 125853 (3) |
| *COL4A2* | V1,V2,V4 | chr13:110512039 | missense_variant | NM_001846.4 | g.110512039G>A | c.4987G>A (p.Gly1663Ser) | heterozygous | {Hemorrhage, intracerebral, susceptibility to}, 614519 (3) |
| *ZFHX2* | V1,V2,V3,V4 | chr14:23535220 | missense_variant | NM_033400.3 | g.23535220G>A | c.106C>T (p.Pro36Ser) | heterozygous | ?Marsili syndrome, 147430 (3) |
| *PRKD1* | V1 | chr14:29927416 | missense_variant | NM_002742.3 | g.29927416G>A | c.97C>T (p.Pro33Ser) | heterozygous | Congenital heart defects and ectodermal dysplasia, 617364 (3) |
| *PTGDR* | V1,V2,V4 | chr14:52274778 | synonymous_variant | NM_000953.3 | g.52274778G>A | c.894G>A (p.Arg298Arg) | heterozygous | {Asthma, susceptibility to, 1}, 607277 (3) |
| *TRMT5* | V1,V2,V4 | chr14:60979271 | synonymous_variant | NM_020810.3 | g.60979271T>G | c.627A>C (p.Leu209Leu) | heterozygous | Peripheral neuropathy with variable spasticity, exercise intolerance, and developmental delay, 616539 (3) |
| *SPTB* | V1,V2,V4 | chr14:64786950 | synonymous_variant | NM_001355436.2 | g.64786950G>A | c.3015C>T (p.Ala1005Ala) | heterozygous | Spherocytosis, type 2, 616649 (3) |
| *LYSET* | V1,V2,V3,V4 | chr14:93186501 | missense_variant | NM_001098621.4 | g.93186501C>A | c.322C>A (p.Leu108Met) | heterozygous | Dysostosis multiplex, Ain-Naz type, 619345 (3) |
| *GLRX5* | V1,V2,V3,V4 | chr14:95535293 | synonymous_variant | NM_016417.3 | g.95535293C>T | c.204C>T (p.Gly68Gly) | heterozygous | Spasticity, childhood-onset, with hyperglycinemia, 616859 (3) |
| *JAG2* | V1,V2,V4 | chr14:105143063 | missense_variant | NM_002226.5 | g.105143063G>A | c.3349C>T (p.Arg1117Trp) | heterozygous | Muscular dystrophy, limb-girdle, autosomal recessive 27, 619566 (3) |
| *HERC2* | V1,V2,V4 | chr15:28257130 | synonymous_variant | NM_004667.6 | g.28257130G>A | c.2448C>T (p.Ser816Ser) | heterozygous | [Skin/hair/eye pigmentation 1, blue/nonblue eyes], 227220 (3) |
| *RASGRP1* | V1,V2,V4 | chr15:38512876 | synonymous_variant | NM_005739.4 | g.38512876G>A | c.756C>T (p.Asn252Asn) | heterozygous | Immunodeficiency 64, 618534 (3) |
| *MGA* | V1,V2,V3,V4 | chr15:41767144 | missense_variant | NM_001400225.1 | g.41767144T>C | c.9209T>C (p.Val3070Ala) | heterozygous | Premature ovarian failure 26, 621065 (3) |
| *TRPM7* | V1,V2,V3,V4 | chr15:50648785 | missense_variant | NM_017672.6 | g.50648785T>C | c.223A>G (p.Ile75Val) | heterozygous | {Amyotrophic lateral sclerosis-parkinsonism/dementia complex, susceptibility to}, 105500 (3) |
| *CILP* | V1,V2,V3,V4 | chr15:65197542 | missense_variant | NM_003613.4 | g.65197542T>G | c.2744A>C (p.Tyr915Ser) | heterozygous | {Lumbar disc disease, susceptibility to}, 603932 (3) |
| *THSD4* | V1,V2,V3,V4 | chr15:71215264 | missense_variant | NM_024817.3 | g.71215264C>T | c.329C>T (p.Thr110Met) | heterozygous | Aortic aneurysm, familial thoracic 12, 619825 (3) |
| *IGF1R* | V1,V2,V3,V4 | chr15:98911384 | missense_variant | NM_000875.5 | g.98911384G>A | c.1532G>A (p.Arg511Gln) | heterozygous | Insulin-like growth factor I, resistance to, 270450 (3) |
| *ABAT* | V1,V2,V3,V4 | chr16:8735769 | synonymous_variant | NM_020686.6 | g.8735769G>A | c.30G>A (p.Leu10Leu) | heterozygous | GABA-transaminase deficiency, 613163 (3) |
| *XYLT1* | V1,V2,V4 | chr16:17198347 | missense_variant | NM_022166.4 | g.17198347G>A | c.1154C>T (p.Pro385Leu) | heterozygous | {Pseudoxanthoma elasticum, modifier of severity of}, 264800 (3) |
| *KATNIP* | V1,V2,V4 | chr16:27749809 | missense_variant | NM_015202.5 | g.27749809C>T | c.2849C>T (p.Ser950Leu) | heterozygous | Joubert syndrome 26, 616784 (3) |
| *PHKB* | V1,V2,V3,V4 | chr16:47698565 | missense_variant | NM_000293.3 | g.47698565C>T | c.3121C>T (p.Arg1041Trp) | heterozygous | Phosphorylase kinase deficiency of liver and muscle, autosomal recessive, 261750 (3) |
| *ZNF423* | V1,V2,V4 | chr16:49635783 | synonymous_variant | NM_001379286.1 | g.49635783G>A | c.3393C>T (p.Pro1131Pro) | heterozygous | Nephronophthisis 14, 614844 (3) |
| *BEAN1* | V1,V2,V4 | chr16:66469744 | synonymous_variant | NM_001178020.3 | g.66469744G>A | c.168G>A (p.Val56Val) | heterozygous | Spinocerebellar ataxia 31, 117210 (3) |
| *HYDIN* | V1,V2,V3,V4 | chr16:70855244 | synonymous_variant | NM_001270974.2 | g.70855244A>G | c.12327T>C (p.Ile4109Ile) | heterozygous | Ciliary dyskinesia, primary, 5, 608647 (3) |
| *HYDIN* | V1,V2,V4 | chr16:70988074 | synonymous_variant | NM_001270974.2 | g.70988074C>A | c.4044G>T (p.Ser1348Ser) | heterozygous | Ciliary dyskinesia, primary, 5, 608647 (3) |
| *HYDIN* | V1,V2,V3,V4 | chr16:71067348 | missense_variant | NM_001270974.2 | g.71067348G>A | c.2017C>T (p.Leu673Phe) | heterozygous | Ciliary dyskinesia, primary, 5, 608647 (3) |
| *HP* | V2,V3,V4 | chr16:72057396 | synonymous_variant | NM_005143.5 | g.72057396A>G | c.195A>G (p.Val65Val) | heterozygous | [Hypohaptoglobinemia], 614081 (3) |
| *HP* | V2,V3,V4 | chr16:72057408 | synonymous_variant | NM_005143.5 | g.72057408T>C | c.207T>C (p.Asn69Asn) | heterozygous | [Hypohaptoglobinemia], 614081 (3) |
| *HP* | V2,V3,V4 | chr16:72057409 | missense_variant | NM_005143.5 | g.72057409G>A | c.208G>A (p.Asp70Asn) | heterozygous | [Hypohaptoglobinemia], 614081 (3) |
| *HP* | V2,V3,V4 | chr16:72057412 | missense_variant | NM_005143.5 | g.72057412A>G | c.211A>G (p.Lys71Glu) | heterozygous | [Hypohaptoglobinemia], 614081 (3) |
| *DNAAF1* | V1,V2,V3,V4 | chr16:84150306 | missense_variant | NM_178452.6 | g.84150306C>A | c.316C>A (p.Pro106Thr) | heterozygous | Ciliary dyskinesia, primary, 13, 613193 (3) |
| *PIEZO1* | V1,V2,V3,V4 | chr16:88732484 | missense_variant | NM_001142864.4 | g.88732484G>A | c.2842C>T (p.Arg948Cys) | heterozygous | [ER blood group system], 620207 (3) |
| *GEMIN4* | V1,V2,V4 | chr17:746458 | missense_variant | NM_015721.3 | g.746458T>A | c.1585A>T (p.Thr529Ser) | heterozygous | Neurodevelopmental disorder with microcephaly, cataracts, and renal abnormalities, 617913 (3) |
| *DNAH2* | V1,V2,V4 | chr17:7740849 | missense_variant | NM_020877.5 | g.7740849T>C | c.1546T>C (p.Tyr516His) | heterozygous | Spermatogenic failure 45, 619094 (3) |
| *DNAH2* | V1,V2,V3,V4 | chr17:7766443 | missense_variant | NM_020877.5 | g.7766443G>A | c.3637G>A (p.Glu1213Lys) | heterozygous | Spermatogenic failure 45, 619094 (3) |
| *CHD3* | V1,V2,V3,V4 | chr17:7898529 | synonymous_variant | NM_001005273.3 | g.7898529C>A | c.2085C>A (p.Pro695Pro) | heterozygous | Snijders Blok-Campeau syndrome, 618205 (3) |
| *ARHGEF15* | V1,V2,V3,V4 | chr17:8312972 | missense_variant | NM_173728.4 | g.8312972C>T | c.652C>T (p.Arg218Trp) | heterozygous | Brain small vessel disease 5 with osteoporosis, 621331 (3) |
| *MYH8* | V1,V2,V3,V4 | chr17:10398565 | missense_variant | NM_002472.3 | g.10398565C>G | c.4057G>C (p.Glu1353Gln) | heterozygous | Trismus-pseudocamptodactyly syndrome, 158300 (3) |
| *MYH8* | V1,V2,V3,V4 | chr17:10404338 | missense_variant | NM_002472.3 | g.10404338C>T | c.2680G>A (p.Val894Ile) | heterozygous | Trismus-pseudocamptodactyly syndrome, 158300 (3) |
| *MYH3* | V1,V2,V4 | chr17:10638363 | missense_variant | NM_002470.4 | g.10638363G>A | c.3409C>T (p.Arg1137Cys) | heterozygous | Contractures, pterygia, and spondylocarpotarsal fusion syndrome 1B, 618469 (3) |
| *DNAH9* | V1,V2,V4 | chr17:11822854 | synonymous_variant | NM_001372.4 | g.11822854C>T | c.9066C>T (p.Val3022Val) | heterozygous | Ciliary dyskinesia, primary, 40, 618300 (3) |
| *KCNJ18* | V1,V2,V4 | chr17:21702795 | synonymous_variant | NM_001194958.2 | g.21702795G>A | c.9G>A (p.Ala3Ala) | heterozygous | {Thyrotoxic periodic paralysis, susceptibility to, 2}, 613239 (3) |
| *KCNJ18* | V1,V2,V4 | chr17:21702830 | missense_variant | NM_001194958.2 | g.21702830T>C | c.44T>C (p.Leu15Ser) | heterozygous | {Thyrotoxic periodic paralysis, susceptibility to, 2}, 613239 (3) |
| *KCNJ18* | V2,V4 | chr17:21702914 | missense_variant | NM_001194958.2 | g.21702914G>A | c.128G>A (p.Arg43His) | heterozygous | {Thyrotoxic periodic paralysis, susceptibility to, 2}, 613239 (3) |
| *KCNJ18* | V1,V2,V4 | chr17:21702999 | missense_variant | NM_001194958.2 | g.21702999G>A | c.213G>A (p.Met71Ile) | heterozygous | {Thyrotoxic periodic paralysis, susceptibility to, 2}, 613239 (3) |
| *KCNJ18* | V1,V2,V4 | chr17:21703029 | synonymous_variant | NM_001194958.2 | g.21703029C>G | c.243C>G (p.Arg81Arg) | heterozygous | {Thyrotoxic periodic paralysis, susceptibility to, 2}, 613239 (3) |
| *KCNJ18* | V1,V4 | chr17:21703211 | missense_variant | NM_001194958.2 | g.21703211C>A | c.425C>A (p.Thr142Asn) | heterozygous | {Thyrotoxic periodic paralysis, susceptibility to, 2}, 613239 (3) |
| *KCNJ18* | V1,V2,V4 | chr17:21703219 | missense_variant | NM_001194958.2 | g.21703219G>A | c.433G>A (p.Gly145Ser) | heterozygous | {Thyrotoxic periodic paralysis, susceptibility to, 2}, 613239 (3) |
| *KCNJ18* | V2,V4 | chr17:21703242 | synonymous_variant | NM_001194958.2 | g.21703242G>A | c.456G>A (p.Thr152Thr) | heterozygous | {Thyrotoxic periodic paralysis, susceptibility to, 2}, 613239 (3) |
| *KCNJ18* | V1,V2,V4 | chr17:21703568 | missense_variant | NM_001194958.2 | g.21703568G>A | c.782G>A (p.Arg261His) | heterozygous | {Thyrotoxic periodic paralysis, susceptibility to, 2}, 613239 (3) |
| *KCNJ18* | V1 | chr17:21703571 | missense_variant | NM_001194958.2 | g.21703571T>G | c.785T>G (p.Ile262Ser) | heterozygous | {Thyrotoxic periodic paralysis, susceptibility to, 2}, 613239 (3) |
| *KCNJ18* | V1 | chr17:21703584 | synonymous_variant | NM_001194958.2 | g.21703584G>A | c.798G>A (p.Ser266Ser) | heterozygous | {Thyrotoxic periodic paralysis, susceptibility to, 2}, 613239 (3) |
| *KCNJ18* | V1 | chr17:21703597 | synonymous_variant | NM_001194958.2 | g.21703597T>C | c.811T>C (p.Leu271Leu) | heterozygous | {Thyrotoxic periodic paralysis, susceptibility to, 2}, 613239 (3) |
| *KCNJ18* | V1 | chr17:21703602 | synonymous_variant | NM_001194958.2 | g.21703602T>C | c.816T>C (p.His272His) | heterozygous | {Thyrotoxic periodic paralysis, susceptibility to, 2}, 613239 (3) |
| *KCNJ18* | V1 | chr17:21703605 | synonymous_variant | NM_001194958.2 | g.21703605A>G | c.819A>G (p.Glu273Glu) | heterozygous | {Thyrotoxic periodic paralysis, susceptibility to, 2}, 613239 (3) |
| *KCNJ18* | V1 | chr17:21703620 | synonymous_variant | NM_001194958.2 | g.21703620C>T | c.834C>T (p.Ser278Ser) | heterozygous | {Thyrotoxic periodic paralysis, susceptibility to, 2}, 613239 (3) |
| *KCNJ18* | V2,V4 | chr17:21703848 | synonymous_variant | NM_001194958.2 | g.21703848G>A | c.1062G>A (p.Thr354Thr) | heterozygous | {Thyrotoxic periodic paralysis, susceptibility to, 2}, 613239 (3) |
| *KCNJ18* | V4 | chr17:21703860 | synonymous_variant | NM_001194958.2 | g.21703860T>C | c.1074T>C (p.Ser358Ser) | heterozygous | {Thyrotoxic periodic paralysis, susceptibility to, 2}, 613239 (3) |

| | | | | | | | | |
|---|---|---|---|---|---|---|---|---|
| *KCNJ18* | V2,V4 | chr17:21703875 | synonymous_variant | NM_001194958.2 | g.21703875A>G | c.1089A>G (p.Val363Val) | heterozygous | {Thyrotoxic periodic paralysis, susceptibility to, 2}, 613239 (3) |
| *KCNJ18* | V2,V4 | chr17:21703899 | missense_variant | NM_001194958.2 | g.21703899T>G | c.1113T>G (p.Ser371Arg) | heterozygous | {Thyrotoxic periodic paralysis, susceptibility to, 2}, 613239 (3) |
| *KCNJ18* | V2,V4 | chr17:21703917 | synonymous_variant | NM_001194958.2 | g.21703917T>C | c.1131T>C (p.Tyr377Tyr) | heterozygous | {Thyrotoxic periodic paralysis, susceptibility to, 2}, 613239 (3) |
| *KCNJ18* | V4 | chr17:21703918 | missense_variant | NM_001194958.2 | g.21703918G>A | c.1132G>A (p.Glu378Lys) | heterozygous | {Thyrotoxic periodic paralysis, susceptibility to, 2}, 613239 (3) |
| *KCNJ18* | V2,V4 | chr17:21703977 | synonymous_variant | NM_001194958.2 | g.21703977C>T | c.1191C>T (p.Asp397Asp) | heterozygous | {Thyrotoxic periodic paralysis, susceptibility to, 2}, 613239 (3) |
| *KCNJ18* | V2,V4 | chr17:21703992 | missense_variant | NM_001194958.2 | g.21703992T>A | c.1206T>A (p.Asp402Glu) | heterozygous | {Thyrotoxic periodic paralysis, susceptibility to, 2}, 613239 (3) |
| *KCNJ18* | V2,V4 | chr17:21704000 | missense_variant | NM_001194958.2 | g.21704000G>T | c.1214G>T (p.Ser405Ile) | heterozygous | {Thyrotoxic periodic paralysis, susceptibility to, 2}, 613239 (3) |
| *KCNJ18* | V2,V4 | chr17:21704040 | synonymous_variant | NM_001194958.2 | g.21704040C>T | c.1254C>T (p.Gly418Gly) | heterozygous | {Thyrotoxic periodic paralysis, susceptibility to, 2}, 613239 (3) |
| *KCNJ18* | V2,V4 | chr17:21704075 | missense_variant | NM_001194958.2 | g.21704075G>A | c.1289G>A (p.Gly430Glu) | heterozygous | {Thyrotoxic periodic paralysis, susceptibility to, 2}, 613239 (3) |
| *KCNJ18* | V2,V4 | chr17:21704087 | stop_retained_variant | NM_001194958.2 | g.21704087G>A | c.1301G>A (p.*434*) | heterozygous | {Thyrotoxic periodic paralysis, susceptibility to, 2}, 613239 (3) |
| *LIG3* | V1,V2,V4 | chr17:35002773 | missense_variant | NM_013975.4 | g.35002773C>T | c.2780C>T (p.Thr927Met) | heterozygous | Mitochondrial DNA depletion syndrome 20 (MNGIE type), 619780 (3) |
| *GPR179* | V1,V2,V4 | chr17:38330430-38330431 | frameshift_variant | NM_001004334.4 | g.38330430_38330431del | c.3138_3139del (p.1046_1047AspTer) | heterozygous | Night blindness, congenital stationary (complete), 1E, autosomal recessive, 614565 (3) |
| *JUP* | V1,V2,V4 | chr17:41763256 | synonymous_variant | NM_002230.4 | g.41763256G>A | c.1224C>T (p.Leu408Leu) | heterozygous | Naxos disease, 601214 (3) |
| *PLEKHM1* | V1,V2,V3,V4 | chr17:45475576 | synonymous_variant | NM_014798.3 | g.45475576G>A | c.447C>T (p.Tyr149Tyr) | heterozygous | Osteopetrosis, autosomal dominant 3, 618107 (3) |
| *MAPT* | V1,V2,V3,V4 | chr17:45983243 | missense_variant | NM_001377265.1 | g.45983243C>T | c.664C>T (p.His222Tyr) | heterozygous | {Parkinson disease, susceptibility to}, 168600 (3) |
| *RAD51C* | V1,V2,V4 | chr17:58696773 | missense_variant | NM_058216.3 | g.58696773G>A | c.485G>A (p.Gly162Glu) | heterozygous | {Breast-ovarian cancer, familial, susceptibility to, 3}, 613399 (3) |
| *SMG8* | V1,V2,V4 | chr17:59210306 | synonymous_variant | NM_018149.7 | g.59210306T>C | c.255T>C (p.Pro85Pro) | heterozygous | Alzahrani-Kuwahara syndrome, 619268 (3) |
| *RHBDF2* | V1,V2,V3,V4 | chr17:76477777 | synonymous_variant | NM_001005498.4 | g.76477777C>T | c.681G>A (p.Ser227Ser) | heterozygous | Tylosis with esophageal cancer, 148500 (3) |
| *RNF213* | V1,V2,V3,V4 | chr17:80383800 | missense_variant | NM_001256071.3 | g.80383800A>G | c.14194A>G (p.Lys4732Glu) | heterozygous | {Moyamoya disease 2, susceptibility to}, 607151 (3) |
| *CSNK1D* | V1,V2,V3,V4 | chr17:82249578 | missense_variant | NM_001893.6 | g.82249578C>T | c.910G>A (p.Ala304Thr) | heterozygous | Advanced sleep-phase syndrome, familial, 2, 615224 (3) |
| *LAMA3* | V1,V2,V3,V4 | chr18:23921039 | synonymous_variant | NM_198129.4 | g.23921039C>T | c.8028C>T (p.Asn2676Asn) | heterozygous | Epidermolysis bullosa, junctional 2C, laryngoonychocutaneous, 245660 (3) |
| *MOCOS* | V1,V2,V3,V4 | chr18:36200050 | missense_variant | NM_017947.4 | g.36200050C>T | c.667C>T (p.Arg223Trp) | heterozygous | Xanthinuria, type II, 603592 (3) |
| *SKOR2* | V1,V2,V3,V4 | chr18:47246697 | synonymous_variant | NM_001278063.4 | g.47246697G>A | c.2487C>T (p.Pro829Pro) | heterozygous | Valence-Farazi cerebellar ataxia syndrome, 621386 (3) |
| *RTTN* | V1,V2,V3,V4 | chr18:70197682 | missense_variant | NM_173630.4 | g.70197682T>G | c.635A>C (p.Asp212Ala) | heterozygous | Microcephaly, short stature, and polymicrogyria with seizures, 614833 (3) |
| *GPX4* | V1,V2,V4 | chr19:1105691 | missense_variant | NM_002085.5 | g.1105691G>A | c.358G>A (p.Ala120Thr) | heterozygous | Spondylometaphyseal dysplasia, Sedaghatian type, 250220 (3) |
| *PIP5K1C* | V1,V2,V4 | chr19:3633466 | missense_variant | NM_012398.3 | g.3633466G>A | c.1975C>T (p.Pro659Ser) | heterozygous | Lethal congenital contractural syndrome 3, 611369 (3) |
| *PLIN4* | V1,V2,V4 | chr19:4511377 | synonymous_variant | NM_001367868.2 | g.4511377T>G | c.2583A>C (p.Thr861Thr) | homozygous | Myopathy with rimmed ubiquitin-positive autophagic vacuolation, autosomal dominant, 601846 (3) |
| *PLIN4* | V1,V2 | chr19:4511383 | synonymous_variant | NM_001367868.2 | g.4511383T>A | c.2577A>T (p.Thr859Thr) | heterozygous | Myopathy with rimmed ubiquitin-positive autophagic vacuolation, autosomal dominant, 601846 (3) |
| *PLIN4* | V1,V2,V4 | chr19:4511387 | missense_variant | NM_001367868.2 | g.4511387G>A | c.2573C>T (p.Ala858Val) | homozygous | Myopathy with rimmed ubiquitin-positive autophagic vacuolation, autosomal dominant, 601846 (3) |
| *PLIN4* | V1,V2 | chr19:4511388 | missense_variant | NM_001367868.2 | g.4511388C>G | c.2572G>C (p.Ala858Pro) | homozygous | Myopathy with rimmed ubiquitin-positive autophagic vacuolation, autosomal dominant, 601846 (3) |
| *PLIN4* | V1,V2,V4 | chr19:4511391 | missense_variant | NM_001367868.2 | g.4511391T>C | c.2569A>G (p.Ile857Val) | homozygous | Myopathy with rimmed ubiquitin-positive autophagic vacuolation, autosomal dominant, 601846 (3) |
| *PLIN4* | V1,V2,V4 | chr19:4511393 | missense_variant | NM_001367868.2 | g.4511393T>G | c.2567A>C (p.Asn856Thr) | homozygous | Myopathy with rimmed ubiquitin-positive autophagic vacuolation, autosomal dominant, 601846 (3) |
| *PLIN4* | V1,V2 | chr19:4511395 | synonymous_variant | NM_001367868.2 | g.4511395T>C | c.2565A>G (p.Gln855Gln) | homozygous | Myopathy with rimmed ubiquitin-positive autophagic vacuolation, autosomal dominant, 601846 (3) |
| *PLIN4* | V1,V2 | chr19:4511397 | missense_variant | NM_001367868.2 | g.4511397G>T | c.2563C>A (p.Gln855Lys) | homozygous | Myopathy with rimmed ubiquitin-positive autophagic vacuolation, autosomal dominant, 601846 (3) |
| *PLIN4* | V1,V2 | chr19:4511401 | synonymous_variant | NM_001367868.2 | g.4511401C>G | c.2559G>C (p.Thr853Thr) | homozygous | Myopathy with rimmed ubiquitin-positive autophagic vacuolation, autosomal dominant, 601846 (3) |
| *PLIN4* | V1,V2 | chr19:4511404 | missense_variant | NM_001367868.2 | g.4511404T>G | c.2556A>C (p.Lys852Asn) | homozygous | Myopathy with rimmed ubiquitin-positive autophagic vacuolation, autosomal dominant, 601846 (3) |
| *PLIN4* | V1,V2 | chr19:4511406 | missense_variant | NM_001367868.2 | g.4511406T>C | c.2554A>G (p.Lys852Glu) | homozygous | Myopathy with rimmed ubiquitin-positive autophagic vacuolation, autosomal dominant, 601846 (3) |
| *PLIN4* | V1,V2 | chr19:4511412 | missense_variant | NM_001367868.2 | g.4511412C>T | c.2548G>A (p.Gly850Arg) | homozygous | Myopathy with rimmed ubiquitin-positive autophagic vacuolation, autosomal dominant, 601846 (3) |
| *PLIN4* | V1,V2 | chr19:4511416 | synonymous_variant | NM_001367868.2 | g.4511416T>C | c.2544A>G (p.Gln848Gln) | homozygous | Myopathy with rimmed ubiquitin-positive autophagic vacuolation, autosomal dominant, 601846 (3) |
| *PLIN4* | V1,V2 | chr19:4511419 | synonymous_variant | NM_001367868.2 | g.4511419C>G | c.2541G>C (p.Val847Val) | homozygous | Myopathy with rimmed ubiquitin-positive autophagic vacuolation, autosomal dominant, 601846 (3) |
| *PLIN4* | V1,V2 | chr19:4511422 | synonymous_variant | NM_001367868.2 | g.4511422A>G | c.2538T>C (p.Ala846Ala) | homozygous | Myopathy with rimmed ubiquitin-positive autophagic vacuolation, autosomal dominant, 601846 (3) |
| *PLIN4* | V1,V2 | chr19:4511425 | synonymous_variant | NM_001367868.2 | g.4511425A>C | c.2535T>G (p.Gly845Gly) | homozygous | Myopathy with rimmed ubiquitin-positive autophagic vacuolation, autosomal dominant, 601846 (3) |
| *PLIN4* | V2 | chr19:4511428 | synonymous_variant | NM_001367868.2 | g.4511428C>T | c.2532G>A (p.Lys844Lys) | homozygous | Myopathy with rimmed ubiquitin-positive autophagic vacuolation, autosomal dominant, 601846 (3) |
| *PLIN4* | V1,V2 | chr19:4511434 | synonymous_variant | NM_001367868.2 | g.4511434C>G | c.2526G>C (p.Val842Val) | homozygous | Myopathy with rimmed ubiquitin-positive autophagic vacuolation, autosomal dominant, 601846 (3) |
| *PLIN4* | V1,V2,V4 | chr19:4511443 | synonymous_variant | NM_001367868.2 | g.4511443A>T | c.2517T>A (p.Ala839Ala) | homozygous | Myopathy with rimmed ubiquitin-positive autophagic vacuolation, autosomal dominant, 601846 (3) |
| *PLIN4* | V1 | chr19:4511446 | synonymous_variant | NM_001367868.2 | g.4511446A>C | c.2514T>G (p.Gly838Gly) | homozygous | Myopathy with rimmed ubiquitin-positive autophagic vacuolation, autosomal dominant, 601846 (3) |
| *PLIN4* | V1,V2,V4 | chr19:4511449 | synonymous_variant | NM_001367868.2 | g.4511449G>C | c.2511C>G (p.Thr837Thr) | homozygous | Myopathy with rimmed ubiquitin-positive autophagic vacuolation, autosomal dominant, 601846 (3) |
| *PLIN4* | V1,V2 | chr19:4511450 | missense_variant | NM_001367868.2 | g.4511450G>A | c.2510C>T (p.Thr837Ile) | homozygous | Myopathy with rimmed ubiquitin-positive autophagic vacuolation, autosomal dominant, 601846 (3) |
| *PLIN4* | V1,V2,V4 | chr19:4511454 | missense_variant | NM_001367868.2 | g.4511454C>G | c.2506G>C (p.Val836Leu) | homozygous | Myopathy with rimmed ubiquitin-positive autophagic vacuolation, autosomal dominant, 601846 (3) |
| *STXBP2* | V1,V2,V4 | chr19:7647401 | missense_variant | NM_006949.4 | g.7647401G>C | c.1586G>C (p.Arg529Pro) | heterozygous | Hemophagocytic lymphohistiocytosis, familial, 5, with or without microvillus inclusion disease, 613101 (3) |
| *TYK2* | V1,V2,V3,V4 | chr19:10353614 | missense_variant | NM_003331.5 | g.10353614C>A | c.2941G>T (p.Val981Leu) | heterozygous | Immunodeficiency 35, 611521 (3) |
| *FARSA* | V1,V2,V4 | chr19:12930628 | missense_variant | NM_004461.3 | g.12930628G>A | c.269C>T (p.Ala90Val) | heterozygous | ?Rajab interstitial lung disease with brain calcifications 2, 619013 (3) |
| *JAK3* | V1,V2,V4 | chr19:17834662 | synonymous_variant | NM_000215.4 | g.17834662G>A | c.2259C>T (p.Ala753Ala) | heterozygous | Severe combined immunodeficiency, autosomal recessive, T-negative/B-positive type, 600802 (3) |
| *IL12RB1* | V1,V2,V4 | chr19:18072187 | missense_variant | NM_005535.3 | g.18072187C>T | c.946G>A (p.Val316Met) | heterozygous | Immunodeficiency 30, 614891 (3) |
| *WDR62* | V1,V2,V3,V4 | chr19:36103774 | missense_variant | NM_001083961.2 | g.36103774C>G | c.3946C>G (p.Gln1316Glu) | heterozygous | Microcephaly 2, primary, autosomal recessive, with or without cortical malformations, 604317 (3) |
| *ACTN4* | V1,V2,V4 | chr19:38729303 | synonymous_variant | NM_004924.6 | g.38729303A>G | c.2607A>G (p.Arg869Arg) | heterozygous | Glomerulosclerosis, focal segmental, 1, 603278 (3) |
| *CYP2B6* | V1,V2,V3,V4 | chr19:40991367 | missense_variant | NM_000767.5 | g.40991367A>T | c.62A>T (p.Gln21Leu) | heterozygous | {Efavirenz central nervous system toxicity, susceptibility to}, 614546 (3) |
| *LIG1* | V1,V2,V4 | chr19:48157028 | missense_variant | NM_000234.3 | g.48157028G>A | c.356C>T (p.Pro119Leu) | heterozygous | Immunodeficiency 96, 619774 (3) |
| *TRPM4* | V1,V2,V4 | chr19:49182889 | stop_gained | NM_017636.4 | g.49182889G>A | c.1575G>A (p.Trp525*) | heterozygous | Progressive familial heart block, type IB, 604559 (3) |
| *KCNC3* | V1,V2,V4 | chr19:50323549 | synonymous_variant | NM_004977.3 | g.50323549G>A | c.1404C>T (p.Tyr468Tyr) | heterozygous | Spinocerebellar ataxia 13, 605259 (3) |
| *FERMT1* | V1,V2,V3,V4 | chr20:6096914 | synonymous_variant | NM_017671.5 | g.6096914C>T | c.1077G>A (p.Ala359Ala) | heterozygous | Kindler syndrome, 173650 (3) |
| *MYLK2* | V1,V2,V3,V4 | chr20:31824298 | synonymous_variant | NM_033118.4 | g.31824298C>T | c.918C>T (p.Ala306Ala) | heterozygous | Cardiomyopathy, hypertrophic, 1, digenic, 192600 (3) |
| *AHCY* | V1,V2,V4 | chr20:34295502 | missense_variant | NM_000687.4 | g.34295502G>A | c.112C>T (p.Arg38Trp) | heterozygous | Hypermethioninemia with deficiency of S-adenosylhomocysteine hydrolase, 613752 (3) |
| *PIGU* | V1,V2,V3,V4 | chr20:34575113 | synonymous_variant | NM_080476.5 | g.34575113G>A | c.1185C>T (p.Asn395Asn) | heterozygous | Neurodevelopmental disorder with brain anomalies, seizures, and scoliosis, 618590 (3) |
| *TOP1* | V1,V2,V4 | chr20:41061479 | synonymous_variant | NM_003286.4 | g.41061479G>A | c.144G>A (p.Lys48Lys) | heterozygous | DNA topoisomerase I, camptothecin-resistant (3) |
| *ZNF335* | V1,V2,V4 | chr20:45969600 | missense_variant | NM_022095.4 | g.45969600C>T | c.293G>A (p.Gly98Glu) | heterozygous | Microcephaly 10, primary, autosomal recessive, 615095 (3) |
| *EDN3* | V1,V2,V4 | chr20:59321024 | missense_variant | NM_207034.3 | g.59321024G>T | c.373G>T (p.Val125Leu) | heterozygous | {Hirschsprung disease, susceptibility to, 4}, 613712 (3) |
| *SYCP2* | V1,V2,V4 | chr20:59901786 | missense_variant | NM_014258.4 | g.59901786G>T | c.1058C>A (p.Thr353Lys) | heterozygous | Spermatogenic failure 1, 258150 (3) |
| *LAMA5* | V1,V2,V3,V4 | chr20:62327310 | missense_variant | NM_005560.6 | g.62327310G>A | c.5035C>T (p.Arg1679Trp) | heterozygous | Nephrotic syndrome, type 26, 620049 (3) |
| *RTEL1* | V1,V2,V3,V4 | chr20:63678326 | synonymous_variant | NM_001283009.2 | g.63678326C>T | c.1017C>T (p.Ser339Ser) | heterozygous | Pulmonary fibrosis and/or bone marrow failure syndrome, telomere-related, 3, 616373 (3) |
| *RTEL1* | V1,V2,V3,V4 | chr20:63689775 | missense_variant | NM_001283009.2 | g.63689775G>A | c.2051G>A (p.Arg684Gln) | heterozygous | Pulmonary fibrosis and/or bone marrow failure syndrome, telomere-related, 3, 616373 (3) |
| *TMPRSS15* | V1,V2,V3,V4 | chr21:18353821 | missense_variant | NM_002772.3 | g.18353821G>A | c.923C>T (p.Ser308Phe) | heterozygous | Enterokinase deficiency, 226200 (3) |
| *TIAM1* | V1,V2,V3,V4 | chr21:31120738 | missense_variant | NM_001353694.2 | g.31120738T>C | c.4406A>G (p.Lys1469Arg) | heterozygous | Neurodevelopmental disorder with language delay and seizures, 619908 (3) |
| *SON* | V1,V2,V4 | chr21:33550895 | missense_variant | NM_138927.4 | g.33550895C>T | c.1664C>T (p.Thr555Met) | heterozygous | ZTTK syndrome, 617140 (3) |
| *COL18A1* | V1,V2,V3,V4 | chr21:45504512-45504529 | inframe_deletion | NM_001379500.1 | g.45504512_45504529del | c.2824_2841del (p.942_947del) | heterozygous | Knobloch syndrome, type 1, 267750 (3) |
| *PCNT* | V1,V2,V3,V4 | chr21:46398242 | missense_variant | NM_006031.6 | g.46398242C>G | c.4571C>G (p.Pro1524Arg) | heterozygous | Microcephalic osteodysplastic primordial dwarfism, type II, 210720 (3) |
| *PCNT* | V1,V2,V3,V4 | chr21:46432135 | missense_variant | NM_006031.6 | g.46432135G>A | c.8671G>A (p.Ala2891Thr) | heterozygous | Microcephalic osteodysplastic primordial dwarfism, type II, 210720 (3) |
| *GGT1* | V1,V2,V4 | chr22:24627525 | missense_variant | NM_001288833.2 | g.24627525G>A | c.1114G>A (p.Glu372Lys) | heterozygous | ?Glutathioninuria, 231950 (3) |
| *TRIOBP* | V1,V2,V4 | chr22:37733381 | missense_variant | NM_001039141.3 | g.37733381G>A | c.4031G>A (p.Arg1344Gln) | heterozygous | Deafness, autosomal recessive 28, 609823 (3) |
| *TRIOBP* | V1,V2,V4 | chr22:37768164 | missense_variant | NM_001039141.3 | g.37768164G>A | c.6563G>A (p.Arg2188Gln) | heterozygous | Deafness, autosomal recessive 28, 609823 (3) |
| *SBF1* | V1,V2,V3,V4 | chr22:50459648 | synonymous_variant | NM_002972.4 | g.50459648G>A | c.3510C>T (p.Ile1170Ile) | heterozygous | Charcot-Marie-Tooth disease, type 4B3, 615284 (3) |
| *SYN1* | V1,V2,V3,V4 | chrX:47605277 | synonymous_variant | NM_006950.3 | g.47605277G>A | c.630C>T (p.Ile210Ile) | homozygous | Intellectual developmental disorder, X-linked 50, 300115 (3) |
| *AR* | V1,V2,V3,V4 | chrX:67545317-67545361 | inframe_deletion | NM_000044.6 | g.67545317_67545361del | c.171_215del (p.57_72delinsLeu) | homozygous | {Prostate cancer, susceptibility to}, 301120 (3) |
