## Supplementary material for "The Revised Diploid Genome Sequence of an Individual Human: An Optimized Assembly Workflow for Scaling of near Telomere-to-Telomere Assemblies": SI_13.pdf

SI-13. Seven Variants Classified as Pathogenic or likely Pathogenic by ACMG Guidelines. This table lists the same variants in Figure 5a with the additional information of variant calling method and MANE transcripts used to annotate the variant.

| Gene | Calling method (V1=short reads, V2=ONT long reads, V3=assembly, V4=HiFi long reads) | Location (GRCh38) | Genomic change | MANE transcript | cDNA and protein change | Zygosity | OMIM Phenotype | ACMG Classification |
|---|---|---|---|---|---|---|---|---|
| *DNAH7* | V1,V2,V3,V4 | chr2:195923649-195923650 | g.195923658dup | NM_018897.3 | c.3770dup (p.Asn1257Lysfs*3) | heterozygous | Ciliary dyskinesia, primary, 50, 620356 (3) (AR) | Likely Pathogenic |
| *COL4A4* | V1,V2,V4 | chr2:227052367 | g.227052367G>C | NM_000092.5 | c.2906C>G (p.Ser969*) | heterozygous | Hematuria, familial benign, 1, 141200 (3) (AD) and Alport syndrome 2, 203780 (AR) | Pathogenic |
| *BTD* | V1,V2,V4 | chr3:15644367 | g.15644367G>A | NM_001370658.1 | c.451G>A (p.Ala151Thr) | heterozygous | Biotinidase deficiency, 253260 (3) (AR) | Pathogenic |
| *ACOX2* | V1,V2,V4 | chr3:58534005-58534008 | g.58534010_58534013del | NM_003500.4 | c.461_464del (p.Thr154Serfs*25) | heterozygous | Bile acid synthesis defect, congenital, 6, 617308 (3) (AR) | Likely Pathogenic |
| *LMBRD1* | V1,V2,V3,V4 | chr6:69796818-69796819 | g.69796819dup | NM_018368.4 | c.63dup (p.Leu22Thrfs*14) | heterozygous | Methylmalonic aciduria and homocystinuria, cblF type, 277380 (3) (AR) | Pathogenic |
| *GPR179* | V1,V2,V4 | chr17:38330430-38330431 | g.38330432_38330433del | NM_001004334.4 | c.3138_3139del (p.Glu1046Aspfs*13) | heterozygous | Night blindness, congenital stationary (complete), 1E, autosomal recessive, 614565 (3) (AR) | Likely Pathogenic |
| *RAD51C* | V1,V2,V4 | chr17:58696773 | g.58696773G>A | NM_058216.3 | c.485G>A (p.Gly162Glu) | heterozygous | Fanconi anemia, complementation group O, 613390 (AR){Breast-ovarian cancer, familial, susceptibility to, 3}, 613399 (3) (AD) | Likely Pathogenic |
