## Supplementary material for "The Revised Diploid Genome Sequence of an Individual Human: An Optimized Assembly Workflow for Scaling of near Telomere-to-Telomere Assemblies": SI_14.pdf

SI-14. Fifty-Nine Rare Structural Variants (SV) Overlapping OMIM Phenotype-Associated Genes. These structural variants are rare (<1% population frequency) and overlap the gene region of OMIM phenotype-associated genes. The variants listed correspond to the blue numbers in the Venn diagram of Figure 4b, with the calling method column denoting which Venn diagram region the variant belongs to.

| Gene | Calling method<br>(V1=Illumina short reads,<br>V2=ONT long reads,<br>V3=assembly,<br>V4=HiFi long reads) | Location (GRCh38) | SV size-type | MANE transcript | Transcript / Protein Change | Zygosity | OMIM Phenotype |
|---|---|---|---|---|---|---|---|
| *DVL1* | V1 | chr1:1339884 | 39bp-insertion | NM_001330311.2 | within intronic region | homozygous | Robinow syndrome, 616331 (3) (AD) |
| *PRDM16* | V2,V4 | chr1:3287478-3287621 | 144bp-deletion | NM_022114.4 | within intronic region | heterozygous | Left ventricular noncompaction 8, 615373 (3) (AD); Cardiomyopathy, dilated, 1LL, 615373 (3) (AD) |
| *PER3* | V1,V2,V3,V4 | chr1:7829913-7829966 | 54bp-deletion | NM_001377275.1 | in-frame deletion of 18AA | heterozygous | Advanced sleep phase syndrome, familial, 3, 616882 (3) (AD) |
| *GRHL3* | V2 | chr1:24344974 | 100bp-insertion | NM_198173.3 | within intronic region | heterozygous | van der Woude syndrome 2, 606713 (3) (AD) |
| *FLG* | V2,V4 | chr1:152305958 | 972bp-insertion | NM_002016.2 | in-frame insertion of 324AA | heterozygous | Ichthyosis vulgaris, 146700 (3) (AD) (AR); {Dermatitis, atopic, susceptibility to, 2}, 605803 (3) (AD) |
| *MUC1* | V2,V3,V4 | chr1:155188854-155189218 | 365bp-deletion | NM_002456.6 (non-MANE) | within intronic region | heterozygous | Tubulointerstitial kidney disease, 2, 174000 (3) (AD) |
| *MUC1* | V2,V3,V4 | chr1:155189843-155189902 | 60bp-deletion | NM_002456.6 (non-MANE) | within intronic region | heterzygous | Tubulointerstitial kidney disease, 2, 174000 (3) (AD) |
| *MUC1* | V3,V4 | chr1:155190203-155190379 | 177bp-deletion | NM_002456.6 (non-MANE) | within intronic region | heterzygous | Tubulointerstitial kidney disease, 2, 174000 (3) (AD) |
| *MUC1* | V2,V3 | chr1:155190829-155191398 | 568bp-deletion | NM_002456.6 (non-MANE) | within intronic region | heterzygous | Tubulointerstitial kidney disease, 2, 174000 (3) (AD) |
| *FMN2* | V2,V4 | chr1:240207734 | 129bp-insertion | NM_020066.5 | in-frame insertion of 43AA | heterzygous | Intellectual developmental disorder, autosomal recessive 47, 616193 (3) (AR) |
| *FMN2* | V3 | chr1:240208028 | 132bp-insertion | NM_020066.5 | in-frame insertion of 43AA | heterzygous | Intellectual developmental disorder, autosomal recessive 47, 616193 (3) (AR) |
| *MYT1L* | V3,V4 | chr2:1834942 | 66bp-insertion | NM_001303052.2 | within intronic region | heterzygous | Intellectual developmental disorder, i:AD 39, 616521 (3) (AD) |
| *RGPD5* | V3,V4 | chr2:109794573 | 345bp-insertion | NM_005054.3 | within intronic region | heterzygous | {Encephalopathy, acute, infection-induced, 3, susceptibility to}, 608033 (3) (AD) |
| *AGXT* | V1,V2,V3,V4 | chr2:240869074 | 74bp-insertion | NM_000030.3 | within intronic region | heterzygous | Hyperoxaluria, primary, type 1, 259900 (3) (AR) |
| *IDUA* | V2,V3,V4 | chr4:999688-999773 | 86bp-deletion | NM_000203.5 | within intronic region | homozygous | Mucopolysaccharidosis Is, 607016 (3) (AR); Mucopolysaccharidosis Ih/s, 607015 (3) (AR); Mucopolysaccharidosis Ih, 607014 (3) (AR) |
| *DOK7* | V1,V2,V3 | chr4:3483081-3483168 | 88bp-deletion | NM_173660.5 | within intronic region | homozygous | Fetal akinesia deformation sequence 3, 618389 (3) (AR); Myasthenic syndrome, congenital, 10, 254300 (3) (AR) |
| *DSPP* | V2,V3,V4 | chr4:87615921-87615974 | 54bp-deletion | NM_014208.3 | in-frame deletion of 18AA | heterzygous | Dentinogenesis imperfecta, Shields type III, 125500 (3) (AD); Dentinogenesis imperfecta, Shields type II, 125490 (3) (AD); Dentin dysplasia, type II, 125420 (3) (AD); Deafness (AD) 39, with dentinogenesis, 605594 (3) (AD) |
| *FLT4* | V2 | chr5:180614208 | 170bp-insertion | NM_182925.5 | within intronic region | heterzygous | Hemangioma, capillary infantile, somatic, 602089 (3); Lymphatic malformation 1, 153100 (3) (AD); Congenital heart defects, multiple types, 7, 618780 (3) (AD) |
| *INTS1* | V1,V2,V3,V4 | chr7:1489311 | 51bp-insertion | NM_001080453.3 | within intronic region | homozygous | Neurodevelopmental disorder with cataracts, poor growth, and dysmorphic facies, 618571 (3) (AR) |
| *PSPH* | V1 | chr7:56021043 | 314bp-insertion | NM_004577.4 | within intronic region | heterzygous | Phosphoserine phosphatase deficiency, 614023 (3) (AR) |
| *RP1L1* | V1,V2,V3,V4 | chr8:10610071 | 192bp-insertion | NM_178857.6 | in-frame insertion of 64AA | heterozygous | Occult macular dystrophy, 613587 (3) (AD); Retinitis pigmentosa 88, 618826 (3) (AR) |
| *AGO2* | V2,V3 | chr8:140622888-140623218 | 330bp-insertion | NM_012154.5 | within intronic region | homozygous | Lessel-Kreienkamp syndrome, 619149 (3) (AD) |
| *AGO2* | V2 | chr8:140623136-140623216 | 81bp-deletion | NM_012154.5 | within intronic region | homozygous | Lessel-Kreienkamp syndrome, 619149 (3) (AD) |
| *KANK1* | V2,V3,V4 | chr9:707479-707593 | 115bp-deletion | NM_015158.5 | within intronic region | homozygous | Cerebral palsy, spastic quadriplegic, 2, 612900 (3) |
| *KANK1* | V3 | chr9:708096-708153 | 58bp-deletion | NM_015158.5 | within intronic region | homozygous | Cerebral palsy, spastic quadriplegic, 2, 612900 (3) |
| *TMC1* | V4 | chr9:72700726-72700765 | 40bp-deletion | NM_138691.3 | within intronic region | heterozygous | Deafness, 36, 606705 (3) (AD); Deafness, autosomal recessive 7, 600974 (3) (AR) |
| *MAN1B1* | V2,V4 | chr9:137102262 | 1672bp-insertion | NM_016219.5 | within intronic region | heterozygous | Rafiq syndrome, 614202 (3) (AR) |
| *MAN1B1* | V2 | chr9:137103070 | 110bp-insertion | NM_016219.5 | within intronic region | heterozygous | Rafiq syndrome, 614202 (3) (AR) |
| *STOX1* | V2,V3,V4 | chr10:68827628-68827963 | 336bp-deletion | NM_152709.5 | within intronic region | homozygous | Preeclampsia/eclampsia 4, 609404 (3) (AD) |
| *EPS8L2* | V2,V4 | chr11:720943 | 228bp-insertion | NM_022772.4 | within intronic region | heterozygous | Deafness autosomal recessive 106, 617637 (3) (AR) |
| *MUC5B* | V2,V3 | chr11:1255551-1255668 | 118bp-deletion | NM_002458.3 | within intronic region | homozygous | {Pulmonary fibrosis, idiopathic, susceptibility to}, 178500 (3) (AD) |
| *IGHMBP2* | V1,V2,V3,V4 | chr11:68929815 | 192bp-insertion | NM_002180.3 | within intronic region | heterozygous | Charcot-Marie-Tooth disease, axonal, type 2S, 616155 (3) (AR); Neuronopathy, distal hereditary motor, autosomal recessive 1, 604320 (3) (AR) |
| *MYO7A* | V1,V2,V3 | chr11:77197617 | 140bp-insertion | NM_000260.4 | within intronic region | heterozygous | Deafness, autosomal recessive 2, 600060 (3) (AR); Usher syndrome, type 1B, 276900 (3) (AR); Deafness (AD) 11, 601317 (3) (AD) |
| *DYNC2H1* | V1,V2,V3,V4 | chr11:103177853 | 328bp-insertion | NM_001080463.2 | within intronic region | heterozygous | Short-rib thoracic dysplasia 3 with or without polydactyly, 613091 (3) (DR), (AR) |
| *ACAN* | V2,V3,V4 | chr15:88855767-88855823 | 57bp-deletion | NM_001369268.1 | in-frame deletion of 19AA | heterozygous | Spondyloepiphyseal dysplasia, Kimberley type, 608361 (3) (AD); Short stature and advanced bone age, with or without early-onset osteoarthritis and/or osteochondritis dissecans, 165800 (3) (AD); Spondyloepimetaphyseal dysplasia, aggrecan type, 612813 (3) (AR) |
| *CHSY1* | V1,V2,V3,V4 | chr15:101185413 | 80bp-insertion | NM_014918.5 | within intronic region | homozygous | Temtamy preaxial brachydactyly syndrome, 605282 (3) (AR) |
| *NPRL3* | V2,V3,V4 | chr16:98340 | 37bp-insertion | NM_001077350.3 | within intronic region | heterozygous | Epilepsy, familial focal, with variable foci 3, 617118 (3) (AD) |
| *CAPN15* | V2,V3,V4 | chr16:553056 | 62bp-insertion | NM_005632.3 | within intronic region | homozygous | Oculogastrointestinal neurodevelopmental syndrome, 619318 (3) (AR) |
| *LMF1* | V4 | chr16:902688-903198 | 511bp-deletion | NM_022773.4 | within intronic region | heterozygous | Lipase deficiency, combined, 246650 (3) (AR) |
| *LMF1* | V2,V3 | chr16:903338-903616 | 279bp-deletion | NM_022773.4 | within intronic region | homozygous | Lipase deficiency, combined, 246650 (3) (AR) |
| *LMF1* | V2,V3,V4 | chr16:904195-904240 | 46bp-deletion | NM_022773.4 | within intronic region | homozygous | Lipase deficiency, combined, 246650 (3) (AR) |
| *CTU2* | V3 | chr16:88712915 | 84bp-insertion | NM_001012759.3 | within intronic region | heterozygous | Microcephaly, facial dysmorphism, renal agenesis, and ambiguous genitalia syndrome, 618142 (3) (AR) |
| *PIEZO1* | V1,V2,V3,V4 | chr16:88731922 | 1591bp-insertion | NM_001142864.4 | within intronic region | homozygous | [ER blood group system], 620207 (3) (AR); Lymphatic malformation 6, 616843 (3) (AR); Dehydrated hereditary stomatocytosis with or without pseudohyperkalemia and/or perinatal edema, 194380 (3) (AD) |
| *CHMP1A* | V2,V3,V4 | chr16:89647581 | 588bp-insertion | NM_002768.5 | within intronic region | heterozygous | Pontocerebellar hypoplasia, type 8, 614961 (3) (AR) |
| *CHMP1A* | V2,V4 | chr16:89648436 | 504bp-insertion | NM_002768.5 | within intronic region | heterozygous | Pontocerebellar hypoplasia, type 8, 614961 (3) (AR) |
| *CCDC40* | V1,V2,V3 | chr17:80090387-80090449 | 63bp-deletion | NM_017950.4 | within intronic region | homozygous | Ciliary dyskinesia, primary, 15, 613808 (3) (AR) |
| *FSCN2* | V3 | chr17:81535255 | 159bp-insertion | NM_012418.4 | within intronic region | heterozygous | Retinitis pigmentosa 30, 607921 (3) |
| *MED16* | V3,V4 | chr19:871891 | 3037bp-insertion | NM_005481.3 | within intronic region | heterozygous | Guillouet-Gordon syndrome, 621220 (3) (AR) |
| *PLIN4* | V2,V4 | chr19:4511725 | 198bp-insertion | NM_001367868.2 | in-frame insertion of 66AA | heterozygous | Myopathy with rimmed ubiquitin-positive autophagic vacuolation (AD), 601846 (3) (AD) |
| *KDM4B* | V2,V4 | chr19:5131563 | 245bp-insertion | NM_015015.3 | within intronic region | heterozygous | Intellectual developmental disorder (AD) 65, 619320 (3) (AD) |
| *KDM4B* | V1,V2,V4 | chr19:5144400 | 225bp-insertion | NM_015015.3 | within intronic region | heterozygous | Intellectual developmental disorder (AD) 65, 619320 (3) (AD) |
| *TRPM4* | V2,V3,V4 | chr19:49167643 | 132bp-insertion | NM_017636.4 | within intronic region | heterozygous | Progressive familial heart block, type IB, 604559 (3) (AD); Erythrokeratodermia variabilis et progressiva 6, 618531 (3) (AD) |
| *OSBPL2* | V2,V3,V4 | chr20:62291826 | 60bp-insertion | NM_144498.4 | within intronic region | heterozygous | Deafness (AD) 67, 616340 (3) (AD) |
| *DNAJC5* | V2,V3,V4 | chr20:63919863 | 56bp-insertion | NM_025219.3 | within intronic region | heterozygous | Ceroid lipofuscinosis, neuronal, 4 (Kufs type), (AD), 162350 (3) (AD) |
| *TSPEAR* | V2,V3,V4 | chr21:44591870 | 357bp-insertion | NM_198688.3 | in-frame insertion of 119AA | heterozygous | Tooth agenesis, selective, 10, 620173 (3) (AR); Deafness, autosomal recessive 98, 614861 (3) (AR); Ectodermal dysplasia 14, hypohidrotic/hair/tooth/nail type, 618180 (3) (AR) |
| *COL6A1* | V1,V2,V3,V4 | chr21:45990465 | 318bp-insertion | NM_001848.3 | within intronic region | heterozygous | Ullrich congenital muscular dystrophy 1A, 254090 (3) (AD), (AR); Bethlem myopathy 1A, 158810 (3) (AD) |
| *COL6A2* | V2,V3,V4 | chr21:46111587-46111648 | 62bp-deletion | NM_001849.4 | within intronic region | homozygous | Myosclerosis, congenital, 255600 (3) (AR); Ullrich congenital muscular dystrophy 1B, 620727 (3) (AD), (AR); Bethlem myopathy 1B, 620725 (3) (AD), (AR) |
| *TRIOBP* | V2,V3,V4 | chr22:37724034 | 291bp-insertion | NM_001039141.3 | in-frame insertion of 97AA | heterozygous | Deafness, autosomal recessive 28, 609823 (3) (AR) |
| *SHANK3* | V1,V2,V3,V4 | chr22:50697544 | 284bp-insertion | NM_001372044.2 (non-MANE) | within intronic region | homozygous | Phelan-McDermid syndrome, 606232 (3) (AD); {Schizophrenia 15}, 613950 (3) (AD) |
