## Supplementary material for "The Revised Diploid Genome Sequence of an Individual Human: An Optimized Assembly Workflow for Scaling of near Telomere-to-Telomere Assemblies": SI_15.pdf

### SI-15: Assembly Continuity of Human Genome from One ONT Flowcell

(a)

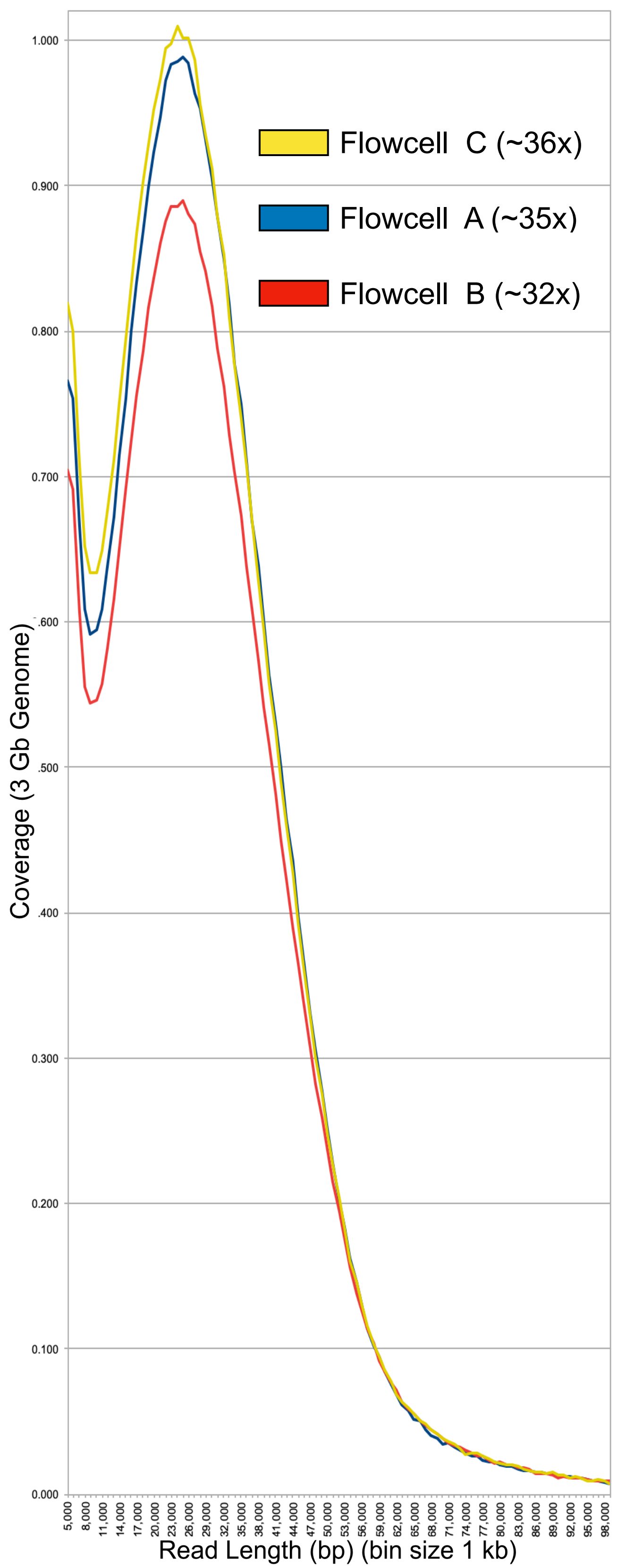

(b)

#### Flowcell A (~35x)

| Sub-Assembly | D | E | H | I | J | K | L | N |
|---|---|---|---|---|---|---|---|---|
| Assembly Size (bp) | 3,045,813,863 | 3,056,788,388 | 3,064,514,992 | 3,063,706,546 | 3,091,085,362 | 3,083,147,597 | 3,065,866,867 | 3,093,153,866 |
| Contig Count | 146 | 153 | 153 | 166 | 218 | 230 | 167 | 220 |
| NG50 (3.0 Gb) (bp) | 85,184,942 | 99,757,501 | 102,647,800 | 132,788,159 | 106,377,225 | 88,209,368 | 96,815,599 | 91,635,406 |
| NG90 (3.0 Gb) (bp) | 23,440,231 | 25,311,022 | 25,326,008 | 25,082,321 | 27,637,374 | 19,563,642 | 21,628,322 | 15,254,842 |
| NG99 (3.0 Gb) (bp) | 4,835,092 | 6,610,508 | 6,475,769 | 9,497,351 | 8,914,334 | 4,818,170 | 5,913,501 | 5,617,134 |
| LG50 (3.0 Gb) | 12 | 11 | 10 | 9 | 10 | 12 | 12 | 12 |
| LG90 (3.0 Gb) | 36 | 31 | 31 | 28 | 29 | 41 | 35 | 38 |
| LG99 (3.0 Gb) | 55 | 47 | 46 | 43 | 44 | 66 | 55 | 64 |
| Mean (bp) | 20,861,739 | 19,979,009 | 20,029,510 | 18,456,064 | 14,179,291 | 13,404,990 | 18,358,484 | 14,059,790 |
| Max (bp) | 203,301,561 | 202,866,167 | 248,301,786 | 248,371,667 | 232,975,508 | 192,053,714 | 191,038,787 | 182,693,416 |
| Min (bp) | 31,857 | 8,289 | 20,795 | 8,289 | 10,270 | 17,140 | 16,607 | 28,014 |
| Chromosomes (no gaps) | 5 | 2 | 3 | 3 | 4 | 2 | 2 | 2 |
| Total Gaps | >91 | >81 | >80 | >74 | >81 | >110 | >99 | >119 |
| Total Gaps (exclude chrY) | 60 | 51 | 50 | 44 | 51 | 80 | 69 | 89 |
| chr1 | 5 | 2 | 0 | **0** | 1 | 2 | 3 | 2 |
| chr2 | 3 | 4 | 3 | **2** | 2 | 4 | 2 | 3 |
| chr3 | **0** | 0 | 0 | 1 | 2 | 3 | 2 | 3 |
| chr4 | 3 | 1 | 2 | 2 | 2 | **0** | 2 | 2 |
| chr5 | 2 | 1 | 1 | **0** | 0 | 3 | 1 | 0 |
| chr6 | 1 | 1 | 1 | **1** | 2 | 2 | 1 | 1 |
| chr7 | 4 | 3 | 3 | 3 | **1** | 6 | 4 | 4 |
| chr8 | 5 | **1** | 3 | 3 | 2 | 5 | 7 | 5 |
| chr9 | 4 | 4 | 4 | **1** | 4 | 3 | 4 | 4 |
| chr10 | **0** | 1 | 0 | 1 | 0 | 0 | 0 | 0 |
| chr11 | **0** | 1 | 2 | 2 | 2 | 4 | 1 | 6 |
| chr12 | **0** | 1 | 1 | 0 | 0 | 2 | 3 | 2 |
| chr13 | 2 | 2 | 1 | 2 | 2 | 2 | 2 | **1** |
| chr14 | 4 | 2 (Chimeric) | 2 | 2 | 4 | **1** | 2 | 2 |
| chr15 | 2 | 2 | 1 | 3 | **0** | 3 | 1 | 2 |
| chr16 | 4 | 6 | **3** | 5 | 4 | 6 | 8 | 5 |
| chr17 | 5 | 4 | 3 | 2 | **1** | 5 | 4 | 8 |
| chr18 | **0** | 0 | 1 | 1 | 1 | 2 | 2 | 1 |
| chr19 | 2 | 2 | 2 | **2** | 5 | 4 | 2 | 7 |
| chr20 | 2 | 2 | 1 | 1 | 1 | 2 | **0** | 1 |
| chr21 | **1** | 2 (Chimeric) | 3 | 2 | 1 | 2 | 2 | 2 |
| chr22 | 2 | 2 | 4 | **1** | 1 | 1 | 1 | 2 |
| chrX | 9 | **7** | 9 | 7 | 13 | 18 | 15 | 26 |
| chrY | >30 | **>30** | >30 | >30 | >30 | >30 | >30 | >30 |

| | |
|---|---|
| chr1 | 0 |
| chr2 | 2 |
| chr3 | 0 |
| chr4 | 0 |
| chr5 | 0 |
| chr6 | 1 |
| chr7 | 1 |
| chr8 | 1 |
| chr9 | 1 |
| chr10 | 0 |
| chr11 | 0 |
| chr12 | 0 |
| chr13 | 1 |
| chr14 | 1 |
| chr15 | 0 |
| chr16 | 3 |
| chr17 | 1 |
| chr18 | 0 |
| chr19 | 2 |
| chr20 | 0 |
| chr21 | 1 |
| chr22 | 1 |
| chrX | 7 |
| chrY | >30 |

| | ONT FC A contigs | ONT FC A chromosomal scaffolds |
|---|---|---|
| Assembly Size (bp) | 3,037,022,651 | 3,037,028,851 |
| Contig/ Scaffold Count | 86 | 24 |
| Complete Chromosomes | 10 | 10 |
| Gaps | 62 | 62 |
| NG50 (3.0 Gb) (bp) | 133,718,319 | 153,026,018 |
| NG90 (3.0 Gb) (bp) | 35,303,050 | 83,144,720 |
| NG99 (3.0 Gb) (bp) | 17,193,060 | 49,908,900 |
| LG50 (3.0 Gb) | 9 | 8 |
| LG90 (3.0 Gb) | 24 | 18 |
| LG99 (3.0 Gb) | 35 | 22 |
| Mean (bp) | 35,314,217 | 126,542,869 |
| Max (bp) | 248,371,667 | 248,371,667 |
| Min (bp) | 31,861 | 18,530,514 |
| Phase Block Count | 3,659 | |
| Phase Block NG50 (bp) | 1,490,519 | |

#### Flowcell B (~32x)

| Sub-Assembly | D | E | H | I | J | K | L | N |
|---|---|---|---|---|---|---|---|---|
| Assembly Size (bp) | 3,057,506,832 | 3,078,654,958 | 3,086,329,236 | 3,092,922,741 | 3,085,748,296 | 3,083,431,670 | 3,086,792,662 | 3,098,281,060 |
| Contig Count | 281 | 283 | 285 | 284 | 356 | 377 | 317 | 468 |
| NG50 (3.0 Gb) (bp) | 72,521,825 | 90,531,966 | 79,876,199 | 88,298,558 | 67,257,601 | 56,531,393 | 54,967,448 | 39,997,658 |
| NG90 (3.0 Gb) (bp) | 11,693,210 | 14,151,838 | 15,502,537 | 16,201,307 | 11,309,602 | 8,429,758 | 9,176,878 | 5,081,952 |
| NG99 (3.0 Gb) (bp) | 2,564,098 | 3,331,752 | 3,342,801 | 3,794,695 | 2,471,966 | 2,089,232 | 2,692,692 | 1,809,099 |
| LG50 (3.0 Gb) | 13 | 12 | 15 | 13 | 15 | 18 | 17 | 24 |
| LG90 (3.0 Gb) | 49 | 44 | 45 | 42 | 52 | 69 | 62 | 102 |
| LG99 (3.0 Gb) | 95 | 83 | 79 | 74 | 98 | 130 | 113 | 194 |
| Mean (bp) | 10,880,807 | 10,878,639 | 10,829,225 | 10,890,573 | 8,667,832 | 8,178,864 | 9,737,516 | 6,620,259 |
| Max (bp) | 171,612,262 | 199,441,607 | 170,295,534 | 170,292,223 | 202,216,172 | 171,538,376 | 157,847,690 | 106,897,330 |
| Min (bp) | 12,636 | 7,043 | 7,235 | 7,044 | 6,239 | 23,696 | 30,910 | 30,910 |
| Chromosomes (no gaps) | 1 | 1 | 1 | 3 | 0 | 1 | 0 | 0 |
| Total Gaps | >135 | >128 | >129 | >127 | >158 | >205 | >178 | >294 |
| Total Gaps (exclude chrY) | 105 | 98 | 99 | 97 | 128 | 175 | 148 | 264 |
| chr1 | 9 | 12 | 7 | **7** | 7 | 9 | 5 (chimeric) | 22 |
| chr2 | 4 | 2 | **1** | 3 | 3 | 10 | 4 | 10 |
| chr3 | 1 | 1 | 4 | 3 | **1** | 2 | 2 | 6 |
| chr4 | 2 | **2** | 3 | 3 | 4 | 3 | 5 | 6 |
| chr5 | 3 | **1** | 3 | 2 | 6 | 6 | 4 | 8 |
| chr6 | 0 | 1 | 0 | **0** | 2 | 0 | 3 | 4 |
| chr7 | 2 | 2 | 2 | **0** | 4 | 4 | 1 | 9 |
| chr8 | 4 | 2 | 4 | **2** | 2 | 3 (chimeric) | 6 | 6 |
| chr9 | 2 | **1** | 3 | 4 | 4 | 8 | 8 | 11 |
| chr10 | 2 | 0 | 1 | **0** | 2 | 7 (chimeric) | 4 | 9 |
| chr11 | 5 | 3 | 3 | **3** | 5 | 4 | 6 | 10 |
| chr12 | 2 | 1 | 2 | **1** | 2 | 3 | 1 | 4 |
| chr13 | **5** | 3 | 4 (chimeric) | 4 | 4 | 6 | 4 (chimeric) | 5 |
| chr14 | 4 (chimeric) | 5 | 4 (chimeric) | 4 | 6 | **5** | 6 | 8 |
| chr15 | **6** | 6 | 5 (chimeric) | 4 | 4 | 6 | 5 | 10 |
| chr16 | **5** | 10 (chimeric) | 13 | 7 | 11 (chimeric) | 11 (chimeric) | 9 | 16 |
| chr17 | 5 | 6 | 7 | **5** | 8 | 13 | 6 | 19 |
| chr18 | **1** | 3 | 1 | 3 | 4 | 3 | 3 | 3 |
| chr19 | 9 | 7 | **6** | 8 | 9 | 8 | 12 | 14 |
| chr20 | 1 | 1 | **1** | 2 | 2 | 4 | 2 | 3 |
| chr21 | 4 | 3 | **2** | 4 | 3 | 2 | 4 (chimeric) | 5 |
| chr22 | 5 (chimeric) | 3 (chimeric) | 3 (chimeric) | 5 | 7 (chimeric) | 7 | **4** | 6 |
| chrX | 24 | 23 | **20** | 23 | 28 | 51 | 34 | 70 |
| chrY | >30 | **>30** | >30 | >30 | >30 | >30 | >30 | >30 |

| | |
|---|---|
| chr1 | 7 |
| chr2 | 1 |
| chr3 | 1 |
| chr4 | 2 |
| chr5 | 1 |
| chr6 | 0 |
| chr7 | 0 |
| chr8 | 2 |
| chr9 | 1 |
| chr10 | 0 |
| chr11 | 3 |
| chr12 | 1 |
| chr13 | 5 |
| chr14 | 5 |
| chr15 | 6 |
| chr16 | 5 |
| chr17 | 5 |
| chr18 | 1 |
| chr19 | 6 |
| chr20 | 1 |
| chr21 | 2 |
| chr22 | 4 |
| chrX | 20 |
| chrY | >30 |

| | ONT FC B contigs | ONT FC B chromosomal scaffolds |
|---|---|---|
| Assembly Size (bp) | 3,029,295,210 | 3,029,309,110 |
| Contig/ Scaffold Count | 163 | 24 |
| Complete Chromosomes | 3 | 3 |
| Gaps | 139 | 139 |
| NG50 (3.0 Gb) (bp) | 111,753,846 | 151,311,756 |
| NG90 (3.0 Gb) (bp) | 21,118,015 | 82,118,234 |
| NG99 (3.0 Gb) (bp) | 2,677,352 | 48,828,961 |
| LG50 (3.0 Gb) | 10 | 8 |
| LG90 (3.0 Gb) | 35 | 18 |
| LG99 (3.0 Gb) | 68 | 23 |
| Mean (bp) | 18,584,633 | 126,221,213 |
| Max (bp) | 202,216,172 | 255,424,760 |
| Min (bp) | 12,835 | 18,476,432 |
| Phase Block Count | 3,669 | |
| Phase Block NG50 (bp) | 1,225,034 | |

#### Flowcell C (~36x)

| Sub-Assemby | D | E | H | I | J | K | L | N |
|---|---|---|---|---|---|---|---|---|
| Assembly Size (bp) | 3,080,096,055 | 3,090,314,120 | 3,083,362,203 | 3,080,075,957 | 3,083,299,506 | 3,082,839,387 | 3,082,575,882 | 3,084,515,396 |
| Contig Count | 157 | 177 | 164 | 179 | 233 | 246 | 167 | 253 |
| NG50 (3.0 Gb) (bp) | 93,741,736 | 104,553,301 | 110,913,785 | 128,573,539 | 74,254,067 | 77,443,702 | 110,617,643 | 61,262,651 |
| NG90 (3.0 Gb) (bp) | 22,595,004 | 33,373,865 | 33,397,445 | 38,637,150 | 21,512,906 | 17,581,182 | 19,923,360 | 13,859,465 |
| NG99 (3.0 Gb) (bp) | 8,609,027 | 7,407,906 | 9,202,824 | 8,282,877 | 5,602,853 | 4,170,994 | 7,763,942 | 3,964,167 |
| LG50 (3.0 Gb) | 12 | 11 | 11 | 10 | 14 | 13 | 9 | 17 |
| LG90 (3.0 Gb) | 34 | 31 | 30 | 27 | 39 | 44 | 31 | 52 |
| LG99 (3.0 Gb) | 50 | 48 | 46 | 41 | 59 | 73 | 49 | 83 |
| Mean (bp) | 19,618,446 | 17,459,402 | 18,800,989 | 17,207,128 | 13,233,045 | 12,531,867 | 18,458,538 | 12,191,760 |
| Max (bp) | 171,492,721 | 197,338,118 | 202,586,169 | 202,506,291 | 188,718,761 | 184,179,097 | 219,899,593 | 188,024,645 |
| Min (bp) | 17,059 | 21,517 | 21,715 | 10,458 | 6,109 | 16,569 | 19,823 | 31,602 |
| Chromosomes (no gaps) | 1 | 5 | 6 | 8 | 1 | 0 | 5 | 0 |
| Total Gaps | >81 | >82 | >74 | >68 | >97 | >116 | >88 | >133 |
| Total Gaps (exclude chrY) | 51 | 52 | 44 | 38 | 67 | 86 | 58 | 103 |
| chr1 | 3 | 2 | 2 | **1** | 3 | 4 | 3 | 9 |
| chr2 | 2 | 2 | 1 | **1** | 2 | 3 | 3 (chimeric) | 4 |
| chr3 | 2 | 0 | 0 | **0** | 1 | 2 | 0 | 3 |
| chr4 | 2 | **0** | 3 | 1 | 2 | 3 | 1 | 3 |
| chr5 | 1 | 1 | 2 | **0** | 2 | 1 | 0 | 1 |
| chr6 | 1 | 1 | 2 | **0** | 2 | 3 | 1 | 6 |
| chr7 | 2 | 1 | **0** | 2 | 2 | 4 | 2 | 5 |
| chr8 | 2 | 2 | 4 | 2 | 2 | **1** | 3 | 7 |
| chr9 | 2 | 4 | **0** | 3 | 5 | 4 | 2 | 4 |
| chr10 | 2 | 0 | 0 | **0** | 1 | 1 | 0 | 1 |
| chr11 | 2 | 0 | 0 | **0** | 2 | 3 | 3 | 1 |
| chr12 | 1 | 0 | 0 | **0** | 1 | 1 | 1 | 2 |
| chr13 | 2 | 2 | **4** | 1 | 2 | 3 | 1 | 1 |
| chr14 | 1 | 1 | 1 | **0** | 3 | 5 | 1 (chimeric) | 3 |
| chr15 | 1 | 3 | 2 | 2 | **1** | 3 | 2 (chimeric) | 4 |
| chr16 | 2 | 4 | 2 | **2** | 2 | 3 | 5 | 1 |
| chr17 | 4 | 3 | 1 | **1** | 4 | 6 | 3 (chimeric) | 4 |
| chr18 | 1 | 1 | **1** | 2 | 1 | 1 | 2 | 1 |
| chr19 | 5 | 8 | **5** | 6 | 7 | 11 | 7 | 6 |
| chr20 | 0 | 1 | 1 | **0** | 0 | 1 | 3 | 2 |
| chr21 | 1 | 2 | 2 | 1 | 1 | 1 | **0** | 2 |
| chr22 | 3 | 2 | 3 | 2 | 3 | 3 | **0** | 4 |
| chrX | 9 | 12 | **8** | 11 | 18 | 19 | 15 | 29 |
| chrY | >30 | **>30** | >30 | >30 | >30 | >30 | >30 | >30 |

| | |
|---|---|
| chr1 | 1 |
| chr2 | 1 |
| chr3 | 0 |
| chr4 | 0 |
| chr5 | 0 |
| chr6 | 0 |
| chr7 | 0 |
| chr8 | 1 |
| chr9 | 0 |
| chr10 | 0 |
| chr11 | 0 |
| chr12 | 0 |
| chr13 | 4 |
| chr14 | 0 |
| chr15 | 1 |
| chr16 | 2 |
| chr17 | 1 |
| chr18 | 1 |
| chr19 | 5 |
| chr20 | 0 |
| chr21 | 0 |
| chr22 | 0 |
| chrX | 8 |
| chrY | >30 |

| | ONT FC C contigs | ONT FC C chromosomal scaffolds |
|---|---|---|
| Assembly Size (bp) | 3,036,666,560 | 3,036,674,260 |
| Contig/ Scaffold Count | 101 | 24 |
| Complete Chromosomes | 13 | 13 |
| Gaps | 77 | 77 |
| NG50 (3.0 Gb) (bp) | 128,573,539 | 154,680,503 |
| NG90 (3.0 Gb) (bp) | 41,530,910 | 82,297,662 |
| NG99 (3.0 Gb) (bp) | 11,236,444 | 50,821,781 |
| LG50 (3.0 Gb) | 10 | 8 |
| LG90 (3.0 Gb) | 25 | 18 |
| LG99 (3.0 Gb) | 35 | 22 |
| Mean (bp) | 30,066,006 | 126,528,094 |
| Max (bp) | 202,506,291 | 247,457,157 |
| Min (bp) | 40,997 | 21,359,553 |
| Phase Block Count | 3,637 | |
| Phase Block NG50 (bp) | 1,392,005 | |

Comparison to T2T-CHM13v2

| | |
|---|---|
| 0 | Chromosome - 0 Gaps (T2T) |
| 1 | Chromosome - 1 Gap |
| 2 | Chromosome - 2 Gaps |
| 3 | Chromosome - 3 Gaps |
| 4+ | Chromosome - > 3 Gaps |
| | Chromosome - 1 or more miss-assembly |
| **N** | Selected for composite assembly |

S15. Assembly Continuity of Human Genome from One ONT Flowcell. (a) Read Length Distribution of Flowcells, A, B, and C. Flowcell B had the lowest coverage in all length categories. (b) Assembly Metrics of the Sub-Assemblies and Final Composite Assembly from single ONT flowcells. Flowcell B produced the worst composite-assembly with 139 gaps, compared to the 62 and 77 gaps from Flowcell A and C, respectively.
