## Supplementary material for "The Revised Diploid Genome Sequence of an Individual Human: An Optimized Assembly Workflow for Scaling of near Telomere-to-Telomere Assemblies": SI_16.pdf

### SI-16: Length distribution of HuRef2.0 Phase-Blocks and Switch-Over Regions

(a)

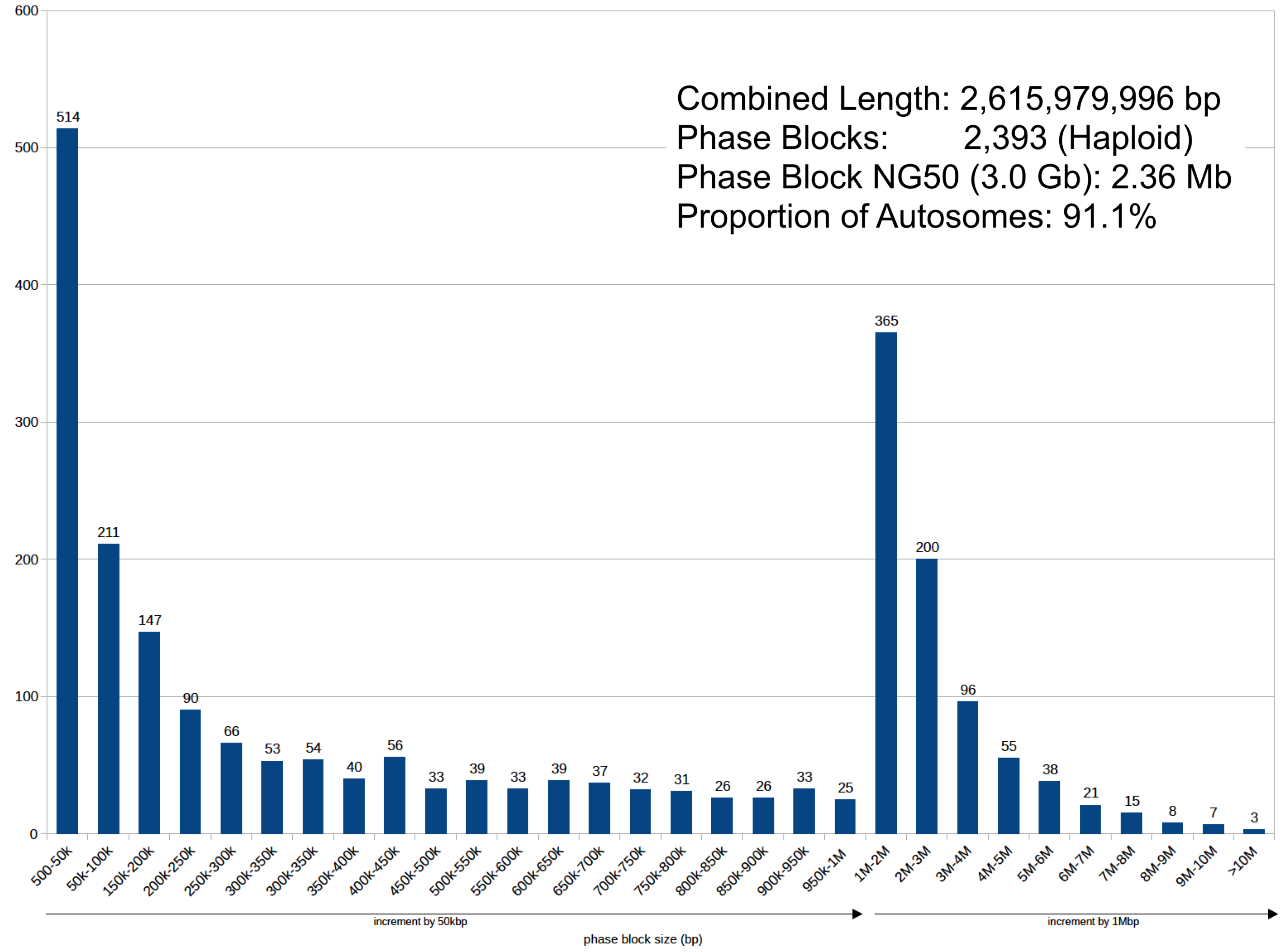


(b)

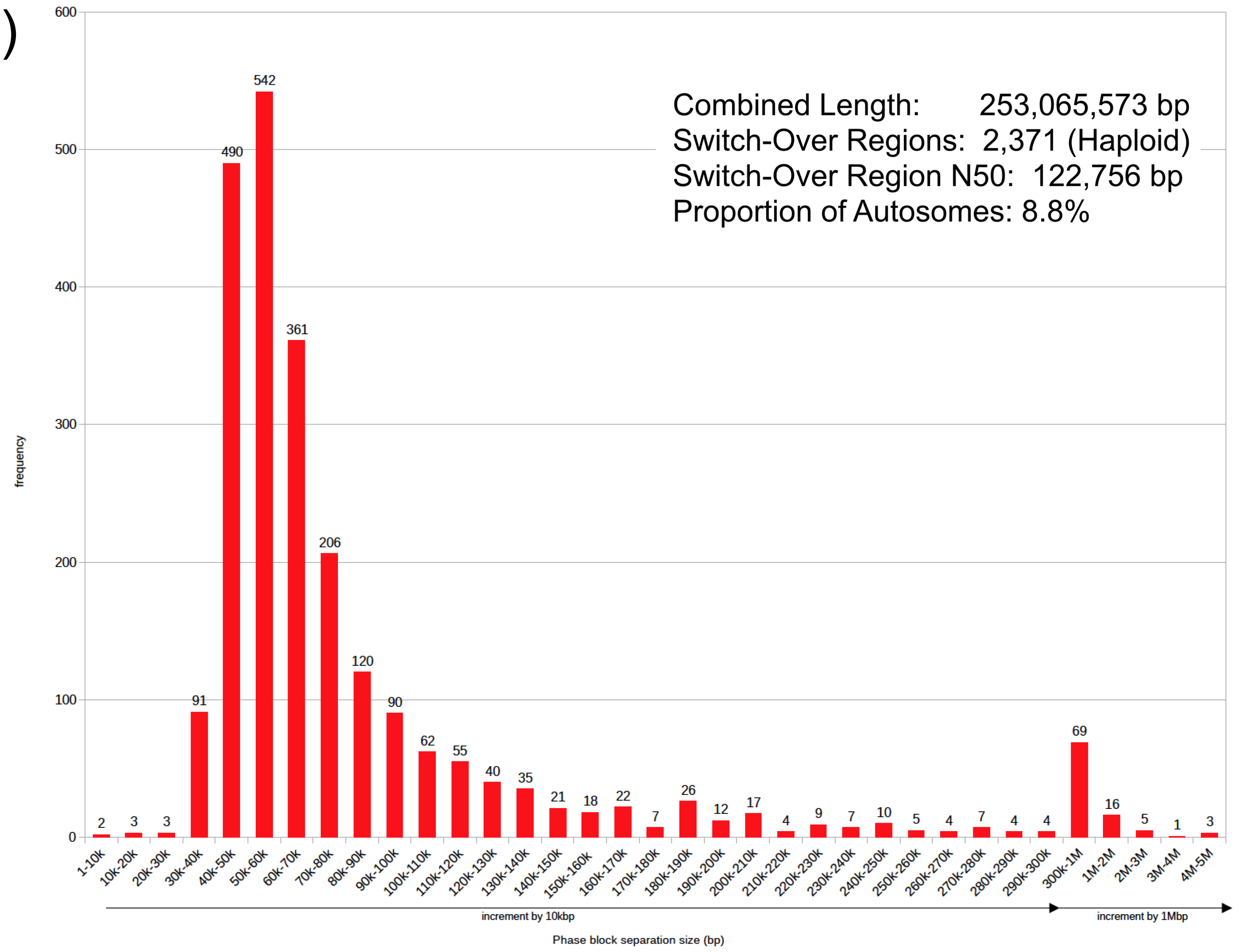

SI-16. Length Distribution of Phase Blocks and the Switch-Over Regions. Over 75% of switch-over regions are less than 100 kb in length, reflecting the phasing potential of ultra-long ONT-reads in future work.
